# Clinical practice competencies for standard critical care nursing: Consensus statement based on a systematic review and Delphi survey

**DOI:** 10.1101/2022.07.24.22277674

**Authors:** Hideaki Sakuramoto, Tomoki Kuribara, Akira Ouchi, Junpei Haruna, Takeshi Unoki, the Committees of Nursing Education and Critical Care Nursing and Working group for Critical Care Nurse Survey Working Group, the AdHoc Committee of Intensive Care Registered Nurse, Japanese Society of Intensive Care Medicine

**Affiliations:** Department of Critical care and Disaster Nursing, Japanese Red Cross Kyushu International College of Nursing, Munakata, Fukuoka, Japan; Department of Acute and Critical Care Nursing, School of Nursing, Sapporo City University, Sapporo, Hokkaido, Japan; Department of Adult Health Nursing, College of Nursing, Ibaraki Christian University, Hitachi, Ibaraki, Japan; Intensive Care Unit, Sapporo Medical University Hospital, Sapporo, Hokkaido, Japan

**Keywords:** clinical practice competency, intensive care unit, critical care nursing

## Abstract

Clinical practice competencies in standard critical care nursing (SCCN) are necessary to improve the quality of care and patient outcomes. Competency enables definition and provides a framework for the evaluation of actual knowledge, skills, and abilities. However, a clear development process and scientifically validated competencies have not yet been developed in Japan. Thus, this study aimed to develop a consensus-based set of SCCN competencies to cover a framework for critical care nursing education, training, and evaluation. A consensus-based set of SCCN competencies was developed in four stages: (1) development of an initial set of SCCN competencies derived from a systematic review; (2) focus group interviews via video conference to supplement and content expert validation based on initial competencies made from a systematic review; (3) a three-round web-based Delphi survey of health professionals to prioritize and gain consensus on the most essential SCCN competencies; and (4) external validation, feedback, and endorsement from critical care experts. A systematic review of 23 studies and reports identified 685 unique competencies. Of the 239 participants representing a range of health professionals (physicians, nurses, and physical therapists) who registered, 218 (91.2% of registered professionals), 209 (98.9% of round 1 participants), and 201 (96.2% of round two participants) participants responded in round one, round two, and round three of the Delphi survey, respectively. The withdrawal rates between enrollment and each round were less than 10%. After three rounds of the Delphi survey and external validation by experts, the final set of competencies was classified into 6 domains, 26 subdomains, 99 elements, and 525 performance indicators. In summary, a consensus-based, contemporary set of SCCN competencies was identified to cover a framework for critical care nursing education, training, and evaluation.

**Trial registration:** None

## Introduction

Critical care nursing is a specialty that deals with specific human responses to actual or potentially life-threatening problems. [1] Critical care is defined by the World Federation of Societies of Intensive and Critical Care Medicine as “a multidisciplinary and interprofessional specialty dedicated to the comprehensive management of patients having, or at risk of developing, acute, life-threatening organ dysfunction”. [2] In recent years, intensive care medicine has undergone significant changes because of the increasing elderly population, complexity, and advances in medical equipment. [2] It is also becoming a discipline that serves the needs of survivors through the issue of post-intensive care syndrome. [3] As a result, critical care nurses must have more complex competencies in the intensive care unit (ICU).

However, the education of critical care nurses requires a long-term training process and is unable to meet rapidly increasing demands, such as disasters. [2, 4] The shortage of critical care nurses worldwide during the COVID-19 pandemic became a serious issue. [4] In Japan, there is no system to identify the number of nurses who could provide standard critical care, and thus, it is impossible to determine the actual shortage of nurses and from where they should be supplied. [5] One of the reasons these issues are highlighted is the lack of clinical practice competencies for standard critical care nursing (SCCN) in Japan.

Competencies are generally defined as a combination of knowledge, skills, attitudes, and values that support effective and efficient performance in professional or occupational areas. [6, 7] A competency framework is a range of required behaviors that provide structural guidelines that enable admission, development, education, training, and evaluation. [7] Therefore, by identifying competencies, SCCN competency enables the definition and provides a framework for the evaluation of actual knowledge, skills, and abilities in the practice of critical care. [8, 9] In addition SCCN competencies would lead to the development of a system to register critical care nurses with those competence characteristics. [5, 7] There are already several national and international clinical practice competencies for critical care nurses. [8, 9] However, a clear development process and scientifically validated competencies have not been previously developed in Japan. In addition, SCCN competency is strongly influenced by sociocultural factors related to healthcare and era. [10]

Therefore, it is necessary to develop a scientific method for identifying the characteristics of SCCN competencies in Japan. This study aimed to develop a consensus-based set of SCCN competencies for teaching, learning programs, and a framework for the evaluation of critical care nursing. The standardized education provided to critical care nurses also challenges many countries because of differences in the era and healthcare culture. [8, 11] Therefore, a detailed description of the design of this study and its results, as well as other competencies, could be used in many countries as a framework for standardized education of critical care nurses and a resource for future research. [8, 11, 12]

## Materials and Methods

### Study design

This study was conducted as a multi-step modified Delphi study with reference to previous studies. [13] First, a systematic review (SR) was conducted to construct the initial competencies that include related potential competencies. Second, focus group interviews were conducted with expert nurses for supplementary and content expert validation. Third, a modified three-round Delphi survey was performed using an internet-based questionnaire to reach a consensus among critical care nurses. Finally, feedback on the final competencies was obtained from external experts (Figure 1).

**Figure 1.**
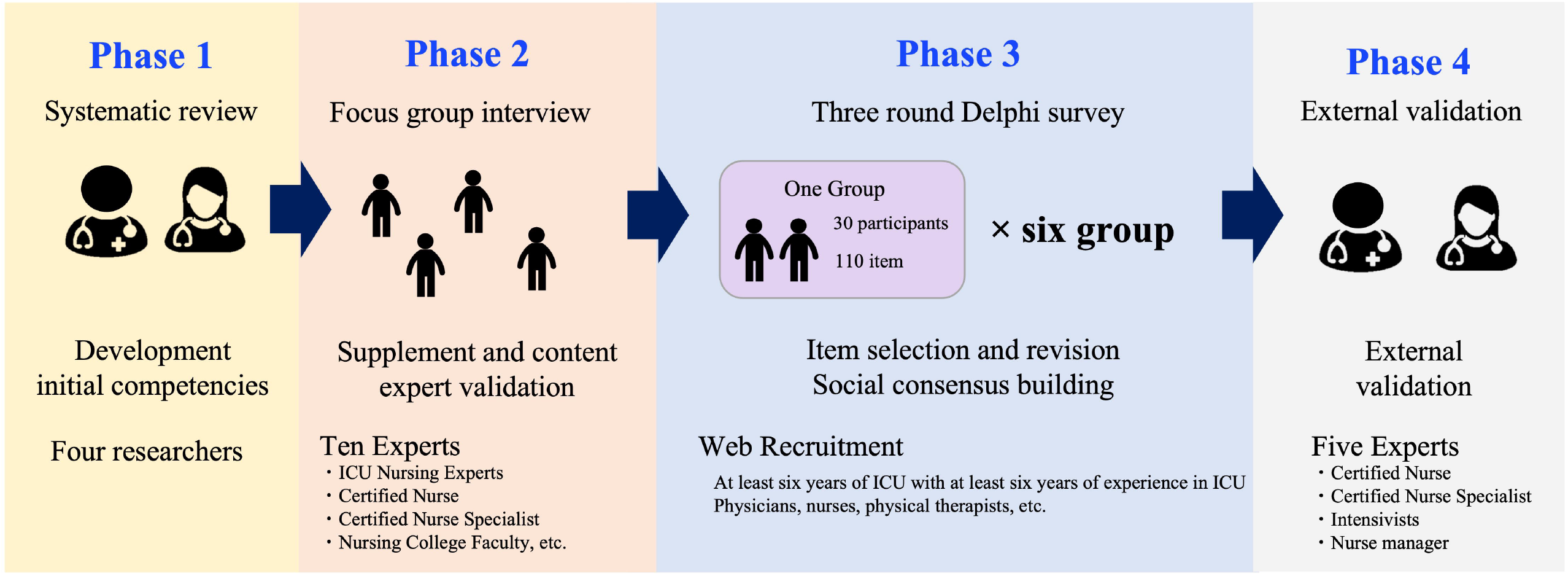
Overall research methods The overall research methods of the study are shown: A consensus-based set of SCCN competencies was developed in four stages.

This study was behalf of the Committees of Nursing Education and Critical Care Nursing and Working group for Critical Care Nurse Survey Working Group and the AdHoc Committee of Intensive Care Registered Nurse, the Japanese Society of Intensive Care Medicine (JSICM).

### Development of initial competencies based on SR

We conducted an SR according to the detailed methodology presented in S1 Text and S2 Text of the Supplement. The eligibility criterion was competencies related to SCCN. MEDLINE using PubMed, CINAHL, and Igaku-Chuo-Zasshi (Ichu-shi) were manually searched for related studies. Ichu-shi is a Japanese medical database that is managed by the Japan Medical Abstract Society. Only studies written in Japanese or English were included. Two author groups (HS and TK, AO and JH) independently screened the titles and abstracts for inclusion eligibility. After screening, two authors independently assessed the full text to identify eligible literature. Disagreements were resolved through a discussion. Subsequently, one of the authors (TK) extracted the competencies from the eligible literature. We translated all competencies into Japanese and reviewed this competency set as the initial competencies for duplication, overlap, and clarity. The research team then classified the words or phrases extracted from the literature into different themes and abstraction levels such as nursing practice and communication. The researchers ultimately classified the domains, subdomains, elements, and performance indicators at the four abstractional levels.

### Focus group interview

We conducted focus group interview (FGI) with expert nurses (certified nurses or certified nurse specialists) or researchers of critical care nursing who had experience in ICU nursing for over 10 years. This FGI was conducted to supplement and validate expert content based on the initial competencies of the SR. A total of 10 participants were recruited for the FGI using purposive and snowball sampling methods. FGI was conducted in two groups involving five members per group, for approximately 60 minutes, using ZOOM®□ (Zoom Video Communications, Inc, San Jose, Calif). The participants who wanted to join the FGI submitted their personal information through the internet. All the researchers were trained beforehand and decided on their roles for the day. FGI was recorded using the recording function of Zoom, and the interviews were transcribed. A qualitative analysis of the verbatim transcript was then performed in three steps. First, we created a code that was shortened to a point where the meaning of the sentence could be understood. Second, the codes and selected keywords regarding clinical practice competencies for SCCN were organized from the FGI. Third, the organized codes and selected keywords from the FGI results were compared with the initial set of competencies obtained in the SR. Competency items for initial competencies were added or revised as needed.

### Three-round Delphi survey

A modified three-round Delphi survey was conducted to reach consensus about SCCN competencies among healthcare professionals who work in critical care settings. [14] The invitation was distributed via the mailing lists of the JSICM and the Japan Society of Education for Physicians and Trainees in Intensive Care. An invitation was also posted in community mailing lists and social network services such as the Japan Association of Certified Intensive Care Nurses, Twitter, and Facebook. Only healthcare professionals who had experience working in the ICU for over 6 years were eligible for the modified Delphi survey. Data were collected from December 4, 2021 to February 10, 2022. Owing to the large number of items, the initial competencies were divided into six separate groups. We planned to include 40 participants in each group for a total of 240 participants, assuming 10 dropouts in each group through the three rounds.

Survey Monkey (Momentive Inc. Sa Mateo, CA, USA) web-based survey service was used for all three rounds of Delphi survey. Participants rated each SCCN competency using a visual analog scale (VAS) anchored with two descriptors labeled “not needed at all” at the far left (0) and “fully needed” at the far right (100), and they wrote free comments. In the first and second rounds, we decided to obtain a consensus for each competency using a median VAS score of >70. In the third round, consensus was obtained based on a median VAS score ≥ 80. A post-meeting to discuss the results of the Delphi round was conducted by the researchers after each Delphi round. In the post meeting, revisions or deletions for competency items that did not reach consensus based on the value of VAS were discussed, based on the free comments.

### External validation

The pre-final set of competencies was sent to five experts (one physician and four nurses) to obtain feedback and ensure the validity, applicability, utility, and clarity of competencies. These five experts were recruited using purposive sampling. The manuscript was revised based on comments from experts, the revised pre-final set of competencies were resent, and a consensus was obtained from all experts.

### Ethical considerations

This study was approved by the Research Ethics Committee of Sapporo City University (approval number:2135-1). For the FGI, Delphi survey, and external validation, consent was obtained from potential participants through the internet format after the participants received adequate explanation of the study.

## Results

### Generation of an Initial Set of Relevant SCCN competency

A total of 685 SCCN competencies were identified in the SR. These competencies were classified into 6 domains, 29 subdomains, 111 elements, and 639 performance indicators after removing duplicates (S3 text, S1 Table, and S3 Table in the Supplementary Material). The two FGIs were conducted by 12 experts. One expert withdrew from the interviews. The characteristics of the experts who conducted the FGIs are shown in S4 Table. The FGI resulted in the addition of three performance indicator items, a synthesis of 2 subdomains, and 10 elements. Revisions were also conducted to the SCCN’s competency representation by FGIs.

### Three-round Delphi survey

The demographic characteristics of the participants are presented in Table 1. Among the registered professionals, 53.6% were female, and the median (IQR) healthcare work and ICU work experiences were 15 (11–20) and 10 (8–13) years, respectively. Of the 239 professionals who registered, 218 participants (91.2% of registered professionals) responded in round one of the Delphi survey, 209 participants (98.9% of round 1 participants) responded in round two of the Delphi survey, and 201 participants (96.2% of round two participants) responded in round three of the Delphi survey. The withdrawal rates between enrollment and each round were less than 10% each (Table1 and Figure 2).

**Table 1.**
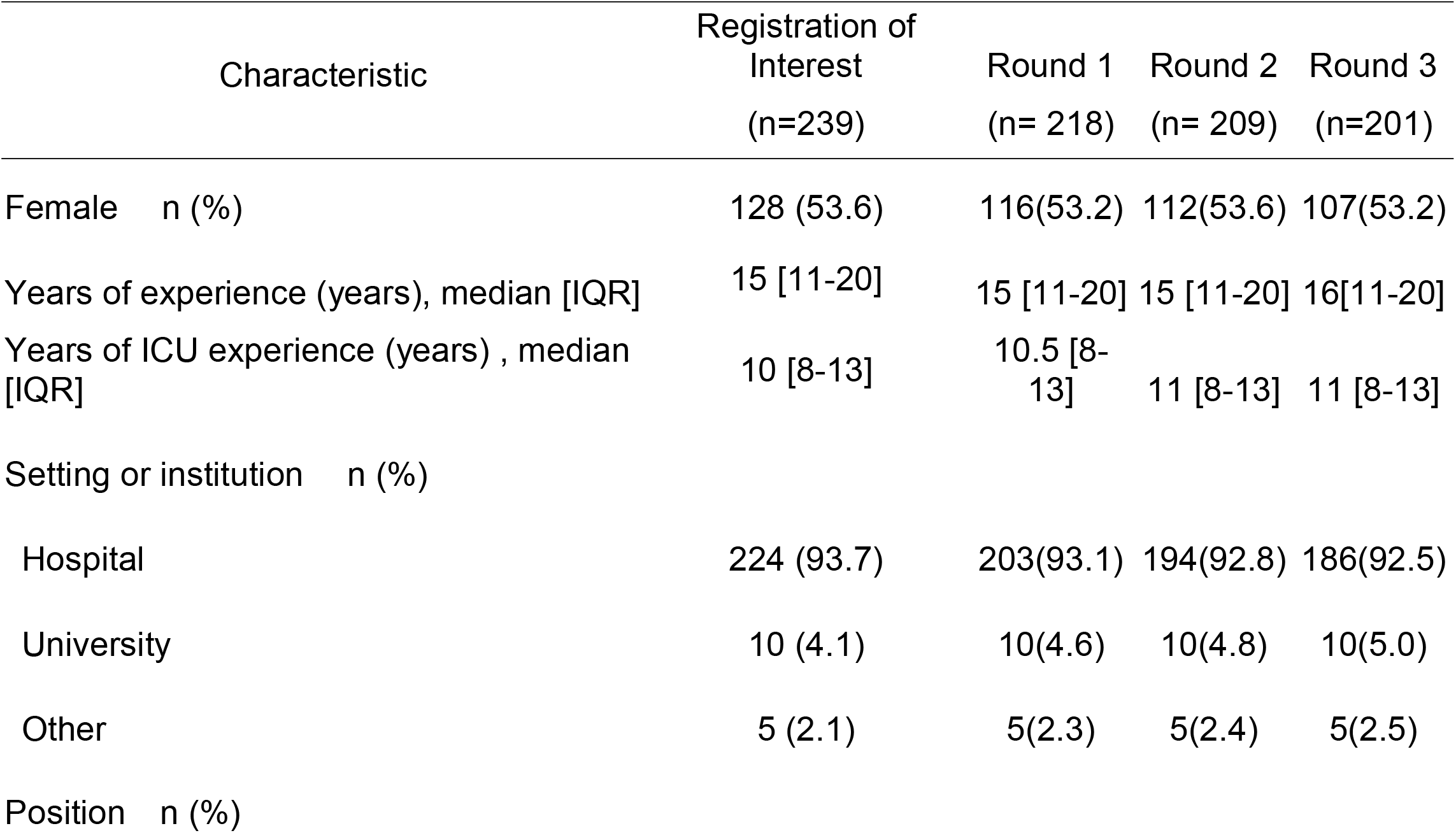
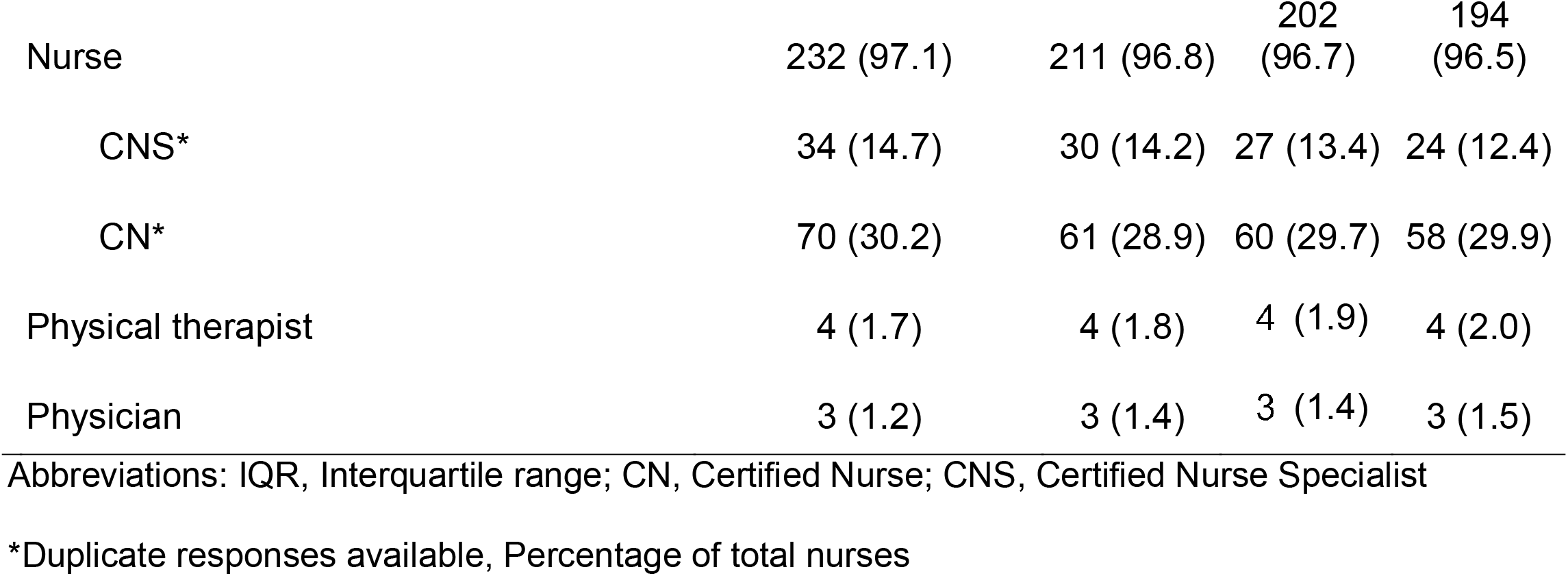
Characteristics of Delphi Round Participants

**Figure 2.**
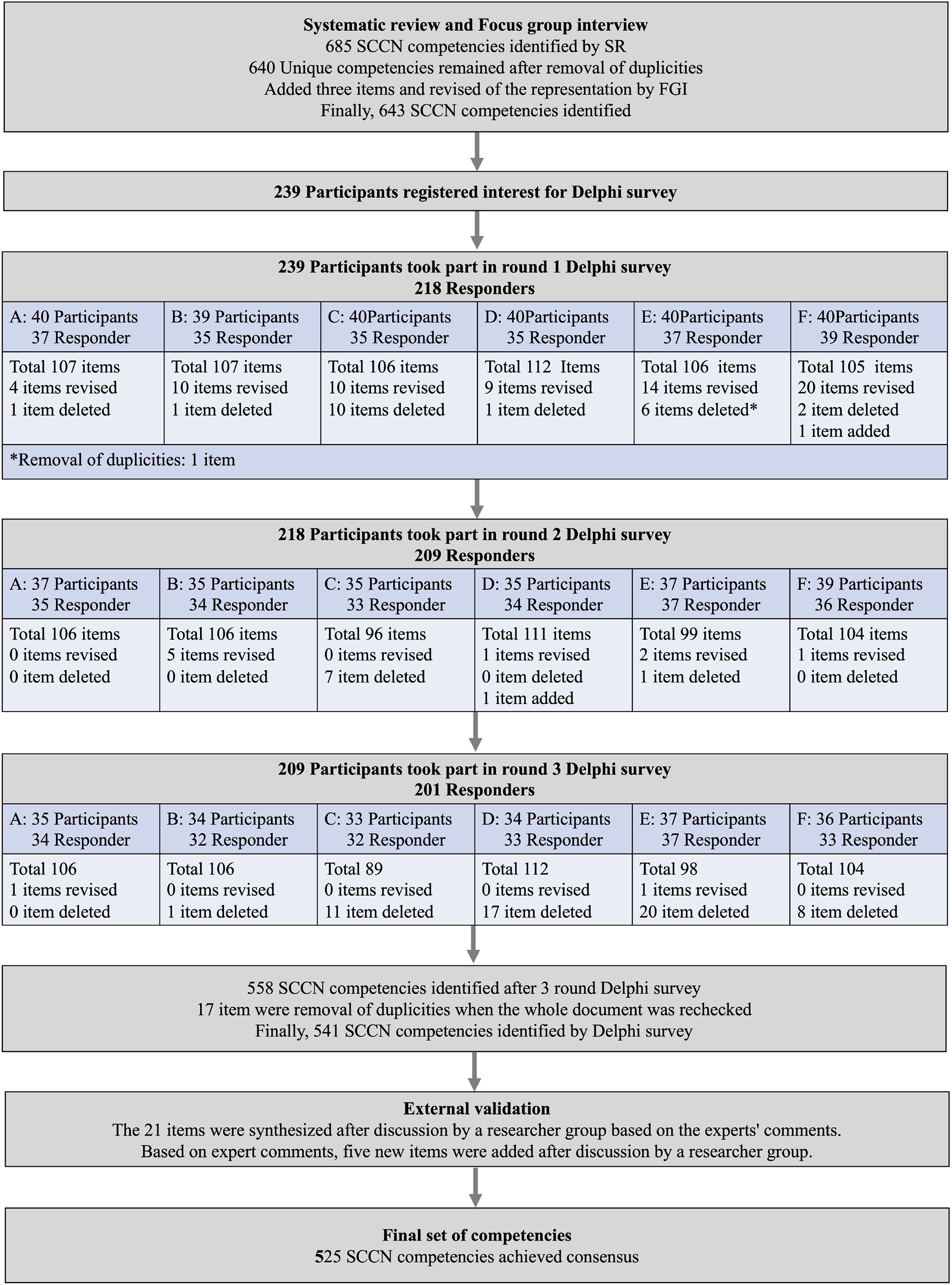
Flow diagram for development of competencies The number of competencies since the systematic review is shown in Figure. The number of competencies revised, deleted, or added to each group in the three Delphi rounds is indicated. The number of withdrawals for each round of participants is shown.

After round one, there were 89 items that were below the consensus level (VAS < 70) or required revision based on the free comments. After discussions among the researchers, 22 items were deleted, and 67 items were revised. In addition, a new item was added based on the free comments. After round two, 17 items were below the consensus level (VAS < 70) or needed revision based on free comments. After discussions among the researchers, eight items were deleted, and nine items were revised. In addition, a new item was added based on free comments. After round three, 57 items were below the consensus level (VAS < 70) and were deleted in the final round. When the entire document was rechecked, 17 items were additionally deleted because they were duplicates. After discussions among the researchers based on the free comments, two items needed to be revised, and one item was added. Figure 2 illustrates the results of the modified Delphi survey (Figure 2). S5 Table in the Supplement presents the detailed results for each round. No revisions were made to the domain subdomain elements.

### External validation

Based on the expert comments, 21 performance indicator items were synthesized, 5 performance indicator items were added, and 60 performance indicator items were revised for representation; 1 subdomain and 2 elements were synthesized after discussion among the researchers. Feedback was received from experts after two revisions to ensure that the final competencies were valid, applicable, useful, and clear. After three rounds of the Delphi survey and external validation by experts, the final set of competencies was classified into 6 domains, 26 subdomains, 99 elements, and 525 performance indicators. Tables 2, S6, and S7 show the final SCCN competencies.

**Table 2.**
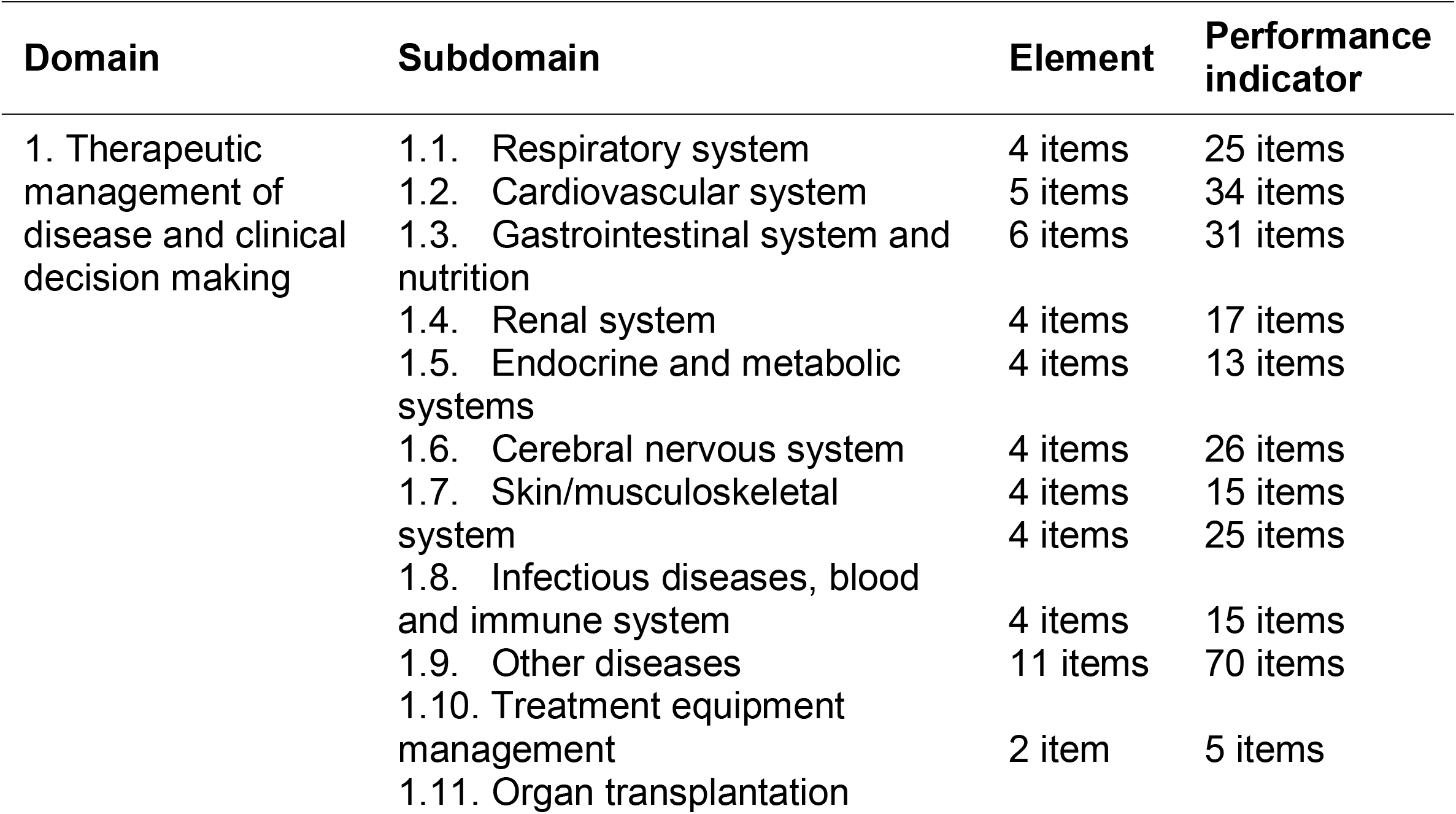
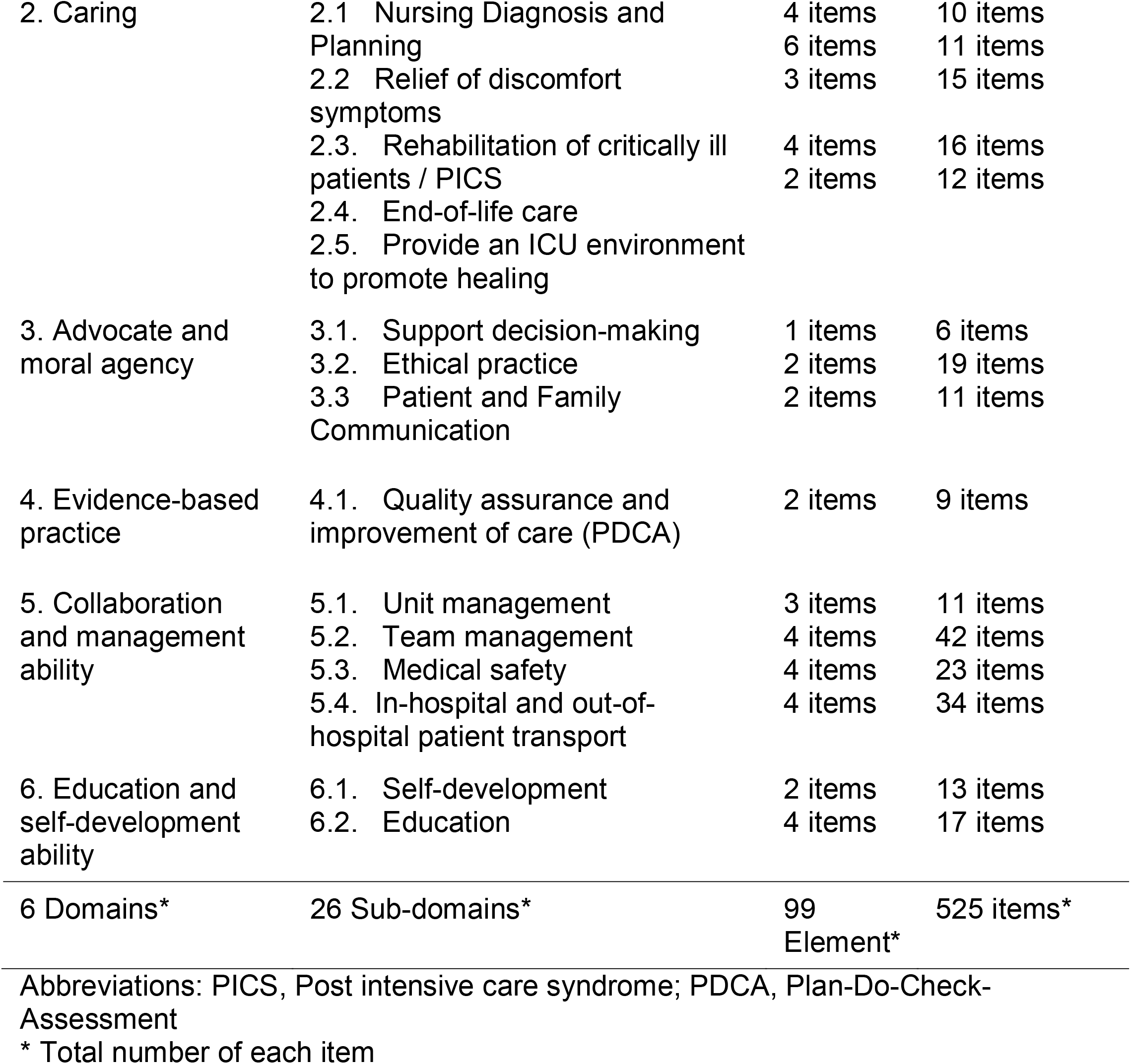
Final set of competencies

## Discussion

In this study, the expert panel reached consensus on the importance of 541/643 competencies for SCCN, after a third round of the Delphi survey. Subsequent revisions through external validity assessment resulted in 525 competencies. Finally, the developed clinical practice competencies for SCCN were categorized into six domains: therapeutic management and clinical judgment; caring, advocate, and moral agency; evidence-based practice; collaboration and management ability; and educational and self-development ability.

The reliability of the results was ensured by the study methodology and design. Delphi results are evaluated with respect to trustworthiness rather than validity, as in quantitative surveys. Trustworthiness encompasses “subconcepts” that consist the components of credibility, transferability, confirmability, and dependability. [15, 16] Previous studies have developed a set of slandered critical care competencies, but it did not use a SR to inform the Delphi survey. [8, 9, 17] The credibility of the findings was also ensured by using previous relevant studies and by the number and expertise of the panelists, who represented various professional groups in critical care. Experts were selected to provide diverse perspectives beyond the nurses themselves to achieve a consensus on the competency framework.

The findings were also generalizable to other settings. Particularly, the clinical practice competencies for SCCN were created from a wide range of experts in the country. Therefore, the competencies obtained in this study can be generalized to critical care nursing throughout Japan. In addition, as the panel of participants changes, so will the interests and opinions of the group, and thus, the results of future studies. The confirmability of the findings (i.e., the ability to ensure data comes from an identifiable source) was ensured through an iterative study design. Results from each round were summated and shared with the study participants, and their feedback on the findings was encouraged. A constant dialogue of data between the researcher and the participant was maintained throughout the study. [16] Confirmability was verified using a replicated study design, as all data were obtained from identifiable sources.

The dependability of the findings (i.e., repeatability in other studies) was ensured with a detailed description of the study design. The dependability of a study result is indicated the repeatability of the results in other studies. [15, 16] In this study, five experts were included to ensure the accuracy of the findings; these experts were asked to review the final competency framework, ensuring dependability. The findings of this study are robust, with low attrition rates and findings from experts across several regions in Japan. The attrition rate at each stage of the Delphi survey is problematic. [18] In the present study, a high response rate was achieved, with an attrition rate of less than 10% over the three rounds. In addition, the experts recruited for this study were active in various areas of critical care and were able to ensure that the competency framework constructed was comprehensive and specific to SCCN practices. Therefore, the results of this study are robust, and the competency framework can be used in multiple areas to improve clinical practice, including the assessment, training, and certification of standard critical care nurses.

When the competency frameworks implemented in this study were compared with those of developed countries, the six domains generally overlapped with the existing competency frameworks that assessed SCCN characteristics. An SR developed the main framework based on previous studies, which was then adjusted to fit the national legislation and the needs of patients’ families. Therefore, the domains were emphasized as cultivating caring, advocacy, altruism, and humanity as well as patient treatment management, physical assessment, and clinical judgment, as in other countries. [8, 17] With respect to differences, there were differences in the level of practice by law and in the performance indicator level according to the needs of the population. Multicultural considerations are common in critical care nursing practice in developed countries. In contrast, most Japanese patients are homogeneous [10], and thus, cultural considerations are less emphasized.

## Strengths and limitations

A key strength of this study is the SR and Delphi survey approach to achieving national consensus on a contemporary set of SCCN competencies. However, our study had some limitations. First, although the Delphi participants were selected to represent a multiplicity of health professions and expertise, but they may not adequately represent the full range of views held by professionals. In addition, the competency in which consensus was reached in this study is the necessity to consider cultural influences on patient attitude toward health, illness, compliance, and care. [10] Second, the competencies are broad and highly detailed, reflecting the scope of work that a standard critical care nurse is expected to accomplish. Therefore, the competency framework may ultimately need to be shortened to improve its learning curve and applicability to clinical practice, in conjunction with professional needs.

## Clinical implications and further research

The competency framework in which consensus was achieved in this study can be used in multiple areas to improve clinical practice, including the assessment, training, and certification of standard critical care nurses. By contrast, we view this set of standard critical care competencies as a dynamic set that reflects the current state of healthcare. As the field matures, new competencies will need to be added and others removed. Therefore, this set of competencies should be revised regularly. The detailed methodology presented will be a useful reference for future research.

## Conclusion

This study found a set of SCCN competencies categorized into 6 domains, 26 subdomains, 99 elements, and 525 performance indicators after a multi-step, modified Delphi study. The results of this study are robust, and the competency framework can be used in multiple areas to improve clinical practice, including the assessment, training, and certification of standard critical care nurses.

## Data Availability

All data produced in the present study are available upon reasonable request to the authors

## Acknowledgements

We thank the members of the Japanese Society of Intensive Care Medicine’s Committees of Nursing Education, Critical Care Nursing, Critical Care Nurse Survey Working Group, and AdHoc Committee of Intensive Care Registered Nurses for their cooperation in this survey (S4 Test, Contributors list).

## CRediT authorship contribution statement

**Hideaki Sakuramoto:** Conceptualization, methodology, validation, investigation, data curation, formal analysis, writing–original draft, project administration. **Tomoki Kuribara:** Conceptualization, methodology, validation, investigation, data curation, formal analysis, writing–original draft, project administration. **Akira Ouchi:** Conceptualization, methodology, validation, investigation, data curation, formal analysis, writing, review, and editing. **Junpei Haruna:** Conceptualization, methodology, validation, investigation, data curation, formal analysis, writing-review, and editing. **Takeshi Unoki:** Conceptualization, methodology, writing–review, and editing.

## Supplementary Information

S1 Text. eMethods_Systematic review for constructing initial competencies

S2 Text. Search terms.

S3 Text. eReferences for included studies in SR

S4 Text. Contributor list

S1 Table. Characteristics of included studies in SR

S2 Table. PRISMA 2020 Checklist

S3 Table. PRISMA Flow chart

S4 Table. Characteristics of participants in the focus group interview

S5 Table. Summary of Delphi survey

S6 Table. Final set of English version of SCCN competencies

S7 Table. Final Set of Japanese version SCCN Competencies

