## Supplementary material for "Clinical practice competencies for standard critical care nursing: Consensus statement based on a systematic review and Delphi survey": S1 table

**eTable 1. Characteristics of included study of systematic review**

| **Author, year** | **Country, language** | **Main results** |
| --- | --- | --- |
| **Papers** | | |
| Canfield, 1981 | US, English | Perceptions of clinical competencies for critical care nurses between baccalaureate educators and nursing employers |
| Inman, 1991 | US, English | Six domains of competency |
| Scribante, 1996 | US, English | Four domains of competency and 25 subdomains |
| Underwood, 1996 | Australia, English | Explanation of Confederation of Australian Critical Care Nurses Inc. Competency Standards for Specialist Critical Care Nurses |
| Dunn, 2000 | Australia, English | Six domains of competency and 20 subdomains |
| Bench, 2003 | UK, English | Four domains of competency |
| Bourgault, 2004 | US, English | Five domains of competency |
| McLean, 2005 | UK, English | Differences between the mentors and students perceptions of competencies |
| Becker, 2006 | US, English | Differences in the practice of clinical nurse specialists and acute care nurse practitioners |
| American Journal of Electroneurodiagnostic Technology, 2008 | US, English | Two section, 16 subdomains of competency about ICU/cEEG |
| Nel, 2011 | South Africa, English | Seven domains of competency and 25 items |
| Sugita, 2012 | Japan, Japanese | Concept analysis of expertise for critical care nurses |
| Hadjibalassi, 2013 | Nicosia, English | Four domains of competency and 72 items |
| Price, 2013 | UK, English | Explanation of national critical care competency framework |
| Gill, 2016 | Australia, English | Four domains of competency and 15 subdomains |
| Deacon, 2017 | UK, English | Explanation of the national critical care competency framework |
| Zhang, 2018 | China, English | Six domains of competency and 92 items |
| Shunman, 2020 | US, English | Three domains of competency |
| **Academic society (Hand search)** | | |
| American Association of Critical-Care Nurses, 2015 | US, English | Six domains of competency and 28 items |
| Australian College of Critical Care Nurses, 2015 | Australia, English | Four domains and 15 subdomains |
| Canadian Association of Critical Care Nurses, 2017 | Canada, English | Seven domains of competency and 40 items |
| Critical Care Networks-National Nurse Leads, 2015 (Step 2) | English | Ten domains of competency and 27 subdomains |
| Critical Care Networks-National Nurse Leads, 2015 (Step 3) | English | Nine domains of competency and 16 subdomains |
| European federation of Critical Care Nursing associations, 2013 | EU, English | Four domains of competency and 14 subdomains |
| Japanese Society of Intensive Care Medicine, | Japan, Japanese | Three domains of competency and 36 subdomains |
| Japanese Society of Intensive Care Medicine, 2019 | Japan, Japanese | Four domains of competency and five steps of ladder |
