## Supplementary material for "Clinical practice competencies for standard critical care nursing: Consensus statement based on a systematic review and Delphi survey": S1 text

**eMethods 1. Systematic review for construction of initial competencies**

**Background and Objectives**

The education level of critical care nurses is important. Particularly, it is necessary to train nurses to provide standard critical care by themselves. This will help increase the number of nurses who can work in intensive care units (ICUs) during regular periods and will be a step toward ensuring adequate nurse availability in the event of a pandemic or other health care system crisis. There are several studies and guidelines, both in Japan and overseas, that indicate the clinical competence of ICU nurses. However, the development process of these competencies in Japan and the validity of these competencies have not been examined. Healthcare systems and abilities required in ICU nursing differ between Japan and other countries. Therefore, the characteristics of clinical practice competencies for providing standard nursing care in ICUs both nationally and internationally should be evaluated using scientific methods to develop educational guidelines and competence characteristics of ICU nurses certified by academic societies in Japan. This study aimed to develop initial set of clinical practice competencies for standard critical care nursing by systematic review (SR) ..

**Methods**

The SR was conducted according to the Preferred Reporting Items for Systematic Reviews and Meta-analyses (PRISMA) reporting guidelines. [1] eTable2 shows the supporting information for the completed PRISMA 2020 checklist. The SR protocol was not registered in PROSPERO.

**Eligible criteria**

**Design:** Any study design

**Population:** Critical care nurses

**Intervention:** Any intervention

**Outcomes:** Clinical practice skills and competencies

**Search strategy**

The following databases were searched on July 19, 2021,: MEDLINE via PubMed; CINAHL; and Igaku-Chuo-Zasshi, a Japanese medical database. The key search terms used to identify potentially relevant studies are listed in S2 Text. In addition, a non-systematic search was conducted using Google Scholar. All non-systematic searches were conducted by four authors. We attempted to identify additional relevant studies by manually searching the reference lists of both the studies returned by the search and articles citing such studies (based on Google Scholar).

**Selection of studies**

Titles and abstracts were independently screened by two of the four reviewers to identify potentially eligible studies, and their full texts were assessed for inclusion. Any concerns regarding study eligibility were deliberated by the authors. Disagreements were resolved by discussion, and if necessary, a third person was brought in for arbitration.

**Data extraction**

The data charts were created by two researchers to determine the variables that were extracted. Details of the study characteristics, including authors, publication year, title, journal, country, sample size, design, population, intervention, and outcomes, were extracted from the study. The original publications and any other additional relevant resources (searching the reference list, and tracking the author’s relevant publications) were reviewed, and information and materials were provided to identify SCCN competencies. One of the four authors independently extracted the competencies from the included articles and continued discussion until consensus was reached. We did not assess the risk of bias of the included studies.

**Results**

Of the 2310 articles identified in our search, 1920 titles and abstracts were screened for eligibility. Of these, 86 full-text articles were obtained for review, 60 articles were excluded, and 26 articles were finally included (S3 text, S1 Table, and S3 Table in the Supplementary Material). A total of 685 critical care nurse competencies were identified in the included studies. The initial list of critical care competencies was reviewed for duplication, consistency, and comprehension, and the competencies were eventually reduced to 640. These competencies were grouped into relevant categories.

1. Page, M.J., et al., *The PRISMA 2020 statement: An updated guideline for reporting systematic reviews.* PLoS Med, 2021. **18**(3): p. e1003583.
