## Supplementary material for "Clinical practice competencies for standard critical care nursing: Consensus statement based on a systematic review and Delphi survey": S2 text

**S2 Text. Search terms.**

**MEDLINE via PubMed**

#1 critical care[mh]

#3 critical care nursing[mh]

#4 #1 OR #2 OR #3

#5 "critical care"[tiab]

#6 "intensive care"[tiab]

#6 "intensive care unit"[tiab]

#7 ICU[tiab]

#8 "critical care nurse"[tiab]

#10 "critical care nursing"[tiab]

#11 #5 OR #6 OR #7 OR #8 OR #9 OR #10

#12 #4 OR #11

#13 competency[tiab]

#14 "competency framework"[tiab]

#15 "clinical ladder"[tiab]

#16 #13 OR #14 OR #15

#17 #12 AND #16

#18 animals [mh] NOT humans [mh]

#19 #16 NOT #18

**CINAHL via EBSCO**

S1 critical care

S2 intensive care unit

S3 critical care nurs*

S4 ICU

S5 S1 OR S2 OR S3 OR S4

S6 competency

S7 "competency framework"

S8 "clinical ladder"

S9 S6 OR S7 OR S8

S10 S5 AND S9

**医学中央雑誌  Igaku-Chuo-Zasshi**

#1 クリティカル

#2 クリティカルケア

#3 集中治療

#4 ICU

#5 #1 OR #2 OR #3 OR #4

#6 看護

#7 #5AND #6

#8 (専門能力/TH or コンピテンシー/AL) or (臨床能力/TH or コンピテンシー/AL)

#9 コンピテンシーフレームワーク

#10 臨床実践能力

#11 職歴の移動/TH or クリニカルラダー/AL

#12  #8 OR #9 OR #10 OR #11

#13 #7AND #11

#14 #13 AND  (PT=原著論文)
