## Supplementary material for "Clinical practice competencies for standard critical care nursing: Consensus statement based on a systematic review and Delphi survey": S3 table

**eTable 3. PRISMA 2020 flow diagram for systematic reviews**

**Identification of studies via databases and registers**

Records removed *before screening*:

Duplicate records removed (n =390 )

Records identified from*:

Pubmed (n = 646)

CHINAHL(n = 1152)

Igaku-Chuo-Zasshi (n = 489)

Handsearch (n = 23)

**Identification**

Records screened

(n = 1920)

Records excluded**

(n = 1834)

Reports sought for retrieval

(n = 86)

Reports not retrieved

(n = 0)

**Screening**

Reports assessed for eligibility

(n = 86)

Reports excluded

(n = 60)

Studies included in review

(n = 26)

**Included**

*Consider, if feasible to do so, reporting the number of records identified from each database or register searched (rather than the total number across all databases/registers).

**If automation tools were used, indicate how many records were excluded by a human and how many were excluded by automation tools.

*From:*  Page MJ, McKenzie JE, Bossuyt PM, Boutron I, Hoffmann TC, Mulrow CD, et al. The PRISMA 2020 statement: an updated guideline for reporting systematic reviews. BMJ 2021;372:n71. doi: 10.1136/bmj.n71

For more information, visit: <http://www.prisma-statement.org/>
