## Supplementary material for "Clinical practice competencies for standard critical care nursing: Consensus statement based on a systematic review and Delphi survey": S4 table

**eTable 4. Characteristics of focus group interview**

| Characteristics | n = 11 |
| --- | --- |
| Years of clinical experience     Median (IQR) | 17 (16-19.5) |
| Female                                 n (%) | 5 (45.5) |
| Facility　　　　　　　　　　　　　　　　　　　　　　 n (%) |  |
| Educational | 3 (27.3) |
| University hospital | 3 (27.3) |
| General hospital | 5 (45.5) |
| Certification n (%) |  |
| Certified Nurses | 5 (45.5) |
| Clinical Nurse Specialists | 5 (45.5) |
| Highest educational level n (%) |  |
| Master’s program | 5 (45.5) |
| Doctoral program | 1 (9.1) |
| Region in Japan n (%) |  |
| Hokkaido/Tohoku | 2 (18.2) |
| Kanto/Chubu | 5 (45.5) |
| Kinki/Chugoku/Shikoku | 2 (18.2) |
| Kyushu/Okinawa | 2 (18.2) |
