## Supplementary material for "Clinical practice competencies for standard critical care nursing: Consensus statement based on a systematic review and Delphi survey": S4 text

S4 Text. Contributors list

Miya Hamamoto, Nursing department, Tosei General Hospital, Seto, Japan.

Junko Tatsuno, Nursing department, Kokura Memorial Hospital, kitakyusyu, Japan.

Yasunobu Tsuda, Nursing department, St. Marianna University Hospital, Kawasaki, Japan.

Megumi Moriyasu, Center of Critical Care, kitasato university Hospital, Sagamihara, Japan.

Satoshi Nakata,Graduate School of Nursing Sience, St Luke’s International University, Chuo, Japan.

Sachie Nishimura, Nursing department, Okayama City Hospital, Okayama, Japan.

Ryutaro Seo. Department of Emergency Medicine, Kobe City Medical Center General Hospital, Kobe, Japan

Akihisa Okuda, Department of Clinical Engineering, Jikei University Katsushika Medical Center, Katsushika, Japan.

Etsuko Moro, Department of Nursing, Jichi Medical University Hospital, Shimotsuke, Japan.

Mio Kitayama, Nursing Department, Kanazawa Medical University Hospital, Uchinada, Japan.

Yusuke Kawai, Department of Nursing, Fujita Health University Hospital, Toyoake, Japan.

Yukiko Katayama, Nursing department, Sakakibara Heart Institute, Fuchu, Japan.

Kosuke Kitabeppu, High Care Unit, Kurashiki Central Hospital, Kurashiki, Japan.

Noriko Inagaki, Faculty of Nursing, Setsunan University, Hirakata, Japan.

Uemura Sakura, Emergency and Critical Care Medical Center, Osaka City General Hospital, Osaka, Japan.

Tomomi Furumaya, Nursing Department, Saitama Red Coss Hospital, Saitama, Japan.
