## Supplementary material for "Clinical practice competencies for standard critical care nursing: Consensus statement based on a systematic review and Delphi survey": S5 table

eTable 5. Delphi Survey Items and Results of Each Survey Item

| Disease Treatment Management and Clinical Decision Making | | | Median (IQR) | | |
| --- | --- | --- | --- | --- | --- |
| **1.1. Respiratory system** | | | 1st round | 2nd round | 3rd round |
| Anatomy and Physiology of the Respiratory System and Understanding of Disease | 1 | Understand the anatomy and physiology involved in the respiratory system | 91 (80-100) | 90 (80-100) | 92 (90-100) |
|  | 2 | Explain the four components of internal and external respiration, cellular respiration, acid-base equilibrium, and respiratory failure | 80 (70-99) | 85 (75-91) | 85 (80-96) |
|  | 3 | Explain the pathophysiology, causes, signs, symptoms, and therapeutic management of respiratory diseases such as pneumonia, asthma, chronic obstructive airway disease, acute lung injury syndrome (ALI/ARDS), pulmonary embolism, and pneumothorax | 80 (70-90) | 85 (79-90) | 85 (80-90) |
|  | 4 | Explain the effects of body position on the respiratory system | 90 (80-100) | 90 (90-100) | 90.5 (90-100) |
| Nursing Practice for Respiratory System | 5 | Perform assessment and nursing practice to improve respiratory function | 95 (80-100) | 95 (90-100) | 95 (90-100) |
|  | 6 | Understand the complications associated with suctioning airway secretions and perform suctioning using methods that minimize or prevent them | 100 (90-100) | 100 (90-100) | 100 (94-100) |
|  | 7 | Appropriately assess the risk of difficult airway clearance (cannot ventilate and cannot intubate: CVCI) and explain how to respond to it | 80 (70-95) | 85 (77-90) | 85 (80-92) |
|  | 8 | Explain the impact, benefits, and risks of each position performed as respiratory therapy | 90 (80-100) | 90 (80-100) | 90 (85-100) |
|  | 9 | Explain how to position the body to optimize respiratory function | 90 (80-100) | 90 (83-100) | 90.5 (90-98) |
|  | 10 | Report abnormalities or changes in the respiratory system to the appropriate health care professions | 100 (89-100) | 100 (93-100) | 100 (98-100) |
| Observation, monitoring and evaluation of the respiratory system | 11 | Assess the airway and secure the airway | 100 (81-100) | 100 (91-100) | 100 (95-100) |
|  | 12 | Assess for airway stenosis or obstruction | 100 (90-100) | 100 (90-100) | 100 (99-100) |
|  | 13 | Assess the amount and nature of airway secretions and sputum culture results | 80 (75-90) | 85 (75-90) | 85 (85-90) |
|  | 14 | Assess respiratory function based on respiratory frequency, breathing patterns, and use of accessory respiratory muscles | 95 (80-100) | 95 (84-100) | 95 (90-97) |
|  | 15 | Assess for diminished or absent breath sounds, left-right differences, and abnormal sounds | 100 (85-100) | 99 (90-100) | 98.5 (93-100) |
|  | 16 | Identify abnormal respiratory condition such as seesaw breathing, cyanosis, subcutaneous emphysema, uneven chest expansion, tension pneumothorax, hypoxia, restlessness, and altered mental status | 95 (80-100) | 95 (88-100) | 95 (90-99) |
|  | 17 | Use of respiratory monitors such as SpO2, SVO2, and capnography | 90 (84-100) | 90 (80-100) | 91 (90-100) |
|  | 18 | Explain the need for blood gas analysis testing | 90 (80-100) | 90 (87-100) | 90 (90-95) |
|  | 19 | Assess the results of blood gas analysis | 90 (80-91) | 90 (82-98) | 90 (85-95) |
|  | 20 | Determine the patient's condition from chest X-P, CT, and MRI (including readings) | 80 (70-90) | 80 (75-90) | 82 (80-90) |
| Administration of drugs related to the respiratory system | 21 | Understand common drugs used in the respiratory system, their indications, mechanisms of action, and potential complications | 80 (70-85) | 80 (75-82) | 80 (80-89) |
|  | 22 | Care for and manage patients requiring respiratory medicine therapy | 80 (75-90) | 80 (75-88) | 81 (80-90) |
|  | 23 | Safely prepare and manage medications related to the treatment of the respiratory system | 90 (80-100) | 92 (85-100) | 90 (86-98) |
|  | 24 | Ability to adequately monitor patients during drug administration | 100 (80-100) | 100 (90-100) | 100 (94-100) |
|  | 25 | Assess the effects of medicines related to the respiratory system and adjust care and treatment according to the patient's condition | 80 (75-90) | 80 (80-90) | 85 (80-94) |
|  | 26 | Manage airway and breathing during procedures requiring sedation | 100 (85-100) | 100 (90-100) | 100 (93-100) |
|  | 27 | Adjust drug dosage based on physician's orders to achieve established goals such as sedation scale | 90 (82-100) | 90 (85-100) | 91 (90-98) |
|  | 28 | Practice care of patients with continuous inhalation of nitric oxide | 70 (60-90) | 77 (70-85) | 80 (70-85) |
|  | 29 | Prepare, perform, and change circuits of nebulizers | 90 (80-100) | 90 (80-100) | 90 (85-100) |
| **1.2.** Cardiovascular system | | |  |  |  |
| Anatomy and Physiology of the Cardiovascular System and Understanding of Disease | 30 | Understand the anatomy and physiology related to the cardiovascular system | 100 (86-100) | 100 (95-100) | 100 (95-100) |
|  | 31 | Explain the pathophysiology, causes, signs, symptoms, and therapeutic management of cardiovascular diseases such as hypertension, peripheral vascular disease, unstable angina, acute myocardial infarction, cardiomyopathy and inflammation, heart failure, pulmonary embolism, cardiac tamponade, arrhythmia, and pacing failure | 85 (80-100) | 87 (80-97) | 89 (85-92) |
|  | 32 | Understand the normal cardiac cycle flow. | 80 (80-100) | 87 (80-99) | 90 (85-99) |
|  | 33 | Understand normal cardiac conduction. | 95 (80-100) | 95 (90-100) | 95 (92-100) |
|  | 34 | Understand the determinants of cardiac output | 90 (80-100) | 92 (90-100) | 95 (90-100) |
|  | 35 | Understand the determinants of blood pressure | 95 (80-100) | 95 (90-100) | 95 (90-100) |
|  | 36 | Understand the determinants of central venous pressure | 82 (80-90) | 85 (80-90) | 89 (80-91) |
|  | 37 | Understand the effects of ventilation and intrathoracic pressure on the cardiovascular system | 90 (80-100) | 90 (90-99) | 90 (87-93) |
| Nursing Practice for the Cardiovascular System | 38 | Manage critically ill patients with rapidly deteriorating circulatory status | 95 (89-100) | 95 (89-100) | 92.5 (90-100) |
|  | 39 | Manage patients after vascular or cardiac surgery | 90 (80-100) | 90 (80-100) | 90 (90-95) |
|  | 40 | Coordinate patient activities and nursing care taking into account circulatory load | 95 (85-100) | 95 (90-100) | 95 (90-95) |
|  | 41 | Explain indications and timing of defibrillation | 100 (80-100) | 100 (99-100) | 100 (96-100) |
|  | 42 | Explain indications and timing of pacing | 90 (76-100) | 90 (87-100) | 90 (90-95) |
| Observation, monitoring and evaluation of the cardiovascular system | 43 | Appropriate hemodynamic monitoring of adult critically ill patients | 94 (80-100) | 95 (85-100) | 95 (90-100) |
|  | 44 | Assess the arterial pressure waveforms | 90 (80-100) | 90 (85-99) | 90 (87-95) |
|  | 45 | Assess the central venous pressure values and waveforms | 80 (70-93) | 83 (80-92) | 85 (80-90) |
|  | 46 | Assess the Swan-Gans catheter readings and waveforms | 80 (71-90) | 82 (80-90) | 83 (80-90) |
|  | 47 | Assess the dynamic indicators such as SVV | 81 (75-90) | 85 (80-90) | 85 (83-90) |
|  | 48 | Assess the capillary reflex | 80 (70-95) | 83 (80-90) | 85 (80-90) |
|  | 49 | Assess the limb and skin temperatures | 95 (80-100) | 95 (90-100) | 95 (90-100) |
|  | 50 | Assess blood test results related to circulatory function | 80 (80-92) | 87 (80-92) | 90 (80-90) |
|  | 51 | Understand normal ECG waveforms | 100 (97-100) | 100 (100-100) | 100 (100-100) |
|  | 52 | Assess the common arrhythmias (atrial tachycardia, ventricular tachycardia, atrial fibrillation, ventricular fibrillation, atrioventricular block, etc.) | 100 (86-100) | 100 (98-100) | 100 (95-100) |
|  | 53 | Appropriate continuous ECG monitoring | 100 (95-100) | 100 (100-100) | 100 (99-100) |
|  | 54 | Correctly measure 12-lead ECG | 100 (95-100) | 100 (100-100) | 100 (99-100) |
|  | 55 | Observations on settings, modes, and readings of advanced circulatory support devices (e.g., VA-ECMO) and circulatory assist devices (e.g., IABP) | 80 (75-100) | 88 (80-90) | 90 (85-90) |
|  | 56 | Report abnormalities or changes in the cardiovascular system to the appropriate health care provider | 100 (95-100) | 100 (99-100) | 100 (94-100) |
| Administration of drugs related to the circulatory system | 57 | Manage critically ill patients with rapidly deteriorating circulatory status with a medical team | 92 (90-100) | 90 (87-97) | 90 (90-95) |
|  | 58 | Explain the indications, contraindications, mechanism of action, and side effects of cardiovascular agents (vasoactive medicines, inotropic medicines, antiarrhythmic medicines, etc.) | 90 (80-100) | 90 (85-95) | 90 (90-95) |
|  | 59 | Explain infusion therapy (understand the need for infusions, follow guidelines, and accurately record fluid balance) | 85 (80-90) | 90 (80-91) | 90 (88-90) |
|  | 60 | Understand the main differences between colloids, crystalloid solutions, and blood products | 80 (73-90) | 83 (80-90) | 85 (80-90) |
|  | 61 | Explain indications, contraindications, mechanism of action, and side effects of blood products | 90 (80-100) | 90 (90-96) | 90 (90-93) |
|  | 62 | Safely administer blood products and follow the facility's policy | 100 (90-100) | 100 (94-100) | 100 (92-100) |
|  | 63 | Assess clinical findings and adjust increase/decrease of circulatory agonists under the direction of a physician | 90 (80-100) | 90 (90-98) | 90 (88-98) |
|  | 64 | Tailor infusion management to the patient's physiological status | 80 (70-90) | 90 (80-95) | 90 (85-94) |
|  | 65 | Assess the effectiveness of drugs related to the cardiovascular system and adjust care and treatment according to the patient's condition | 85 (76-100) | 89 (80-91) | 90 (85-91) |
| Shock Management | 66 | Explain the classification and treatment of shock (cardiogenic, abnormal blood distribution, obstructive, hypovolemic) | 91 (90-100) | 93 (90-99) | 91 (90-97) |
|  | 67 | Understand treatment protocols according to different shock classifications and be able to assist in treatment | 90 (80-100) | 90 (88-95) | 90 (90-97) |
|  | 68 | Understand oxygenated and non-oxygenated blood flow | 90 (80-100) | 93 (90-100) | 92.5 (90-100) |
|  | 69 | Implement practices that provide emotional reassurance and support to shock patients | 99 (85-100) | 100 (95-100) | 100 (95-100) |
|  | 70 | Perform necessary assessment and monitoring for shock patients (ECG, blood pressure, temperature, urine output, IV fluids, skin, limb temperature, blood tests) | 97 (90-100) | 99 (90-100) | 100 (90-100) |
|  | 71 | Recognize and correct electrolyte, glucose, and acid-base disturbances in shock patients | 80 (70-90) | 80 (80-90) | 89.5 (80-94) |
| **1.3. Gastrointestinal system and nutrition** | | |  |  |  |
| Anatomy and physiology of the gastrointestinal system and understanding of diseases | 72 | Understand the anatomy and physiology of the gastrointestinal system (including the gastrointestinal tract, pancreas, gallbladder, and liver) | 85 (80-95) | 85 (80-94) | 85 (80-90) |
|  | 73 | Understand the endocrine and exocrine systems of the pancreas | 80 (70-90) | 80 (80-85) | 81.5 (80-88) |
|  | 74 | Understand the action of liver enzymes and understand abnormalities | 80 (72-92) | 80 (76-85) | 80 (80-85) |
|  | 75 | Understand the causes of lactic acid abnormalities | 90 (80-97) | 90 (85-97) | 90 (85-95) |
|  | 76 | Explain the pathophysiology, causes, signs, symptoms, and therapeutic management of gastrointestinal disorders such as gastrointestinal bleeding/ischemia/perforation, bowel obstruction, esophageal varices, pancreatitis, cirrhosis/liver failure, and abdominal compartment syndrome | 85 (75-90) | 85 (80-90) | 85 (80-90) |
|  | 77 | Understand the process of critical illness from physiologic changes associated with chronic and acute liver and biliary tract disorders | 80 (75-90) | 80 (80-85) | 81.5 (80-87) |
|  | 78 | Understand and assess symptoms of shock and deterioration of general condition due to gastrointestinal disease | 90 (80-95) | 90 (80-91) | 90 (85-95) |
|  | 79 | Understanding of Bacterial Translocation | 90 (80-100) | 90 (85-100) | 90 (88-95) |
|  | 80 | Understand the effects associated with increased intra-abdominal pressure | 85 (80-97) | 90 (85-96) | 90 (86-92) |
| Nursing Practice for the Gastrointestinal System | 81 | Manage patients with gastrointestinal diseases such as liver failure and shock due to gastrointestinal diseases | 80 (80-95) | 85 (80-92) | 85.5 (85-91) |
|  | 82 | Manage patients after typical abdominal surgeries such as Hartmann, esophagectomy, and bowel resection | 85 (80-100) | 85 (80-90) | 86 (85-90) |
|  | 83 | Manage patients with increased intra-abdominal pressure | 90 (80-92) | 90 (85-95) | 90 (85-90) |
|  | 84 | Manage drains associated with abdominal disease | 90 (80-100) | 90 (88-99) | 90 (90-98) |
|  | 85 | Insert a gastric tube in a critically ill patient | 50 (5-79) |  | |
|  | 86 | Manage gastric tubes in critically ill patients | 100 (90-100) | 100 (90-100) | 100 (91-100) |
|  | 87 | Manage the rectal balloon catheter for severe diarrhea | 90 (74-100) | 90 (85-96) | 90 (86-94) |
| Observation, monitoring and assessment of the gastrointestinal system | 88 | Determine the need for monitoring patients at risk for deterioration related to gastrointestinal tract function | 90 (80-100) | 90 (85-96) | 90 (89-92) |
|  | 89 | Assess diarrhea, constipation, etc. using appropriate scales | 90 (80-100) | 90 (87-99) | 90 (90-98) |
|  | 90 | Assess the need to measure intra-abdominal pressure and measure it appropriately | 86 (90-99) | 85 (80-90) | 87 (83-90) |
|  | 91 | Assess blood test results related to gastrointestinal function | 80 (76-90) | 84 (80-90) | 85 (80-90) |
|  | 92 | Report abnormalities or changes in the gastrointestinal system to the appropriate health care provider | 90 (90-100) | 91 (90-100) | 90 (90-100) |
| Administration of medicines related to the gastrointestinal system | 93 | Assess the effectiveness of medicines related to the gastrointestinal system and adjust care and treatment according to the patient's condition | 80 (75-90) | 80 (80-89) | 85 (80-90) |
|  | 94 | Understand medicine specificity (e.g., contraindications to grinding) | 90 (77-100) | 90 (83-97) | 90 (89-91) |
|  | 95 | Adjust dosage of medication to achieve therapeutic effect under the direction of a physician | 90 (80-100) | 90 (83-100) | 90 (90-95) |
|  | 96 | Understand intestinal motility and motility enhancers, laxatives, anti-stimulants, insulin/hypoglycemic agents, probiotics, steroids, antidiarrheals, and antisecretory agents | 81 (75-90) | 80 (80-90) | 84.5 (80-90) |
| Nursing Practice for Nutritional Management | 97 | Understand the nutritional needs of individuals (calorie requirements, protein levels, etc.) | 80 (70-90) | 80 (80-90) | 84 (80-90) |
|  | 98 | Assess changes in nutritional status | 80 (86-92) | 85 (80-90) | 85 (80-90) |
|  | 99 | Understand the need for vitamins and minerals | 80 (70-90) | 80 (80-90) | 81 (80-90) |
|  | 100 | Assess the nutrition-related laboratory values | 80 (75-90) | 80 (80-90) | 84.5 (80-90) |
|  | 101 | Explain to the patient's past medical history and describe diseases affecting digestive function | 80 (70-90) | 80 (80-88) | 83.5 (80-90) |
|  | 102 | Adjust nutritional strategies according to guidelines and other recommendations | 80 (70-80) | 80 (79-90) | 80.5 (80-86) |
|  | 103 | Assess the results of nutritional therapy and adjust nutritional therapy according to the patient's condition | 78 (60-80) | 87 (79-97) | 90 (85-93) |
| Nursing Practice for Dysphagia | 104 | Explain the mechanism of swallowing | 80 (80-100) | 81 (80-90) | 85 (80-90) |
|  | 105 | Explain risk factors for dysphagia | 90 (80-100) | 90 (80-96) | 90 (85-90) |
|  | 106 | Assess swallowing function (water-only test, repeated saliva-only test, food test, etc.) | 90 (76-100) | 90 (85-97) | 90 (83-95) |
|  | 107 | Provide consultation to specialists regarding the causes of dysphagia and therapeutic rehabilitation methods. | 80 (72-90) | 82 (80-90) | 85 (80-90) |
|  | 108 | Prepare the environment for eating, adjust food form and thickening, and provide appropriate dietary assistance. | 90 (80-100) | 90 (85-100) | 95 (90-100) |
| **1.4. Renal system** | | |  |  |  |
| Anatomy and Physiology of the Renal and Urinary System and Understanding of Disease | 109 | Understand the anatomy and physiology of the renal and urinary system | 80 (70-100) | 80 (72-91) | 83.5 (76.5-90) |
|  | 110 | Understand the function of the kidneys | 87 (80-100) | 90 (80-100) | 90 (80-95) |
|  | 111 | Understand electrolyte excretion | 80 (70-100) | 83 (76-100) | 88.5 (88-98) |
|  | 112 | Understand the renal blood supply | 75 (70-89) | 80 (75-100) | 85 (79-100) |
|  | 113 | Explain electrolyte abnormalities | 90 (80-100) | 90 (80-100) | 93 (84-100) |
|  | 114 | Explain the pathophysiology, causes, signs, symptoms, and therapeutic management of renal and urologic diseases, including renal disorders (acute kidney injury, chronic kidney injury, end-stage renal disease) and electrolyte abnormalities | 85 (80-100) | 80 (74-95) | 84 (73.5-91.5) |
| Nursing Practice for Renal and Urinary System | 115 | Insert a urinary catheter | 100 (83-100) | 100 (100-100) | 100 (100-100) |
|  | 116 | Manage a urinary catheter after placement | 100 (100-100) | 100 (100-100) | 100 (100-100) |
|  | 117 | Understand guideline-based treatment strategies for acute kidney injury | 80 (66-90) | 80 (70-90) | 80 (75-90) |
|  | 118 | Understand the selection and principles of renal replacement therapy in HD, CHDF, CVVHD, CVVH, SLED and peritoneal dialysis | 80 (70-98) | 80 (71-90) | 80 (77-92) |
|  | 119 | Develop an individualized plan of care for renal replacement therapy | 75 (60-80) | 77 (60-90) | 79 (71-84) |
| Observation, monitoring and evaluation of the renal and urinary system | 120 | Determine the need for monitoring of people at risk of renal function decline | 80 (80-100) | 80 (80-97) | 86 (80-99) |
|  | 121 | Understand how to monitor fluid status, fluid delivery, and renal function in patients at risk for renal impairment (cardiovascular, dehydration, overflow, balance check, body weight, ml/kg, Cr, nephrotoxic drugs, drug dosage adjustment in renal failure, fluid overload, hyperkalemia) | 90 (80-100) | 90 (80-100) | 90 (82.5-98) |
|  | 122 | Understand how to measure and record fluid volume | 95 (80-100) | 96 (80-100) | 100 (90-100) |
|  | 123 | Understand the causes of fluid loss (drains, gastrointestinal system, hemorrhage, insensate excretion, etc.) | 95 (80-100) | 90 (85-100) | 99.5 (89-100) |
|  | 124 | Assess blood collection results in patients with renal and urinary dysfunction | 87 (80-100) | 88 (80-95) | 90 (82.5-98.5) |
|  | 125 | Appropriately report abnormalities or changes in the renal and urinary system | 90 (81-100) | 95 (86-100) | 92.5 (90-100) |
| Administration of drugs related to the renal and urinary system | 126 | Understand diuretics, glucose and insulin, salbutamol (nebulizer), calcium, sodium bicarbonate, and other medicines related to the kidney | 80 (75-94) | 80 (79-93) | 80 (76-95.5) |
|  | 127 | Assess the effectiveness of infusions and medications related to the renal and urologic system and adjust care and treatment according to the patient's condition | 80 (70-90) | 80 (75-90) | 82 (80-94.5) |
| **1.5. Endocrine and metabolic systems** | | |  |  |  |
| Anatomy and physiology of the endocrine and metabolic systems and understanding of disease | 128 | Understand the anatomy and physiology of the endocrine and metabolic systems | 76 (65-90) | 80 (70-90) | 80 (70.5-90) |
|  | 129 | Explain the pathophysiology, causes, signs, symptoms, and therapeutic management of metabolic and endocrine disorders such as diabetes, adrenal insufficiency, and thyroid | 75 (60-90) | 80 (70-90) | 80 (75-90) |
| Nursing Practice for Endocrine and Metabolic Systems | 130 | Explain the need for blood glucose monitoring in critically ill patients | 95 (81-100) | 96 (89-100) | 100 (90-100) |
|  | 131 | Explain the care and management of patients with diabetic ketoacidosis, nonketotic hyperosmotic syndrome, and hypoglycemia | 84 (76-100) | 85 (75-94) | 85.5 (81-95) |
|  | 132 | Explain the care and management of patients with abnormal thyroid function (hyperthyroidism including crisis, hypothyroidism) | 80 (60-90) | 80 (70-90) | 80 (74.5-90) |
| Observation, monitoring and evaluation of endocrine and metabolic systems | 133 | Explain the need for blood glucose monitoring in critically ill patients | 90 (80-100) | 95 (90-100) | 100 (90-100) |
|  | 134 | Assess periodic blood glucose readings based on patient condition and the effects of medications used | 90 (80-100) | 90 (85-100) | 90 (86.5-100) |
|  | 135 | Make necessary observations and assess patients with abnormal thyroid function (hyperthyroidism including crisis, hypothyroidism) appropriately | 80 (70-90) | 80 (70-90) | 80 (75.5-91.5) |
|  | 136 | Assess the results of blood tests related to endocrine and metabolic functions | 80 (70-93) | 80 (70-95) | 80 (78-90) |
|  | 137 | Report endocrine and metabolic abnormalities and changes to appropriate health care providers | 86 (70-100) | 90 (80-100) | 90 (83-98) |
| Management of drugs related to endocrine and metabolic systems | 138 | Understand insulin and other medicines related to the endocrine and metabolic systems | 80 (75-100) | 85 (80-98) | 90 (80.5-98.5) |
|  | 139 | Assess the effects of medicines related to the endocrine and metabolic systems and adjust care and treatment according to the patient's condition | 80 (68-90) | 80 (75-90) | 82.5 (80-90) |
|  | 140 | Manage insulin administration based on blood glucose levels | 95 (85-100) | 99 (90-100) | 100 (93-100) |
| **1.6. Cerebral nervous system** | | |  |  |  |
| Anatomy and physiology of the nervous system and understanding of disease | 141 | Understand the anatomy and physiology of the cranial nervous system | 90 (80-100) | 90 (80-100) | 91.5 (82.5-97.5) |
|  | 142 | Explain the Monroe Kelly's Law | 75 (50-88) | 72 (60-80) | 76 (67.5-80) |
|  | 143 | Explain the Cushing's phenomenon | 90 (80-100) | 90 (80-100) | 90 (80.5-100) |
|  | 144 | Understand primary and secondary brain injury | 81 (70-100) | 85 (80-100) | 88.5 (82.5-100) |
|  | 145 | Be able to determine how neurological impairment may compromise patient safety | 86 (80-100) | 85 (80-100) | 90 (82.5-99) |
|  | 146 | Explain the pathophysiology, causes, signs, symptoms, and therapeutic management of neurological disorders such as dementia, cerebral hemorrhage, cerebral infarction, subarachnoid hemorrhage, seizures, and encephalitis and meningitis | 90 (80-100) | 90 (80-100) | 90 (81.5-96) |
| Nursing Practice for the Brain Nervous System | 147 | Provide nursing care for patients with neurological dysfunctions | 98 (80-100) | 95 (88-100) | 95 (90-100) |
|  | 148 | Understand the symptoms associated with seizures and manage and care for them with safety in mind | 90 (80-100) | 90 (86-100) | 92 (89-100) |
|  | 149 | Manage the airway in relation to impaired consciousness | 100 (90-100) | 100 (95-100) | 100 (92.5-100) |
|  | 150 | Adjust body position with an understanding of changes in cerebral pressure and cerebral blood flow | 95 (80-100) | 96 (85-100) | 96 (88.5-100) |
|  | 151 | Know the indications for CT and MRI of the head | 80 (60-100) | 85 (71-100) | 85 (80-96) |
|  | 152 | Provide care that is aware of the potential impact on ICP | 90 (80-100) | 90 (80-100) | 90 (86-100) |
|  | 153 | Devise and implement appropriate strategies to maintain adequate cerebral perfusion pressure or MAP when ICP cannot be monitored | 80 (80-97) | 90 (80-100) | 90 (85-99) |
|  | 154 | Assess nursing care for rising ICP and adjust care plans accordingly | 90 (80-100) | 90 (80-100) | 90 (86.5-99) |
|  | 155 | Explain the criteria for transport to a specialized hospital for neurological diseases | 70 (40-80) |  | |
| Observation, monitoring and evaluation of the brain nervous system | 156 | Assess and accurately record Glasgow Coma Scale (GCS) | 100 (100-100) | 100 (99-100) | 100 (100-100) |
|  | 157 | Observe and assess pupils (size, shape, responsiveness) | 100 (100-100) | 100 (100-100) | 100 (100-100) |
|  | 158 | Assess cranial nervous system function such as paralysis of the extremities | 100 (90-100) | 100 (90-100) | 100 (99-100) |
|  | 159 | Understand the mechanisms of normal control of cerebral perfusion and intracranial pressure (ICP) and explain normal parameters of intracranial pressure (ICP) and cerebral perfusion pressure (CPP) | 80 (80-100) | 82 (80-93) | 85 (80-92.5) |
|  | 160 | Recognize signs and symptoms of elevated intracranial pressure (ICP) | 90 (80-100) | 90 (80-100) | 90.5 (88.5-99.5) |
|  | 161 | Monitor hemodynamics with consideration of the impact on cranial nerve damage | 90 (80-100) | 90 (80-100) | 91 (86.5-100) |
|  | 162 | Ability to adequately monitor patients with brain nervous system disorders during hypothermia therapy. | 87 (80-100) | 90 (83-100) | 91 (86.5-100) |
|  | 163 | Recognize abnormalities in continuous EEG monitoring for seizure patients and others. | 70 (50-80) | 75 (70-94) | 80 (70-89.5) |
|  | 164 | Appropriately report abnormalities or changes in the cranial nervous system | 95 (85-100) | 100 (90-100) | 100 (92.5-100) |
| Administration of drugs related to the nervous system | 165 | Prepare and administer therapeutic agents (osmotic treatments, analgesics, muscle relaxants, anticonvulsants, catecholamines, steroids, antihypertensive medications, etc.) for the treatment of cranial nervous system disorders | 89 (80-100) | 90 (81-100) | 90 (86-96.5) |
|  | 166 | Assess fluid balance with consideration of the impact on cranial nerve damage and consult on the administration of appropriate fluids | 85 (79-100) | 85 (80-100) | 87.5 (85-95) |
|  | 167 | Assess the effects of medicines related to the nervous system, consult with patients on their conditions, and coordinate their care | 85 (80-99) | 85 (80-100) | 87.5 (85-95) |
| **1.7. Skin/musculoskeletal system** | | |  |  |  |
| Anatomy and physiology of the skin, musculoskeletal system, and understanding of disease | 168 | Understand the anatomy and physiology of the skin, musculoskeletal system | 80 (70-90) | 80 (72-92) | 81.5 (80-90) |
|  | 169 | Explain ICU-AW | 85 (75-100) | 85 (80-100) | 86 (80-100) |
|  | 170 | Explain risk factors for ICU-AW | 85 (77-100) | 85 (80-100) | 87 (80-94.5) |
|  | 171 | Explain risk factors for pressure ulcers, pressure wounds on medical devices, and skintightness in critically ill patients | 90 (80-100) | 90 (83-100) | 90 (85-100) |
| Nursing practice for skin, musculoskeletal system | 172 | Explain how to prevent and treat ICU-AW | 85 (71-100) | 85 (80-100) | 87 (80-96.5) |
|  | 173 | Prepare, treat, and care for wounds accordingly | 85 (80-100) | 90 (82-99) | 90 (85-100) |
|  | 174 | Explain how to prevent pressure ulcers, pressure wounds on healthcare-related equipment, and skin lesions in critically ill patients. | 90 (85-100) | 90 (85-100) | 92 (88.5-100) |
|  | 175 | Explain the care of critically ill patients with pressure ulcers, pressure wounds and skin lesions on medical related equipment. | 90 (80-100) | 90 (80-100) | 90 (84.5-100) |
| Observation, monitoring and evaluation of skin, musculoskeletal system | 176 | Monitor and assess musculoskeletal system related issues such as muscle strength (MMT, MRC score, grip strength, etc.) and ADL assessment | 85 (70-99) | 90 (80-99) | 90 (85.5-96) |
|  | 177 | Observe skin and wounds (including trauma and post-operative) and detect abnormalities early | 100 (90-100) | 100 (95-100) | 100 (90-100) |
|  | 178 | Assess pressure ulcer risk (e.g., Braden scale) in critically ill patients | 90 (80-100) | 90 (85-100) | 95 (85-100) |
|  | 179 | Continuous monitoring (e.g. DESIGN-R) and evaluation of pressure ulcers in critically ill patients | 90 (80-100) | 90 (83-100) | 94.5 (86.5-100) |
|  | 180 | Appropriately report abnormalities or changes in the skin, musculoskeletal system | 90 (80-100) | 92 (85-100) | 90 (85-99) |
| Knowledge of pharmacology as it relates to the skin, musculoskeletal system | 181 | Explain agents that affect the musculoskeletal system, including ICU acquired muscle weakness | 80 (70-89) | 80 (70-99) | 84.5 (79-91.5) |
|  | 182 | Explain the treatment methods (drugs) for pressure ulcers, pressure wounds on medical-related equipment, and skintare in critically ill patients | 85 (75-99) | 85 (80-100) | 85.5 (81-95.5) |
|  | 183 | Assess the effects of medicines related to the skin, musculoskeletal system, and adjust care and treatment according to the patient's condition | 81 (70-95) | 95 (80-100) | 90 (80-95) |
| **1.8. Infectious diseases, blood and immune system** | | |  |  |  |
| Anatomy and physiology and disease understanding of infectious diseases, blood and immune systems | 184 | Understand the anatomy and physiology of the blood and immune system | 80 (65-96) | 80 (70-93) | 84 (76-92) |
|  | 185 | Explain the pathophysiology, causes, signs, symptoms, and treatment management of sepsis | 95 (80-100) | 97 (85-100) | 98 (87.5-100) |
|  | 186 | Understand the definition and diagnostic criteria for sepsis | 90 (80-100) | 91 (80-100) | 92.5 (85-100) |
|  | 187 | Understand the septic shock | 99 (85-100) | 100 (90-100) | 100 (94-100) |
| Nursing Practice for Infectious Diseases, Hematology, and Immune System | 188 | Recognize patients with infectious diseases who are at risk of worsening their condition | 85 (80-100) | 88 (80-95) | 90 (83.5-99) |
|  | 189 | Refer to sepsis guidelines and practice appropriate care bundles | 90 (80-100) | 90 (80-100) | 93 (90-100) |
|  | 190 | Take necessary infection control precautions for individual patients and treatment environments | 99 (85-100) | 100 (90-100) | 100 (96-100) |
|  | 191 | Take precautions against multimedicine-resistant bacteria | 95 (85-100) | 100 (90-100) | 100 (94.5-100) |
| Prevention and Nursing Care of Device-Related Infections | 192 | Explain risk factors for catheter-related bloodstream infections | 90 (90-100) | 94 (86-100) | 95 (90-100) |
|  | 193 | Explain how to prevent and treat catheter-related bloodstream infections | 90 (80-100) | 93 (85-100) | 94 (90-100) |
|  | 194 | Explain risk factors for ventilator-associated pneumonia and ventilator-associated events (VAE) | 91 (80-100) | 95 (86-100) | 95 (91-100) |
|  | 195 | Explain how to prevent and treat ventilator-associated pneumonia and ventilator-associated events (VAE) | 93 (80-100) | 96 (88-100) | 95 (90-100) |
|  | 196 | Explain how to prevent and treat urinary catheter-related urinary tract infections | 95 (80-100) | 95 (82-100) | 95.5 (90-100) |
|  | 197 | Explain how to prevent and treat urinary catheter-related urinary tract infections | 91 (80-100) | 95 (84-100) | 95 (86.5-100) |
|  | 198 | Explain risk factors for surgical site infection | 90 (78-100) | 90 (86-100) | 92 (86.5-100) |
|  | 199 | Explain how to prevent and treat surgical site infections | 87 (80-100) | 90 (80-100) | 90 (88-100) |
| Nursing Practice for Blood Coagulation Abnormality | 200 | Practice nursing care for patients with blood coagulation abnormalities | 90 (75-100) | 90 (80-100) | 91 (85-100) |
|  | 201 | Understand hematological disorders such as major bleeding requiring massive blood transfusions, immunosuppression, and immunodeficiency | 85 (70-100) | 88 (80-100) | 90 (81-93) |
|  | 202 | Explain the transfusion-related adverse events such as transfusion-related acute lung injury (TRALI), Graft versus host disease (GVHD), etc. | 80 (70-91) | 81 (78-100) | 83.5 (77-95) |
|  | 203 | Understand disseminated intravascular coagulation syndrome (DIC) | 90 (72-100) | 92 (81-100) | 90 (89.5-100) |
|  | 204 | Understand the indications and administration of anticoagulants | 88 (75-100) | 90 (80-100) | 90 (82-100) |
|  | 205 | Explain risk factors for deep vein thrombosis (DVT) | 95 (80-100) | 99 (80-100) | 99 (88.5-100) |
|  | 206 | Explain how to prevent and treat deep vein thrombosis (DVT) | 95 (80-100) | 97 (82-100) | 96.5 (89-100) |
| **1.9. Other diseases** | | |  |  |  |
| Nursing Practice for Resuscitation and Sudden Changes | 207 | Anticipate rapidly changing patient conditions and respond proactively | 100 (90-100) | 100 (94-100) | 100 (88-100) |
|  | 208 | Assess and respond to rapidly changing patient and treatment situations | 95 (85-100) | 95 (90-100) | 96 (90-100) |
|  | 209 | Recognize early warning signs quickly and consult with other nursing staff, medical team members, and others as needed | 90 (80-100) | 100 (95-100) | 100 (94.5-100) |
|  | 210 | Consider care and clinical priorities in response to emergencies and unforeseen circumstances | 95 (85-100) | 98 (90-100) | 98 (90-100) |
|  | 211 | Recognize and assess critically ill patients who deteriorate rapidly and manage them toward condition stabilization | 92 (80-100) | 91 (89-100) | 94.5 (90-100) |
|  | 212 | Respond to cardiopulmonary arrest (BLS, ACLS) | 100 (100-100) | 100 (100-100) | 100 (100-100) |
|  | 213 | Defibrillate according to resuscitation protocols | 98 (89-100) | 100 (90-100) | 100 (93-100) |
|  | 214 | Identify and respond to lethal arrhythmias | 100 (100-100) | 100 (100-100) | 100 (100-100) |
|  | 215 | Manage post-resuscitation, including management of airway, breathing, circulation, arrhythmias, and abnormal metabolic states | 99 (80-100) | 99 (80-100) | 97 (90-100) |
| Nursing Practice for Trauma | 216 | Explain the pathophysiology, causes, signs, symptoms, and treatment management of multiple trauma, head trauma, thoracoabdominal trauma, extremity pelvic trauma, and spinal trauma | 80 (65-90) | 80 (70-85) | 80.5 (77.5-89) |
|  | 217 | Provide nursing care for trauma patients | 70 (50-80) | 60 (50-70) |  |
| Nursing Practice for Burns | 218 | Explain the pathophysiology, causes, signs, symptoms, and management of burns | 80 (60-90) | 80 (70-85) | 80 (70-87.5) |
|  | 219 | Provide nursing care for burn patients | 80 (70-90) | 80 (70-94) | 80 (75-89) |
| Nursing Practice for Acute Poisoning | 220 | Explain the pathophysiology, causes, signs, symptoms, and treatment management of acute poisoning, including acute drug poisoning, carbon monoxide poisoning, and natural poison poisoning | 75 (60-80) | 71.5 (65-80) | 74 (69-80) |
|  | 221 | Provide nursing care for acutely poisoned patients, including acute drug poisoning, carbon monoxide poisoning, and natural poison poisoning | 80 (56-90) | 75.5 (70-85) | 75 (70-80) |
| Nursing practice for abnormal body temperature | 222 | Explain the pathophysiology, causes, signs, symptoms, and treatment management of hyperthermia and hypothermia, including heat stroke, malignant hyperthermia, and malignant syndrome | 80 (65-90) | 80 (70-87) | 80 (71.5-85) |
|  | 223 | Provide nursing care for patients with hyperthermia and hypothermia, including heat stroke, malignant hyperthermia, and malignant syndromes | 80 (70-90) | 80 (70-90) | 80 (78.5-85) |
|  | 224 | Handle, assist, observe, and manage the use of temperature control devices. | 90 (80-100) | 90 (71-100) | 85 (80-90.5) |
| Nursing Practice for Emergency Conditions of Pregnant Women | 225 | Explain the pathophysiology, causes, signs, symptoms, and treatment management of gestational hypertension, amniotic fluid embolism, cervical laceration, and flaccid hemorrhage | 50 (31-80) | 50 (45-62) |  |
|  | 226 | Provide nursing care for patients with gestational hypertension, amniotic fluid embolism, cervical laceration, and flaccid hemorrhage | 50 (40-80) | 50 (45-70) |  |
| **1.10. Treatment equipment management** | | |  |  |  |
| Noninvasive ventilatory management | 227 | Explain the indications for non-invasive ventilation | 100 (90-100) | 100 (97-100) | 100 (94-100) |
|  | 228 | Explain the advantages and disadvantages of noninvasive ventilation | 100 (90-100) | 100 (97-100) | 100 (95.5-100) |
|  | 229 | Explain the physiological and psychological effects of noninvasive ventilation on patients | 100 (85-100) | 100 (100-100) | 100 (96-100) |
|  | 230 | Correctly prepare non-invasive ventilators (including circuit assembly, proper parameter and alarm settings) | 86 (78-100) | 99 (80-100) | 95 (84-100) |
|  | 231 | Use of a heated humidifier for non-invasive ventilators | 100 (90-100) | 100 (95-100) | 100 (90-100) |
|  | 232 | Provide initiate, administer, and wean from noninvasive ventilation | 92 (84-100) | 96 (85-100) | 99 (87-100) |
|  | 233 | Explain the different types of masks and mask fitting methods | 100 (90-100) | 100 (90-100) | 100 (95-100) |
|  | 234 | Prevent complications of non-invasive ventilation (e.g., skin problems) | 100 (90-100) | 100 (90-100) | 100 (95-100) |
|  | 235 | Assist in the daily living of patients on non-invasive ventilation while dealing with their symptoms | 100 (89-100) | 100 (95-100) | 100 (93.5-100) |
|  | 236 | （Adjust sedation and other treatments and ventilator settings according to the condition of the patient being non-invasively ventilated (based on physician orders) | 90 (80-100) | 92.5 (80-100) | 90 (80-100) |
|  | 237 | Correctly troubleshoot non-invasive ventilatory equipment | 98 (85-100) | 98 (85-100) | 99 (85-100) |
| Invasive ventilatory management | 238 | Explain the indications for invasive ventilation | 100 (90-100) | 100 (90-100) | 100 (90-100) |
|  | 239 | Explain the physiological and psychological effects of invasive mechanical ventilation on patients | 100 (90-100) | 100 (90-100) | 100 (92.5-100) |
|  | 240 | Invasive mechanical ventilator is properly prepared (including circuit assembly, proper parameter and alarm settings) | 90 (80-100) | 94 (85-100) | 92.5 (84-100) |
|  | 241 | Use of a Heated humidifier for invasive mechanical ventilators | 100 (90-100) | 100 (95-100) | 100 (91-100) |
|  | 242 | Initiate, manage, and wean from invasive mechanical ventilation | 94 (80-100) | 100 (85-100) | 100 (86-100) |
|  | 243 | Assist with daily living while dealing with symptoms of patients on invasive ventilation | 100 (90-100) | 100 (97-100) | 100 (92-100) |
|  | 244 | Identify ventilation modes and ventilator settings explain them | 94 (80-100) | 98.5 (80-100) | 100 (89-100) |
|  | 245 | Explain how to prevent complications of invasive mechanical ventilation (e.g., lung protective ventilation for ventilator lung injury) | 90 (80-100) | 95 (80-100) | 95 (90-100) |
|  | 246 | （Adjust treatment and mechanical ventilator settings, such as sedation, according to the patient's condition during invasive ventilation (based on physician's orders) | 90 (82-100) | 90.5 (80-100) | 94 (84.5-100) |
|  | 247 | Troubleshoot invasive mechanical ventilator (high pressure, low pressure, low tidal volume, high peak airway pressure, high breathing and other alarms, power failure response, equipment malfunction, etc.) | 91 (80-100) | 94.5 (80-100) | 95.5 (82.5-100) |
|  | 248 | Explain the significance of practicing the invasive mechanical ventilator Care Bundle | 90 (80-100) | 92 (80-100) | 95 (82-100) |
|  | 249 | Coordinate and implement personnel needed for prone therapy during invasive mechanical ventilation | 86 (80-100) | 89 (79-100) | 90 (81.5-100) |
|  | 250 | Assist with bronchoscopy during invasive mechanical ventilation | 90 (70-100) | 90 (76-100) | 90 (80-100) |
| Nursing Practices Related to Oral Intubation | 251 | Explain the indications, advantages, and disadvantages of intubation | 100 (90-100) | 100 (96-100) | 100 (93.5-100) |
|  | 252 | Understand the intubation process and prepare necessary supplies and medications | 100 (95-100) | 100 (95-100) | 100 (92.5-100) |
|  | 253 | Assist with intubation and extubation | 100 (95-100) | 100 (100-100) | 100 (100-100) |
|  | 254 | Appropriately manage intubation tube (including position, fixation, and cuff pressure management) | 100 (95-100) | 100 (99-100) | 100 (90.5-100) |
|  | 255 | Assess the patency of the artificial airway and assess and respond to urgent airway problems (airway obstruction due to sputum, unscheduled tracheal tube removal or dislodgement, pneumothorax). | 90 (80-100) | 92 (85-100) | 95 (89.5-100) |
|  | 256 | Assess the need for suctioning during ventilation using indicators such as cough, secretions, hypoxemia, restlessness, high airway pressure, and hemodynamic changes | 100 (90-100) | 100 (90-100) | 99.5 (90-100) |
|  | 257 | Explain the advantages and disadvantages of subglottic suction | 90 (80-100) | 96.5 (85-100) | 99 (89.5-100) |
|  | 258 | Practice measures to minimize the causes and risks of emergency reintubation | 90 (80-100) | 93 (80-100) | 96 (90-100) |
|  | 259 | Properly care for the oral health of intubated patients | 100 (90-100) | 100 (97-100) | 100 (92-100) |
|  | 260 | Capable of manual ventilation using a bag-valve device | 100 (85-100) | 100 (98-100) | 100 (92-100) |
| Practices related to tracheostomy | 261 | Understand percutaneous tracheostomy, surgical tracheostomy, and minitrack, their indications, advantages, and disadvantages | 90 (75-100) | 92.5 (80-100) | 92 (80-99) |
|  | 262 | Appropriate timing of tracheostomy can be identified. | 80 (62-90) | 85 (77-100) | 89.5 (80-99.5) |
|  | 263 | Explain the indications and necessity for emergency airway clearance procedures such as emergency cricothyrotomy | 80 (65-100) | 80 (70-98) | 84 (77-95) |
|  | 264 | Explain complications after tracheostomy | 100 (80-100) | 100 (84-100) | 100 (85-100) |
|  | 265 | Understand the process of performing a tracheostomy and be able to practice preparing and assisting with necessary supplies and medications | 90 (76-100) | 90.5 (80-100) | 93.5 (84.5-100) |
|  | 266 | Ability to practice patient care and observation before, during, and after tracheostomy | 90 (80-100) | 91 (80-100) | 93 (86-100) |
|  | 267 | Ability to properly manage (including position, fixation, and cuff pressure) tracheostomy tubes, including speaking valves | 90 (74-100) | 90 (75-100) | 90 (80-92.5) |
|  | 268 | Explain the advantages and disadvantages of suction above the cuff | 100 (80-100) |  | |
|  | 269 | Observe patients for possible physical and psychological effects associated with tracheostomy and respond accordingly | 90 (80-100) | 90 (80-100) | 90 (81-100) |
| Knowledge of renal replacement therapy | 270 | Explain the types of renal replacement therapy and the advantages and disadvantages of each method | 90 (75-100) | 90 (80-100) | 90 (84.5-100) |
|  | 271 | Ensure that the necessary circuits for renal replacement therapy are assembled correctly | 71 (50-100) | 78 (60-85) | 79 (70-99.5) |
|  | 272 | Explain the advantages and disadvantages of different types of dialysis solutions | 75 (50-92) | 75 (60-85) | 75 (68.5-92.5) |
|  | 273 | Select the treatment mode that best suits the individual patient and set individual predefined treatment goals and adjust treatment according to coagulation, electrolyte, and acid-base goals. | 71 (50-80) | 70 (60-87) | 80 (70-84.5) |
|  | 274 | Properly manage equipment during renal replacement therapy | 87 (70-100) | 80.5 (75-100) | 85 (80-94.5) |
|  | 275 | Explain complications during renal replacement therapy and how to minimize them (thrombocytopenia/clotting disorders, anemia, circulation, electrolytes, bleeding, hypothermia, infection, thrombosis/embolism) | 86 (70-100) | 80.5 (70-95) | 85 (78-93) |
|  | 276 | The impact of renal replacement therapy on the metabolism of medicines and how to deal with it appropriately | 80 (60-100) | 80 (70-95) | 80 (76.5-90) |
|  | 277 | Appropriate equipment monitoring (access pressure, membrane pressure, etc.) and troubleshooting during renal replacement therapy | 80 (70-100) | 80.5 (70-90) | 81.5 (75.5-93.5) |
|  | 278 | Report any abnormalities or changes during renal replacement therapy to the appropriate health care provider | 100 (80-100) | 100 (87-100) | 100 (87.5-100) |
|  | 279 | Provide psychological care for patients undergoing renal replacement therapy | 90 (80-100) | 93.5 (83-100) | 95 (82.5-100) |
|  | 280 | Accurate fluid balance including cumulative balance after renal replacement therapy to record | 91 (80-100) | 96 (84-100) | 96 (81-100) |
| Drain management | 281 | Correctly assemble evidence-based equipment required for insertion of a thoracic drain | 100 (81-100) | 100 (93-100) | 100 (90-100) |
|  | 282 | Assist with insertion and removal of chest drains | 100 (90-100) | 100 (95-100) | 100 (91-100) |
|  | 283 | Appropriately manage patients with chest drains inserted | 100 (100-100) | 100 (91-100) | 100 (90-100) |
|  | 284 | Adjust treatment modalities related to thoracic drainage according to the patient's condition | 90 (50-100) | 100 (80-100) | 100 (91.5-100) |
|  | 285 | Assist with emergency decompression of tension pneumothorax | 90 (80-100) | 90 (85-100) | 90 (80-100) |
|  | 286 | Troubleshoot problems related to various drains, including ventricular drains and abdominal drains | 99 (80-100) | 98.5 (90-100) | 100 (84.5-100) |
|  | 287 | Manage various types of drains, including ventricular drains and abdominal drains | 100 (84-100) | 100 (93-100) | 100 (87.5-100) |
|  | 288 | Provide psychological care to patients with various drain insertions, including ventricular drains and abdominal drains | 100 (82-100) | 100 (88-100) | 100 (84-100) |
| Management of circulatory assist devices (IABP, IMPELLA, etc.) | 289 | Understand the indications for circulatory assist devices (IABP, IMPELLA, etc.) | 90 (80-100) | 90 (80-98) | 91 (80-100) |
|  | 290 | Manage and care for patients on circulatory assist devices (IABP, IMPELLA, etc.) | 92 (75-100) | 90 (80-99) | 90 (80-100) |
| Management of auxiliary circulatory equipment (VA-ECMO, etc.) | 291 | Explain indications for assistive circulatory devices (e.g., VA-ECMO) | 90 (71-100) | 80 (93-100) | 91 (80-100) |
|  | 292 | Manage and care for patients on assisted circulatory devices (e.g., VA-ECMO) | 71 (90-100) | 94 (80-100) | 94 (80-100) |
| Management of pulmonary function assist devices (e.g. VV-ECMO) with membrane type artificial lungs | 293 | Explain indications for pulmonary function assist devices (e.g., VV-ECMO) using membrane-type artificial lungs | 74 (90-100) | 90 (80-100) | 90 (80-100) |
|  | 294 | Manage and care for patients using (e.g., VV-ECMO, a pulmonary function support device using membrane-type artificial lungs) | 90 (77-100) | 90.5 (80-100) | 92.5 (80-100) |
| Pacemaker Management | 295 | Explain indications and timing of defibrillation | 90 (70-100) | 90 (74-100) | 90 (80-100) |
|  | 296 | Explain indications and timing of pacing | 96 (80-100) | 95.5 (78-100) | 95 (84.5-100) |
| Various line management | 297 | Assist, observe, and handle the use of an implantable pacemaker | 100 (95-100) | 100 (98-100) | 100 (90-100) |
|  | 298 | Assist, observe, and respond to the use of external (temporary) pacing | 100 (95-100) | 100 (99-100) | 100 (90.5-100) |
| **1.11. organ transplant** | | |  |  |  |
| Indications for organ transplantation and donor identification | 299 | Understand the anatomy and physiology of the brainstem | 91 (80-100) | 90 (80-100) | 90 (80.5-100) |
|  | 300 | Understand the causes of irreversible disorders of consciousness | 90 (80-100) | 90 (79-100) | 90 (83-100) |
|  | 301 | Know the procedures for early identification of patients indicated for organ transplantation. | 70 (60-86) | 68 (51-80) |  |
|  | 302 | Identify donors and discuss with the medical team the possibility of tissue or organ donation | 68 (50-80) | 69.5 (51-76) |  |
|  | 303 | Initiate systematic and timely referrals to organ donation as part of a team in accordance with national policy when there is potential for organ and tissue donation | 61 (05-80) |  | |
| Donor/recipient support | 304 | Able to support physiologic optimization with donor care bundles (cardiovascular, endocrine/metabolic, respiratory, renal, hematologic, temperature) | 60 (50-90) |  | |
|  | 305 | Provide appropriate care to patients undergoing organ donation | 68 (50-100) |  | |
| Collaboration with teams for organ transplantation | 306 | Work with organ donor teams | 70 (50-100) | 71 (60-88) | 70.5 (60-81) |
|  | 307 | Understand the nurse's role in organ donation and access to resources | 66 (50-90) |  | |
| Assistance with brain death determination | 308 | Understand the prerequisites for testing for brainstem death | 70 (50-100) | 70 (60-82) | 70 (60.5-80) |
|  | 309 | Explain the method of determining brain death | 70 (50-80) | 70 (60-80) | 70 (60-72.5) |
|  | 310 | Manage relevant equipment for brainstem testing and assist with testing | 60 (34-83) |  | |
|  | 311 | Various procedures can be performed, such as recording the time of death | 99 (52-100) | 76.5 (54-95) | 72.5 (60-81.5) |
| Support for brain-dead family members | 312 | Discuss organ donation with family members as needed | 80 (50-100) | 78 (56-85) | 71 (59.5-80) |
|  | 313 | Explain the donor's condition, what will be done, and how to relieve the family's anxiety | 78 (45-100) | 68 (52-81) |  |
|  | 314 | Explain the purpose of each test to the family and explain that the patient is not in pain | 77 (50-100) | 69 (50-84) |  |
|  | 315 | Respond to family requests to visit the patient during the examination and provide immediate support in case of distress | 77 (60-100) | 75 (70-84) | 76 (65-82) |
|  | 316 | Provide reassurance and appropriate methods when family members are distressed or express concern about organ donation | 80 (50-100) | 77.5 (65-89) | 75 (60-81) |
| Legal Understanding for Organ Transplantation | 317 | Understand current national policies, protocols, and guidelines for brain stem testing and organ donation | 70 (45-82) |  | |
|  | 318 | Understands procedures for early identification of potential organ donors and organized and timely referral of potential organ donors | 58 (50-80) |  | |
|  | 319 | Understand the procedures for planning and implementing a collaborative approach to families to obtain consent for organ and tissue donation | 61 (45-80) |  | |
|  | 320 | Understand the legal, ethical, and consent issues related to organ and tissue donation for transplantation and research | 65 (47-80) |  | |
|  | 321 | Understands patient confidentiality issues related to organ transplantation | 80 (60-100) | 82 (74-90) | 80 (70-85) |
| **2. Caring** | | |  |  |  |
| **2.1. Nursing Diagnosis and Planning** | | |  |  |  |
| Collect appropriate information on critically ill patients | 322 | Collect physiological, psychosocial, cultural, developmental, and spiritual data with appropriate resources, depending on the patient's condition | 75 (65-90) | 77.5 (70-85) | 78 (70-81) |
|  | 323 | Use appropriate evidence-based assessment techniques to determine a patient's overall needs (including learning needs, psychological support, and psycho-social needs) | 80 (65-90) | 80 (72-90) | 80 (75-87) |
| Appropriate assessment of critically ill patients | 324 | Use appropriate evidence-based assessment techniques and tools to get a complete picture of the patient | 80 (74-90) | 80 (71-90) | 81 (80-85) |
|  | 325 | Prioritize data collection activities according to patient characteristics | 85 (80-90) | 80 (72-90) | 80 (77-85) |
|  | 326 | Relate collected data to current and predicted future conditions according to patient characteristics | 80 (75-85) | 80 (72-87) | 80 (75-85) |
|  | 327 | Analyze data from multiple sources to determine patient and family needs | 80 (73-90) | 80 (76-89) | 80 (80-85) |
| Develop appropriate care plans for critically ill patients | 328 | Identify and prioritize evidence-based interventions to promote and restore health and prevent further disease and disability | 80 (70-85) | 80 (70-88) | 80 (77-84) |
|  | 329 | Develop goals that take into account relevant risks, benefits, current evidence, expertise, and costs | 78 (60-80) | 76.5 (63-90) | 77 (70-80) |
|  | 330 | Work with patients, families, and multidisciplinary teams to develop a plan | 81 (70-90) | 80 (72-90) | 80 (76-81) |
|  | 331 | Assess, document, and communicate pertinent and relevant data within the team | 80 (67-85) | 80 (67-90) | 80 (74-81) |
|  | 332 | Collaborate with patients, families, significant others, and professionals to develop a plan of care in a manner that promotes each member's contribution to the desired outcome | 75 (65-85) | 75 (70-82) | 75 (71-80) |
|  | 333 | Individualize and critically evaluate a plan of care, taking into account the individual requirements and clinical situation of the patient | 70 (65-89) | 75 (70-81) | 75 (71-80) |
|  | 334 | Recognize and understand the economic and resource implications when developing a plan of care | 70 (60-76) | 71.5 (66-81) | 72 (70-79) |
|  | 335 | Develop accurate care plans with a view to life after discharge from the hospital | 75 (55-80) | 75 (65-80) | 75 (70-80) |
|  | 336 | Document and communicate data, diagnoses and related issues, and measurable goals in a clear and searchable format within the team | 70 (60-80) | 75.5 (70-82) | 75 (70-80) |
| Assessment of Critical Care | 337 | Assess the effectiveness of interventions in a timely manner and modify treatment and care modalities as needed to achieve expected outcomes | 80 (64-100) | 77.5 (70-84) | 80 (75-81) |
|  | 338 | Involve patients, families, and multidisciplinary professionals in the evaluation process as needed | 72 (60-85) | 75 (65-81) | 75 (72-80) |
|  | 339 | Properly record the results of the evaluation | 80 (73-90) | 80 (75-87) | 80 (80-87) |
| **2.2. Relief of discomfort symptoms** | | |  |  |  |
| Nursing Practice for Pain | 340 | Assess, prevent, and treat pain, including pharmacological and non-pharmacological interventions | 85 (72-100) | 85 (77-90) | 85 (80-86) |
| Nursing practice for restlessness and sedation | 341 | Appropriate use of neuromuscular blocking agents and proper patient monitoring and assessment during use (including BIS: depth of sedation monitor)/TOF: nerve locator stimulator) | 70 (59-83) | 71 (60-80) | 75 (65-80) |
|  | 342 | Responsive to the assessment, prevention, and treatment of delirium, including pharmacological and non-pharmacological interventions | 80 (75-90) | 80 (73-90) | 80 (80-88) |
| Nursing Practice for Delirium | 343 | Respond to the assessment, prevention, and treatment of insomnia and sleep disorders, including pharmacological and non-pharmacological interventions | 85 (80-100) | 85 (77-90) | 85 (80-90) |
|  | 344 | Respond to the assessment, prevention, and treatment of anxiety, including pharmacological and non-pharmacological interventions | 85 (75-96) | 82.5 (75-90) | 82 (80-89) |
| Nursing Practice for Insomnia | 345 | Be responsive to the assessment, prevention, and treatment of insomnia and sleep disorders, including pharmacological and non-pharmacological interventions | 80 (71-90) | 80 (75-85) | 80 (80-88) |
| Nursing practice for anxiety, depression, fear, etc. | 346 | Respond to the assessment, prevention, and treatment of anxiety, including pharmacological and non-pharmacological interventions | 80 (70-90) | 80.5 (75-85) | 81 (79-85) |
|  | 347 | Ensure that patients and families make informed treatment and care choices and understand the consequences | 80 (70-95) | 81 (75-90) | 80 (75-85) |
|  | 348 | Provide supportive care and coaching for patients and families during difficult procedures | 76 (66-90) | 77.5 (70-81) | 80 (75-81) |
|  | 349 | Minimize the psychological impact related to critical illness and treatment on the patient and family | 75 (65-90) | 78 (75-80) | 80 (76-84) |
|  | 350 | Respond to patients and families in the event of bereavement or traumatic events | 80 (65-90) | 80 (71-87) | 80 (73-82) |
| Nursing practice for other discomfort symptoms | 351 | Assess, prevent, and treat discomfort symptoms such as dry mouth, dyspnea, and fatigue, including pharmacological and non-pharmacological interventions | 80 (70-90) | 80 (72-80) | 80 (74-85) |
| **2.3. Rehabilitation of Critically Ill Patients / PICS** | | |  |  |  |
| Understanding and implementing the rehabilitation needs of patients and families | 352 | Explain PICS/PICS-F and its risks and prevention methods | 84 (75-100) | 80 (70-85) | 80 (75-85) |
|  | 353 | Explain the challenges of rehabilitation in the critical care area | 80 (75-92) | 80 (75-85) | 84 (77-86) |
|  | 354 | Understand why critically ill patients need ongoing rehabilitation | 90 (76-100) | 85 (75-90) | 85 (80-90) |
|  | 355 | Set rehabilitation goals (short, medium, and long term) appropriate for individual critically ill patients | 80 (70-90) | 79 (70-85) | 80 (75-85) |
|  | 356 | Coordinate rehabilitation of critically ill patients according to guidelines and other recommendations | 80 (70-90) | 80 (72-85) | 80 (79-85) |
|  | 357 | Understand and recognize rehabilitation prescriptions for critically ill patients | 80 (70-95) | 80 (75-85) | 80 (75-85) |
|  | 358 | Utilize specialized therapies for critical care rehabilitation | 75 (64-90) | 76 (73-80) | 78 (75-82) |
|  | 359 | Understand the issue of individual patient diversity and how it affects the patient's rehabilitation needs | 80 (70-90) | 80 (73-84) | 80 (75-82) |
|  | 360 | Properly assess the effectiveness of rehabilitation | 75 (70-95) | 75 (70-82) | 75 (74-80) |
| Nursing practice for maintenance and recovery of physical functions | 361 | Practice positioning of critically ill patients | 90 (80-100) | 90 (80-90) | 90 (85-92) |
|  | 362 | Practice joint range of motion training in critically ill patients | 80 (70-90) | 82.5 (73-90) | 85 (75-86) |
|  | 363 | Practice respiratory rehabilitation of critically ill patients | 82 (75-91) | 80 (78-90) | 85 (80-88) |
|  | 364 | Practice swallowing rehabilitation in critically ill patients | 75 (66-90) | 75 (69-80) | 80 (75-90) |
|  | 365 | Practice early mobilization of critically ill patients | 88 (79-100) | 86 (79-92) | 87 (85-90) |
|  | 366 | Implement practices to maintain motor function and improve ADLs in critically ill patients | 80 (75-97) | 80 (76-87) | 85 (80-88) |
|  | 367 | Understand risk assessment related to rehabilitation of critically ill patients and rehabilitate them | 85 (74-100) | 83.5 (76-87) | 85 (80-89) |
| Nursing practice for maintenance and recovery of cognitive function and mental health | 368 | Explain rehabilitation methods to prevent and ameliorate cognitive dysfunction and delirium | 77 (70-90) | 80 (74-85) | 84 (80-89) |
|  | 369 | Explain how to prevent and improve mental health disorders (depression, anxiety, PTSD) in patients and families | 75 (65-90) | 80 (73-82) | 81 (80-85) |
|  | 370 | Understand the advantages and disadvantages of ICU diaries in recovery from serious illness | 70 (56-85) | 71 (60-80) | 76 (70-80) |
|  | 371 | Understand and comply with legal and ethical considerations regarding ICU diaries | 71 (58-85) | 70 (60-80) | 73 (70-80) |
| Utilize available resources for rehabilitation of critically ill patients | 372 | Involve the patient and family in the rehabilitation process, including prompt referral to appropriate multidisciplinary team members and family involvement | 75 (60-84) | 75 (70-80) | 75 (74-80) |
|  | 373 | Help facilitate health and lifestyle changes appropriate to the patient and family | 73 (60-80) | 73 (70-78) | 75 (70-80) |
| **2.4. End of life care** | | |  |  |  |
| Evaluation of patient prognosis | 374 | Assess the patient severity | 90 (80-100) | 90 (80-91) | 90 (80-93) |
|  | 375 | Appropriate prognostic evaluation of critically ill patients | 80 (61-90) | 80 (75-82) | 80 (75-87) |
|  | 376 | Be prepared to anticipate, prevent, and recognize life-threatening situations and intervene | 85 (80-100) | 85 (79-90) | 85 (80-90) |
| Advance Care Planning | 377 | Encourage patients and families to discuss advance care planning and directives | 76 (55-88) | 79 (65-81) | 78 (72-81) |
|  | 378 | Utilize appropriate resources to guide ethically complex situations and find effective coping strategies and possible solutions | 80 (65-90) | 80 (72-85) | 80 (75-83) |
| Withholding or withdrawing treatment | 379 | Explain the palliative care procedures for critically ill patients | 75 (52-90) | 75 (71-80) | 75 (70-85) |
|  | 380 | Communicate care plans and discuss end-of-life care with patients and families | 80 (69-90) | 80 (75-87) | 80 (71-83) |
|  | 381 | Discuss end-of-life care plans with the patient's family |  | | 80 (74-85) |
|  | 382 | Provide symptom relief and individualized treatment and care plans for terminally ill patients according to the patient's needs | 80 (65-90) | 80 (75-85) | 80 (75-85) |
|  | 383 | In a multidisciplinary team, assess futility and other factors and consider withholding or terminating treatment as appropriate | 80 (70-90) | 80 (74-82) | 80 (75-85) |
|  | 384 | Consider end-of-life care options appropriate for the patient, taking into account the patient's and family's wishes, including the choice of ward after treatment is discontinued. | 80 (75-95) | 80 (77-84) | 80 (77-85) |
|  | 385 | Understand the procedures for forming and recording agreements regarding treatment cessation, including legal restrictions on treatment cessation or withholding, mental capacity laws, and ethical principles | 80 (60-90) | 80 (73-85) | 80 (76-83) |
|  | 386 | Understand what care is appropriate for the patient after discontinuation of treatment | 80 (75-90) | 80 (79-85) | 81 (80-86) |
| Assess, monitor, and observe terminally ill patients | 387 | Observe and assess symptoms of terminally ill patients, including pain, nausea, agitation, dyspnea, and fatigue. | 80 (75-100) | 81 (79-87) | 85 (80-87) |
|  | 388 | Recognize that the palliative approach integrates palliative care principles (symptom management, patient-centered care) throughout the patient's life and is not just for the last few days of life | 90 (75-100) | 87.5 (78-90) | 85 (80-90) |
|  | 389 | Work with patients, families, and professionals to determine end-of-life wishes, identify available resources, and implement strategies to promote dignity, comfort, and quality care at the end of life | 80 (70-90) | 82 (70-88) | 84 (79-87) |
|  | 390 | Seek and incorporate patient and family input to provide quality end-of-life care that meets their needs | 80 (80-100) | 82.5 (80-86) | 85 (77-87) |
| Utilize appropriate resources for end-of-life issues | 391 | Work with multidisciplinary teams to facilitate palliative care and end-of-life discussions, decisions, and care | 81 (70-90) | 80 (75-87) | 80 (80-85) |
|  | 392 | Maintain ongoing communication with family and professionals regarding palliative approaches/care at the end of life and provide ongoing emotional support | 80 (70-92) | 80 (76-88) | 82 (80-86) |
|  | 393 | Can refer to social support such as bereavement support | 60 (50-76) |  | |
| **2.5. Providing an ICU environment that promotes healing** | | |  |  |  |
| Environmental management to promote healing for patients and families | 394 | Appropriate assessment of patient's sensory perceptions, such as noise, illumination, smell, care, and other stimuli | 80 (70-95) | 80 (77-88) | 81 (80-89) |
|  | 395 | Ability to manage the environment appropriately for the patient's senses, including noise, lighting, smells, care, and other stimuli | 85 (75-98) | 83 (80-90) | 84 (78-90) |
|  | 396 | Minimize environmental risk factors that could cause physical harm or injury to patients, families, and health care providers | 81 (70-95) | 80.5 (79-90) | 83 (79-87) |
|  | 397 | Can promote proper day/night sleep cycles for critical care patients | 90 (80-100) | 88 (80-90) | 86 (80-90) |
|  | 398 | Able to provide a safe, healing and caring environment that respects the individual | 91 (80-100) | 89 (83-90) | 90 (81-90) |
|  | 399 | Able to effectively facilitate patient orientation | 80 (70-90) | 83 (78-90) | 85 (80-89) |
| Improve patient and family services | 400 | Obtain and act upon feedback and experiences from patients, caregivers and service users | 78 (60-85) | 78 (70-85) | 80 (77-85) |
|  | 401 | Question existing ways of doing things and propose new ways to current performance and culture | 80 (72-90) | 80 (72-85) | 82 (80-89) |
|  | 402 | Contribute to change management initiatives led by experienced colleagues | 80 (70-93) | 82.5 (80-88) | 85 (80-90) |
|  | 403 | Contribute to quality improvement initiatives taking place within the department | 85 (75-95) | 85 (76-90) | 85 (80-90) |
|  | 404 | Able to practice care that restores, supports, promotes, and maintains physiological and psychosocial stability for patients of all ages across the lifespan | 80 (70-90) | 82 (77-86) | 83 (80-85) |
| **3. Advocate and moral agent** | | |  |  |  |
| **3.1. Support decision-making** | | |  |  |  |
| Support for decision-making | 405 | Take into account patient and family preferences for treatment and intervention | 85 (73-92) | 85 (70-88) | 85 (80-85) |
|  | 406 | Support ethical decision making | 85 (75-100) | 85.5 (80-90) | 85 (80-88) |
|  | 407 | Provide information to help patient families make decisions | 84 (75-96) | 83 (78-88) | 85 (80-89) |
|  | 408 | Leverage and use a variety of evidence in making complex decisions | 76 (57-88) | 79.5 (70-84) | 80 (80-85) |
|  | 409 | Support problem solving and decision making for a variety of clinical situations for the patient's family | 80 (71-90) | 80 (73-87) | 80 (78-86) |
|  | 410 | Consider patient and family preferences for treatment and interventions | 85 (78-94) | 84.5 (78-90) | 85 (80-89) |
|  | 411 | Embracing equality and diversity and respecting without discrimination age, gender, religion, sexual orientation, race, disability, sentiments, social status, etc. | 88 (80-100) | 85 (79-90) | 85 (80-90) |
| **3.2. Ethical Practices** | | |  |  |  |
| Practices based on ethical principles and compliance with the law | 412 | Act ethically and responsibly and participate in ethical discussions and decision-making processes | 90 (78-100) | 86 (80-90) | 85 (80-90) |
|  | 413 | Understand ethical practices, autonomy, equality and diversity, and legal regulations to protect the rights of patients and families | 87 (75-99) | 85 (76-90) | 85 (78-88) |
|  | 414 | Practices that consistently nurture the autonomy, dignity, values, beliefs, and rights of patients and their families | 85 (75-92) | 85 (75-90) | 85 (81-89) |
|  | 415 | Articulate an understanding of ethical principles relevant to the delivery of critical care | 85 (75-100) | 85 (70-90) | 85 (80-87) |
|  | 416 | Promote ethical accountability and integrity in relationships, organizational decision-making, and resource management | 80 (70-96) | 80 (74-87) | 85 (80-88) |
|  | 417 | Document patient care and its ongoing evaluation in a clear, concise, accurate and timely manner, while respecting privacy and confidentiality of personal information | 80 (70-92) | 80.5 (77-85) | 80 (80-86) |
|  | 418 | Provide care in a fair and equitable manner that meets the diverse needs of patients, families, and the community | 90 (76-100) | 88 (80-91) | 85 (80-90) |
|  | 419 | Report unethical, illegal, or impaired conduct | 90 (80-100) | 90 (80-95) | 90 (85-90) |
|  | 420 | Demonstrate awareness of patient autonomy, consent, and relevant local and national laws | 85 (70-90) | 83.5 (80-89) | 85 (80-90) |
|  | 421 | Demonstrate responsibility for self-care and self-advocacy | 80 (68-90) | 90 (83-99) | 92 (90-100) |
|  | 422 | Understand and comply with national regulations and laws, such as the Medical Care Act and the Health and Nursing Care Act | 90 (73-100) | 90 (75-91) | 90 (82-91) |
|  | 423 | Protect patient confidentiality within legal regulations | 92 (81-100) | 90 (80-95) | 90 (85-92) |
|  | 424 | Take into consideration and practice standards of confidentiality, data protection, and documentation | 95 (85-100) | 90 (80-99) | 90 (85-94) |
|  | 425 | Comply with working conditions, employment rights, and work environment considerations set by the state and the facility (e.g., disposal of hazardous materials) | 90 (80-100) | 90 (80-95) | 90 (82-93) |
|  | 426 | Understand and comply with local and national regulations and laws regarding the prevention, reporting, and monitoring of adverse events, including medication errors, adverse events, and equipment malfunctions | 87 (80-100) | 90 (80-95) | 90 (81-93) |
|  | 427 | Working conditions, employment rights, and working environment considerations (e.g., disposal of hazardous materials) | 89 (80-100) | 87 (80-95) | 87 (84-90) |
|  | 428 | Recognize and comply with court decisions and laws relevant to their role | 85 (69-91) | 85 (73-91) | 85 (80-90) |
|  | 429 | Demonstrate an understanding of the law as it relates to patient care and health care delivery | 80 (65-90) | 83 (71-90) | 85 (80-90) |
| Practices for resolving ethical issues | 430 | Use available resources in making ethical decisions | 84 (73-95) | 84 (72-90) | 85 (80-88) |
|  | 431 | Make ethical decisions using available resources | 85 (75-95) | 85 (75-90) | 84 (80-87) |
|  | 432 | Address the needs of patients and families facing unanticipated treatment, quality of life, and end-of-life decisions | 85 (78-95) | 85 (80-90) | 85 (80-87) |
|  | 433 | Maintain a therapeutic and professional nurse-patient relationship within the appropriate role assignment | 87 (80-100) | 86 (80-90) | 85 (80-87) |
|  | 434 | Contribute to the resolution of ethical issues involving patients, families, and multidisciplinary teams | 83 (70-100) | 80 (70-86) | 80 (70-88) |
| **3.3. Patient and Family Communication** | | |  |  |  |
| Communicate and provide information in a respectful manner to patients and families | 435 | Respect others' viewpoints in discussions with patients and families | 86 (70-100) | 85 (77-93) | 85 (80-90) |
|  | 436 | Attentive to patient and family wishes and communicate information in a respectful manner according to needs, developmental stage, and level of understanding | 85 (70-100) | 88 (78-92) | 90 (80-100) |
|  | 437 | Communicate effectively with patients and families regarding treatment and nursing plans and the patient's clinical status | 80 (70-100) | 84 (80-99) | 85 (75-90) |
|  | 438 | Effectively communicate and explain difficult clinical information using terms and language that patients and families can understand | 80 (70-100) | 83 (79-96) | 84 (80-90) |
|  | 439 | Effectively communicate with and support patients and families in crisis situations | 80 (70-90) | 85 (75-93) | 81 (75-90) |
|  | 440 | Effective communication in complex situations. | 75 (69-90) | 80 (70-88) | 80 (67-84) |
|  | 441 | Notify bad news in a sensitive and compassionate way | 80 (70-92) | 80 (75-86) | 80 (73-85) |
| Accountability for nursing practice (informed consent) | 442 | Explain their roles and responsibilities to patients and families and facilitate relationship building | 80 (60-90) | 80 (74-92) | 80 (74-86) |
|  | 443 | Recognize and comply with regulations and laws related to the nursing role | 80 (70-100) | 80 (72-90) | 81 (80-90) |
|  | 444 | Indicate an understanding of national laws related to patient care and health care delivery | 64 (51-85) |  | |
|  | 445 | Explain treatment options to patients and families and assist in facilitating informed decision making | 80 (65-100) | 80 (70-89) | 80 (70-83) |
|  | 446 | Enable patients and families to make informed choices and understand the consequences | 85 (70-100) | 81 (80-90) | 82 (77-86) |
| **4. Evidence-based practice** | | |  |  |  |
| **4.1. Quality assurance and improvement of care (PDCA)** | | |  |  |  |
| Quality Assessment and Improvement Activities | 447 | Actively pursue new knowledge and skills to provide quality nursing care | 90 (80-100) | 90 (80-99) | 90 (80-95) |
|  | 448 | Implement practices to improve care processes and outcomes based on evidence, expertise, and patient preferences | 85 (71-100) | 85 (70-90) | 80 (74-87) |
|  | 449 | Explain clinical problems using evidence from the department or organization, including data on patient outcomes and facility quality improvement | 80 (66-90) | 75 (69-85) | 75 (70-80) |
|  | 450 | Participate in the development, implementation, evaluation, and revision of facility policies and procedures to improve the quality and effectiveness of nursing practice | 80 (70-90) | 80 (60-85) | 75 (68-80) |
|  | 451 | Ensure that nursing practices are in accordance with formal procedures and regulations | 90 (80-100) | 88 (70-100) | 83 (76-90) |
|  | 452 | Ongoing monitoring and evaluation of care processes and outcomes to determine best practices for individuals, groups, and populations | 80 (70-93) | 80 (70-90) | 80 (70-82) |
|  | 453 | Ability to question medical practices as needed to improve safety and quality | 90 (80-100) | 85 (75-96) | 83 (80-90) |
|  | 454 | Propose practices to challenge performance and culture | 65 (52-80) |  | |
|  | 455 | Improve services together with colleagues | 80 (70-91) | 80 (70-89) | 80 (77-86) |
| Promoting Evidence-Based Practices | 456 | Know how to search for evidence and literature using available resources | 80 (67-90) | 80 (70-90) | 80 (75-85) |
|  | 457 | Keep updated and current in knowledge of evidence-based practice | 80 (70-100) | 80 (70-90) | 80 (70-87) |
|  | 458 | Understand the importance of evidence-based practice and be able to critically evaluate the literature | 70 (60-80) | 70 (60-80) | 75 (67-80) |
|  | 459 | Demonstrate an understanding of evidence-based practice and its translation into clinical practice | 75 (60-85) | 70 (61-80) | 75 (70-80) |
|  | 460 | Engage in and contribute to evidence-based critical care nursing practice | 85 (71-100) | 80 (70-88) | 80 (75-80) |
|  | 461 | Know what care practices are based on the guidelines | 85 (75-94) | 80 (70-90) | 80 (75-85) |
|  | 462 | Disseminate information on evidence-based best practices | 77 (70-85) | 75 (69-80) | 77 (70-80) |
| **4.2. Clinical research** | | |  |  |  |
| Research activities to establish evidence | 463 | Explain research questions based on clinical practice to improve quality of care | 70 (59-80) | 70 (63-80) | 70 (65-78) |
|  | 464 | Promote research, evidence-based practice, and the dissemination of nursing knowledge | 70 (57-83) | 75 (65-84) | 74 (69-80) |
|  | 465 | Participation in research activities | 71 (60-80) | 75 (70-82) | 75 (70-80) |
|  | 466 | Participate in the development of evidence-based protocols and guidelines | 70 (41-75) | 70 (60-80) | 70 (65-80) |
|  | 467 | Facilitate the development and implementation of innovative solutions in intensive care nursing | 60 (40-80) |  | |
| **5. Collaboration and management ability** | | |  |  |  |
| **5.1. Unit Management** | | |  |  |  |
| Therapeutic intervention and system management | 468 | Clearly explain the role, function, and benefits of a critical care organization at your facility | 80 (68-90) | 78 (66-82) | 79 (70-80) |
|  | 469 | Organizational awareness of how services related to critical care are planned at their facility | 70 (60-82) | 70 (60-80) | 70 (68-80) |
|  | 470 | Understands the services and improvement plans related to critical care at his/her facility | 74 (65-85) | 70 (60-80) | 70 (67-77) |
|  | 471 | Implement service plans and projects related to critical care at their facility | 71 (60-85) | 70 (60-78) | 78 (70-80) |
|  | 472 | Assist in the development of guidelines and policies for critical care practices at their facility | 70 (50-80) | 65 (60-80) |  |
|  | 473 | Incorporate professional, legal, ethical, and critical care standards into practice | 80 (70-90) | 75 (60-82) | 75 (70-81) |
|  | 474 | Political awareness of key policies affecting critical services in the country | 60 (40-70) |  | |
| Logistics Management and Cost Awareness | 475 | Implement cost and waste reduction practices | 76 (62-85) | 77 (70-85) | 80 (73-85) |
|  | 476 | Recognize the nurse's financial responsibilities and role in managing the logistics of the organization | 68 (50-80) |  | |
|  | 477 | Identify and report logistics management issues (e.g., insufficient stock, damaged equipment, supplies not fit for purpose) through appropriate systems | 70 (60-85) | 75 (70-85) | 78 (71-84) |
|  | 478 | Effective use of logistics systems including capacity, supplies, equipment, and consumables | 70 (54-80) | 75 (66-80) | 77 (70-82) |
| Participation in organizational activities | 479 | Participate in committees, councils, and multidisciplinary teams | 80 (71-100) | 81 (70-90) | 81 (78-87) |
|  | 480 | Promote professional development through participation in professional organizations | 75 (60-85) | 75 (70-80) | 78 (70-84) |
|  | 481 | Participate in strategies and activities to promote healthy communities | 65 (46-80) |  | |
|  | 482 | Facilitate the development and implementation of innovative solutions in intensive care nursing | 70 (40-80) |  | |
| Healthy work environment and work style | 483 | Demonstrate an understanding of work-life balance | 82 (75-100) | 85 (77-94) | 85 (80-90) |
|  | 484 | Work-life balance. | 85 (80-100) | 85 (78-95) | 85 (80-90) |
|  | 485 | Contribute to and support efforts to improve the quality of the critical care environment and work-life balance at your facility | 71 (61-80) | 75 (70-80) | 79 (72-81) |
|  | 486 | Contribute to the creation and maintenance of a healthy work environment | 80 (70-98) | 80 (75-86) | 80 (77-84) |
|  | 487 | Indicate flexibility and patient focus in a rapidly changing environment | 90 (76-100) | 90 (80-100) | 85 (80-92) |
|  | 488 | Ask for an appropriate number of staff with the knowledge and skills to care for critically ill patients when needed | 82 (75-100) | 80 (75-90) | 80 (78-85) |
|  | 489 | Adhere to standards governing the behavior of health care professionals to create a healthy work environment that promotes cooperation, respect, and trust | 88 (75-100) | 85 (78-94) | 83 (80-88) |
|  | 490 | Understand, embrace and actively participate in change management and related processes | 80 (64-90) | 80 (66-86) | 80 (79-85) |
|  | 491 | Maintain a physical and psychosocial environment that promotes safety, security, and optimal health | 80 (75-95) | 80 (75-90) | 81 (76-85) |
| **5.2. Team Management** | | |  |  |  |
| Membership and Followership | 492 | Recognize common team objectives and respect team decisions | 85 (80-100) | 85 (80-90) | 83 (76-86) |
|  | 493 | Adopt a team approach and be able to recognize and appreciate efforts, contributions and compromises | 80 (70-90) | 80 (77-90) | 80 (77-85) |
|  | 494 | Share information widely with staff | 95 (81-100) | 90 (80-100) | 90 (80-96) |
|  | 495 | Care plans can be shared | 90 (80-100) | 90 (80-100) | 88 (80-95) |
|  | 496 | Share knowledge and collaborate with others to create change and positive outcomes | 85 (74-98) | 83 (80-95) | 80 (73-85) |
|  | 497 | Actively seek the opinions of others | 90 (80-100) | 88 (80-95) | 85 (80-90) |
|  | 498 | Utilize SBAR and report appropriately to team members and other professionals | 87 (80-100) | 88 (80-97) | 85 (79-90) |
|  | 499 | Able to recognize, respect, and promote cooperation with staff | 93 (80-100) | 92 (80-100) | 90 (80-95) |
|  | 500 | Respect others' viewpoints and actively ask their opinions in discussions with other healthcare professionals. | 90 (80-100) | 85 (80-100) | 86 (80-90) |
|  | 501 | Recognize and participate in the need to support and debrief with colleagues and each other | 89 (80-100) | 85 (76-100) | 85 (80-90) |
|  | 502 | Support others in providing good patient care and better service | 88 (80-100) | 85 (80-93) | 85 (80-90) |
|  | 503 | Opportunities to be more effective by working with others | 90 (80-100) | 85 (80-93) | 85 (80-90) |
|  | 504 | Proactively and independently derive solutions to recurring problems and issues | 80 (71-98) | 80 (73-90) | 80 (75-80) |
| leadership | 505 | Clearly recognize their role, responsibilities, and purpose within the nursing team | 87 (80-100) | 85 (80-90) | 85 (80-90) |
|  | 506 | Have a sense of role in the team in order to form an effective team | 83 (80-98) | 85 (80-90) | 82 (80-86) |
|  | 507 | Demonstrate effective interpersonal communication, leadership, negotiation, and conflict resolution skills to build positive relationships with colleagues, patients, and families | 80 (75-95) | 82 (74-90) | 80 (75-85) |
|  | 508 | Facilitate collaboration among team members | 81 (75-90) | 84 (77-85) | 80 (75-86) |
|  | 509 | Foster a spirit of transparency, trust, and respect among team members | 88 (75-100) | 80 (76-90) | 80 (76-85) |
|  | 510 | Provide leadership and support for service improvement initiatives within the department | 80 (78-91) | 80 (73-85) | 80 (75-85) |
|  | 511 | Emphasize the value of shared responsibility in decision-making and support shared leadership and coordination roles | 80 (70-90) | 80 (70-85) | 80 (75-85) |
|  | 512 | Resolve interpersonal and personal issues affecting individual and team performance | 80 (70-90) | 80 (70-85) | 78 (74-80) |
|  | 513 | Encourage open and constructive discussion and respect the diverse viewpoints of others | 88 (80-97) | 81 (80-90) | 80 (75-85) |
| Management and coordination of care and operations, stress management | 514 | Manage activities on a daily or shift basis | 87 (70-100) | 87 (80-91) | 85 (76-90) |
|  | 515 | Manage patient care to ensure that it is effective and efficient | 80 (71-97) | 85 (75-90) | 83 (75-87) |
|  | 516 | Plan and prioritize work and workload during the shift according to patient needs | 90 (80-100) | 88 (80-95) | 85 (81-90) |
|  | 517 | Adjust priorities and change work plans as circumstances dictate | 90 (80-100) | 85 (80-96) | 85 (80-90) |
|  | 518 | Delegate appropriate tasks to other team members as needed | 85 (80-100) | 85 (80-91) | 85 (80-90) |
|  | 519 | Assess and adjust the workload and workload of staff and other team members to meet the changing needs of the patient's family | 85 (75-96) | 85 (80-90) | 82 (80-90) |
|  | 520 | Effectively manage the workload in your department | 80 (70-92) | 80 (75-87) | 80 (78-85) |
|  | 521 | Identify and, if necessary, coordinate the work of professional or other positions (including clerical, etc.) | 80 (70-88) | 80 (70-85) | 80 (75-85) |
|  | 522 | Recognize and effectively utilize coping strategies to deal with stressful situations in the clinical environment | 82 (75-95) | 80 (73-90) | 80 (75-83) |
|  | 523 | Know how to prevent stressors in the workplace | 80 (70-89) | 80 (75-89) | 80 (70-85) |
|  | 524 | Observe and address the health status of other team members | 80 (70-90) | 80 (75-90) | 80 (73-87) |
| Use of other experts (consultation) | 525 | Assess individual patient needs and select the available resources (other professionals) to achieve the best outcomes | 85 (80-90) | 80 (72-90) | 80 (72-83) |
|  | 526 | Consider cost-effectiveness issues and use resources (other professionals) efficiently | 80 (70-84) | 78 (70-85) | 77 (70-83) |
|  | 527 | Facilitate sharing of information and resources (other professionals) among staff | 80 (74-90) | 80 (78-90) | 80 (75-85) |
|  | 528 | Suggest additional and necessary resources (other professionals) to improve nursing practice and quality of care for critically ill patients | 80 (70-89) | 80 (75-90) | 80 (74-85) |
|  | 529 | Assist patients and families in identifying and securing appropriate services to address their health-related needs, depending on available resources (other professionals) | 80 (70-86) | 80 (70-81) | 80 (71-85) |
|  | 530 | Collaborate to provide optimal care, respecting each other's roles, responsibilities, and abilities. | 85 (80-100) | 83 (78-90) | 84 (80-90) |
|  | 531 | Contribute to the development of effective interprofessional relationships to meet the needs of patients and their families | 80 (77-91) | 80 (73-90) | 80 (77-85) |
|  | 532 | Ability to consult with appropriate staff/professionals to develop/review care plans and facilitate continuity of care | 85 (75-99) | 85 (80-91) | 85 (80-89) |
|  | 533 | Work to resolve conflicts between multiple professions and contribute to interprofessional care planning | 80 (70-90) | 80 (70-86) | 80 (70-85) |
|  | 534 | Effective verbal and written communication with all members of the multidisciplinary team | 85 (80-100) | 83 (74-90) | 85 (77-85) |
|  | 535 | Ask questions of all members of the multidisciplinary team about the care process and the rationale supporting decisions | 80 (70-90) | 80 (71-85) | 80 (76-85) |
|  | 536 | Offer own professional perspective in discussions with multidisciplinary teams | 80 (76-100) | 80 (75-90) | 80 (80-85) |
|  | 537 | Communicate and collaborate effectively within a multidisciplinary team | 90 (80-100) | 89 (80-90) | 85 (80-90) |
|  | 538 | Choose appropriate methods in communicating with other professionals | 86 (78-100) | 84 (90-90) | 85 (80-90) |
|  | 539 | Explain and facilitate their roles and responsibilities to other members of the health care community | 85 (76-95) | 80 (76-89) | 80 (80-85) |
|  | 540 | Facilitate sharing of information and resources (other professionals) among staff | 85 (70-100) | 80 (80-90) | 80 (80-85) |
| **5.3. Medical safety** | | |  |  |  |
| Compliance with Nursing Standards | 541 | Understand and participate in clinical audits, including patient safety, environmental safety, and medical equipment maintenance | 80 (70-90) | 80 (75-90) | 80 (80-88) |
|  | 542 | Understand the value of quality indicators related to patient outcomes (e.g., length of stay, ventilatory time, prevention of infections, etc.) and participate in data collection | 70 (55-80) | 80 (72.5-87.5) | 80 (80-90) |
|  | 543 | Contribute to the collection of data on quality indicators related to patient outcomes (length of stay, ventilatory time, prevention of infections, etc.) |  | 70 (70-80) | 75 (70-84) |
|  | 544 | Understand and be able to apply academic and national recommendations, policies, and guidelines to the department | 70 (55-87) | 76 (68.5-84) | 75 (70-80) |
| Safety Culture and Incident Reporting | 545 | Understand their role in influencing the quality of safe and effective critical care services | 80 (64-100) | 80 (77.5-90) | 85 (80-90) |
|  | 546 | Respond quickly to protect patient safety | 100 (80-100) | 100 (94.5-100) | 100 (90-100) |
|  | 547 | Use safe and effective written, verbal, telephone, and electronic communication strategies | 80 (70-90) | 80 (75-90) | 80 (79-86) |
|  | 548 | Take necessary actions against actual or potential risks and incidents | 85 (80-100) | 88.5 (80-95) | 90 (83-90) |
|  | 549 | Identify and demonstrate observations and concerns regarding safety, hazards, and errors in the care and practice environment | 80 (70-90) | 80 (79-90) | 85 (80-90) |
|  | 550 | Provide feedback to the organization on issues related to the practices of others | 70 (67-80) | 72.5 (70-81.5) | 75 (70-84) |
|  | 551 | Report adverse or potential risks through the organization's internal incident reporting system | 90 (72-100) | 90 (80-100) | 90 (80-95) |
|  | 552 | Quickly identify serious or adverse incidents that occur in the hospital | 80 (70-90) | 84.5 (80-90) | 85 (80-90) |
|  | 553 | Respond appropriately to critical or adverse incidents | 90 (80-100) | 90 (80-100) | 90 (80-100) |
|  | 554 | Properly report serious and adverse incidents in accordance with unit, hospital, and national policies and protocols | 80 (60-100) | 80 (74-90) | 80 (78-85) |
|  | 555 | Understand and comply with local and national regulations and laws regarding the prevention, reporting, and monitoring of adverse events, including medication errors, adverse events, and equipment malfunctions | 80 (61-90) | 80 (71-90) | 80 (79-90) |
|  | 556 | Consider measures to prevent recurrence of medical accidents | 86 (80-100) | 90 (80-90) | 90 (85-95) |
|  | 557 | Actively participate in the recognition, response, disclosure, reporting, and prevention of recurrence of adverse events and near-misses | 90 (80-100) | 90 (80-90) | 90 (83-92) |
|  | 558 | Promote a safe culture of learning from and responding to risk | 85 (75-100) | 90 (85-90) | 90 (82-93) |
|  | 559 | Build a culture of safety for patients, families, and providers | 80 (65-95) | 84 (80-90) | 85 (80-90) |
| Awareness of infection prevention and toxic exposure | 560 | Safe handling and disposal of contaminated, biological, chemical, and toxic materials in accordance with local and national policies and protocols | 80 (50-90) | 80 (71-90) | 80 (80-90) |
|  | 561 | Promote proper use of disposable items in the department | 80 (60-100) | 80 (80-90) | 82 (80-90) |
|  | 562 | Communicate environmental health risks and exposure reduction measures to patients, families, and health care providers | 80 (75-99) | 80.5 (80-90) | 80 (80-89) |
|  | 563 | Use reliable sources of information to determine if a product or treatment has a negative impact on the environment | 71 (68-80) | 74.5 (70-80) | 77 (73-80) |
| Handling of complex medical equipment | 564 | Maintain a safe medical environment (e.g., bed space organization, emergency equipment) | 90 (80-100) | 90 (81.5-95) | 90 (80-95) |
|  | 565 | Safe use of medical equipment required for critical care | 100 (87-100) | 100 (88-100) | 100 (85-100) |
|  | 566 | Safely assist or perform advanced and complex procedures required for medical device operation | 90 (80-100) | 90 (81.5-95) | 90 (85-95) |
| Safe medication and medication administration | 567 | Maintain and update knowledge of medications administered in the critical care setting | 91 (80-100) | 90 (85.5-100) | 90 (85-94) |
|  | 568 | Knowing the right patient, the right time, the right route, the right medicine, the right dose, the right label, the right calculation, and the relevant details will ensure that medication errors are prevented | 95 (80-100) | 95 (85.5-100) | 95 (90-100) |
|  | 569 | Follow and promote unit, hospital, and national guidelines for critical care medications | 80 (65-94) | 80 (80-90) | 80 (78-90) |
|  | 570 | Participate in the development of protocols and guidelines for safe medication administration in critical care | 60 (46-80) |  | |
| **5.4. In-hospital and out-of-hospital patient transport** | | |  |  |  |
| Evaluation of indications for intra- and extra-hospital patient transport | 571 | Describe the bed management system at your facility | 70 (50-80) | 70 (69.5-80) | 74 (70-80) |
|  | 572 | Work as a team to perform a comprehensive risk assessment to ensure the patient is suitable for transport | 80 (65-90) | 80 (76-82.5) | 80 (75-85) |
|  | 573 | Explain the potential risks associated with transporting critically ill patients | 80 (70-92) | 80 (76-90) | 80 (76-84) |
|  | 574 | Explain what may happen to the patient during transport | 80 (76-96) | 80 (77.5-89) | 80 (76-85) |
|  | 575 | Determine if the transport is urgent and time-critical | 90 (73-95) | 85 (78.5-90) | 80 (80-85) |
|  | 576 | Review patient priorities and needs and coordinate patient transfers | 80 (70-96) | 80 (73.5-86.5) | 80 (76-84) |
|  | 577 | Reassess safety and risk factors prior to transport | 82 (70-100) | 80 (75.5-88.5) | 80 (78-85) |
| Preparation for in-hospital and out-of-hospital patient transport | 578 | Understand the preparation process for in-hospital/out-of-hospital transport of critically ill patients | 80 (70-95) | 80 (78-85) | 80 (79-85) |
|  | 579 | Understands the roles of team members in arranging and conducting intra-hospital and inter-hospital transports | 80 (70-95) | 80 (75-85.5) | 80 (76-85) |
|  | 580 | Assess patient needs prior to transport | 80 (72-90) | 80 (76-87) | 80 (78-88) |
|  | 581 | Assess patient's clinical condition prior to transfer out of the ICU | 88 (80-100) | 90 (81-95) | 90 (85-90) |
|  | 582 | Anticipate potential problems with conveyance and plan to reduce risk | 89 (80-94) | 85 (80-90) | 85 (80-90) |
|  | 583 | Prepare spare battery packs and alternative equipment in case of failure | 83 (70-90) | 85 (80-90) | 85 (80-90) |
|  | 584 | Prepare medical records, radiology results, and recent blood test results, etc prior to transport | 90 (80-99) | 90 (80-90.5) | 85 (80-91) |
|  | 585 | Prepared transport monitoring equipment | 95 (90-100) | 91 (86.5-100) | 90 (83-95) |
|  | 586 | Understand the ethical issues and legal requirements related to transfers | 70 (61-80) | 70.5 (70-80) | 74 (65-80) |
|  | 587 | Explain referrals to receiving hospitals (including critical care and specialty consultants) | 70 (55-86) | 70 (64-73) | 70 (65-80) |
|  | 588 | Explain the care that should be responsibly performed during transport | 80 (71-100) | 80 (80-90) | 80 (77-90) |
|  | 589 | Explain about indemnity insurance | 60 (30-80) |  | |
|  | 590 | Assess the competence and skills of transport personnel | 75 (60-90) | 78.5 (71.5-82) | 80 (75-83) |
|  | 591 | Consider contingency plans/backups | 80 (70-90) | 80 (76-90) | 80 (80-84) |
|  | 592 | Communicate with the receiving hospital that will transport the patient. | 80 (65-90) | 80 (70-85) | 80 (70-85) |
|  | 593 | Explain transport time issues | 76 (60-90) | 74.5 (68.5-80) | 79 (70-80) |
|  | 594 | Explain the types of transport available and their advantages (out-of-hospital: ambulance/helicopter transport, in-hospital: stretcher, wheelchair, etc.) | 80 (75-90) | 78 (70-83.5) | 80 (70-83) |
|  | 595 | Prepare and complete necessary paperwork such as transport documents, progress notes, nursing plans, etc. | 86 (80-100) | 84 (79.5-90) | 80 (78-90) |
|  | 596 | Work with patients, families, and multidisciplinary teams to facilitate effective and safe transport across care settings | 81 (70-90) | 80 (80-89) | 80 (76-83) |
| Practice and evaluation during transport | 597 | Monitor and assess changes in physiological indicators of patients during transport | 90 (80-100) | 90 (81.5-100) | 90 (80-95) |
|  | 598 | Manage necessary medications to patients during transport | 90 (80-100) | 90 (83-95) | 90 (80-95) |
|  | 599 | Appropriately recognize situations that may affect the quality and safety of critical care transfers | 90 (80-95) | 90 (81-91) | 89 (80-90) |
|  | 600 | Identify issues and problems that can be remedied during transport | 82 (73-95) | 81 (77.5-90) | 82 (78-86) |
|  | 601 | Reflect on the conveyance experience | 85 (80-100) | 85 (74.5-90) | 85 (80-90) |
|  | 602 | Coordinate the provision of ongoing care, including specialist treatment and observation, for patients being transported | 80 (65-90) | 80 (74-87.5) | 80 (77-85) |
| Communication necessary during transport, such as signing off | 603 | Provide information and informed consent for transporting a conscious patient | 90 (80-100) | 90 (84.5-91.5) | 90 (80-90) |
|  | 604 | Communicate with family members during transport and provide ongoing status reports as needed | 90 (80-100) | 90 (83-95) | 90 (80-90) |
|  | 605 | Communicate openly with multiple disciplines to minimize risks associated with patient transfers, intra-hospital transfers, and transitions of care | 89 (75-97) | 88.5 (80-90.5) | 85 (80-90) |
|  | 606 | Properly contact the receiving unit upon departure when moving out | 85 (80-100) | 85 (80-90) | 85 (75-90) |
|  | 607 | Communicate patient status and physiological requirements to transport team/staff | 90 (79-100) | 90 (85-93.5) | 85 (75-90) |
|  | 608 | Share information with the team related to safety, risk assessment, and planning for the unexpected regarding transport | 86 (74-96) | 85 (80-90) | 85 (80-90) |
|  | 609 | Appropriate transfer of patient status to receiving unit at time of transfer | 90 (81-100) | 90 (85-95) | 87 (80-90) |
| **6. Education and self-development ability** | | |  |  |  |
| **6.1. Self-development** | | |  |  |  |
| Introspective Practice | 610 | Actively ask feedback on your practice from patients, family members, colleagues, and professionals | 88 (80-95) | 85.5 (90-90) | 80 (75-90) |
|  | 611 | Reflect on nursing practice through an introspective and self-aware approach | 83 (71-100) | 81 (80-88) | 80 (80-90) |
|  | 612 | Reflect on current activities and identify how future activities should be developed | 80 (70-90) | 80 (80-87) | 80 (75-90) |
|  | 613 | Deepen self-awareness and recognize one's own strengths and limitations | 89 (80-100) | 90 (80-93) | 90 (84-94) |
|  | 614 | Admit failure and treat it as a learning opportunity | 90 (80-100) | 90 (82.5-95) | 90 (90-95) |
|  | 615 | Change behavior based on feedback and reflection | 90 (85-100) | 90(80-91) | 90 (83-92) |
|  | 616 | Maintain competence in critical care by reflecting on clinical experiences and participating in educational programs necessary for professional development | 90 (80-100) | 81.5 (80-90) | 80 (75-90) |
|  | 617 | Recognize their own strengths and limitations | 90 (80-100) | 90 (80-94.5) | 90 (83-99) |
| Self-development and professional development | 618 | Understand the need for education in critical care | 99 (89-100) | 99.5 (85-100) | 100 (89-100) |
|  | 619 | Facilitate the transfer of information and professional development through writing, publishing, and presentations to professional and general audiences | 80 (60-85) | 80 (70.5-82) | 80 (75-85) |
|  | 620 | Able to share acquired knowledge and skills | 81 (80-94) | 80 (80-90) | 85 (80-90) |
|  | 621 | Willingness to learn on one's own initiative | 90 (80-100) | 90 (80-100) | 90 (87-95) |
|  | 622 | Actively seek opportunities and challenges for personal learning and development | 90 (80-100) | 90 (80.5-98) | 90 (80-90) |
|  | 623 | Take initiative in one's own educational and professional development | 89 (80-100) | 90 (80.5-95.5) | 90 (81-91) |
|  | 624 | Know how to access learning and teaching opportunities/resources that support continuing professional development | 85 (70-97) | 84 (80-90) | 84 (80-90) |
|  | 625 | Recognize and, when possible, participate in local, national, and international conferences and committees related to critical care nursing | 80 (60-90) | 80 (70-85) | 80 (75-86) |
|  | 626 | Participate in ongoing learning experiences and activities to develop and maintain clinical and professional skills and knowledge | 80 (70-98) | 80 (77-88) | 80 (80-90) |
|  | 627 | Keep records and activity histories that provide evidence of professional competence and lifelong learning | 80 (60-95) | 80 (70-83.5) | 80 (73-85) |
| **6.2. Education** | | |  |  |  |
| Fostering a learning environment | 628 | Participate in activities that facilitate the teaching and learning of others in a multidisciplinary team | 80 (70-90) | 80 (74.5-82.5) | 81 (80-88) |
|  | 629 | Support individual and group self-development in a critical care environment | 80 (70-91) | 80 (78.5-85.5) | 80 (80-85) |
|  | 630 | Contribute to a work environment conducive to professional education | 80 (70-90) | 80 (75-81.5) | 80 (80-85) |
| Feedback and Coaching | 631 | Provide regular feedback to colleagues and acknowledge successes and need for improvement as needed | 80 (70-90) | 80 (78-84) | 80 (78-85) |
|  | 632 | Provide formal or informal constructive feedback on colleagues' practices and role performance | 75 (70-90) | 80 (71-80) | 80 (78-85) |
| Educational involvement with colleagues | 633 | Behave in a professional manner as a role model for juniors and students | 80 (75-90) | 80 (79-90) | 80 (79-85) |
|  | 634 | Serve as a resource, educator, role model, preceptor, advocate, and mentor to students, colleagues, and multidisciplinary team members | 80 (75-90) | 80 (79-85.5) | 80 (75-85) |
|  | 635 | Demonstrate ability to be an effective mentor and role model as needed | 83 (77-95) | 81 (78.5-88) | 80 (77-85) |
|  | 636 | Support staff and other colleagues through mentoring and other professional development | 80 (70-90) | 80 (77.5-83.5) | 80 (75-85) |
|  | 637 | Share acquired educational knowledge, experience, and ideas with peers | 83 (75-90) | 82 (80-85.5) | 84 (78-88) |
|  | 638 | Teach and review practices with colleagues | 80 (75-90) | 84.5 (80-90) | 85 (80-90) |
| training of new recruits | 639 | Support newcomers. | 90 (80-100) | 90 (86.5-100) | 90 (83-91) |
|  | 640 | Assist newcomers in developing their skills | 90 (80-100) | 88.5 (80-90) | 85 (80-90) |
|  | 641 | Motivate and encourage newcomers to develop their skills | 83 (72-91) | 83.5 (80-90) | 82 (80-85) |
|  | 642 | Guide, supervise, and assess nursing care provided by less experienced colleagues | 83 (74-95) | 84 (80-88) | 85 (80-90) |
|  | 643 | Coach colleagues on tasks, goals, processes, and performance standards | 80 (70-93) | 80 (74-86) | 80 (75-85) |
| Learning Support for Patients/Families | 644 | Educate patients and families as needed | 83 (80-98) | 85 (80-90) | 85 (80-88) |
|  | 645 | Provide patients and families with the study information they need | 84 (70-94) | 84.5 (80-90) | 85 (80-86) |

*Abbreviations: Acute lung injury; ALI, Acute respiratory distress syndrome; ARDS, Cannot ventilate and cannot intubate; CVCI, Stroke Volume Variation; SVV, Glasgow Coma Scale; GCS, Intracranial pressure; ICP, Cerebral perfusion pressure; CPP, ICU acquired weakness; ICU-AW, Ventilator associated event; VAE, Transfusion related acute lung injury; TRALI, Graft versus host disease; GVHD, Basic Life Support; BLS, Advanced Cardiovascular Life Support; ACLS, Intra Aortic Balloon Pumping; IABP, Venoarterial-extracorporeal membrane oxygenation; VA-ECMO, Venovenous-extracorporeal membrane oxygenation; VV-ECMO, Post intensive care syndrome/ post intensive care syndrome - family; PICS/PICS-F, Post Traumatic Stress Disorder; PTSD, Activities of Daily Living; ADL, Quality of Life; QOL, Medical Research Council; MRC, manual muscle testing; MMT
