## Supplementary material for "Clinical practice competencies for standard critical care nursing: Consensus statement based on a systematic review and Delphi survey": S6 table

**eTable6. Characteristics of clinical practice skills necessary to provide standard nursing care in the intensive care unit**

Based on previous research, we define those that meet the following as capable of providing standard nursing care in the intensive care unit, and list in a table the characteristics of clinical practice skills necessary to meet this definition for which there is social consensus.

The patient is able to immediately grasp the whole picture of a typical patient/family and plan and practice individualized nursing care for critically ill patients.

Based on the individuality of the critically ill patients/families, the system is able to deal with possible problems in a predictive and preventive manner.

The ability to practice nursing care that is holistic and respectful of the values of patients and families with serious illnesses.

He tries to use the latest guidelines and information for nursing.

Recognize the limitations of their abilities and consult with the necessary resources (other nurses, multidisciplinary professionals).

Participate in the training of new and inexperienced staff.

Please note that this does not imply or imply that all of the listed clinical practice competencies must be met in order to be able to provide standard nursing care in the intensive care unit.

| **1. Therapeutic Management of Disease and Clinical Decision Making** | | |
| --- | --- | --- |
| **1.1. Respiratory system** | | |
| Anatomy and Physiology of the Respiratory System and Understanding of Disease | 1 | Understand the anatomy and physiology involved in the respiratory system |
|  | 2 | Explain the four components of internal and external respiration, cellular respiration, acid-base equilibrium, and respiratory failure |
|  | 3 | Explain the pathophysiology, causes, signs, symptoms, and therapeutic management of respiratory diseases such as pneumonia, asthma, chronic obstructive airway disease, acute lung injury syndrome (ALI/ARDS), pulmonary embolism, and pneumothorax |
| Nursing Practice for Respiratory System | 4 | Perform assessment and nursing practice to improve respiratory function |
|  | 5 | Understand the complications associated with suctioning airway secretions and perform suctioning using methods that minimize or prevent them |
|  | 6 | Appropriately assess the risk of difficult airway clearance (cannot ventilate and cannot intubate: CVCI) and explain how to respond to it |
|  | 7 | Explain the impact, benefits, and risks of each position performed as respiratory therapy |
|  | 8 | Explain how to position to optimize respiratory function |
|  | 9 | Report abnormalities or changes in the respiratory system to the appropriate health care professions |
| Observation, monitoring and assessment of the respiratory system | 10 | Assess the airway and secure the airway |
|  | 11 | Assess for airway stenosis or obstruction |
|  | 12 | Assess the amount and nature of airway secretions and sputum culture results |
|  | 13 | Assess respiratory function based on respiratory frequency, breathing patterns, and use of accessory respiratory muscles |
|  | 14 | Assess for diminished or absent breath sounds, left-right differences, and abnormal sounds |
|  | 15 | Identify abnormal respiratory condition such as seesaw breathing, cyanosis, subcutaneous emphysema, uneven chest expansion, tension pneumothorax, hypoxia, restlessness, and altered mental status |
|  | 16 | Use of respiratory monitors such as SpO2, SVO2, and capnography |
|  | 17 | Explain the need for blood gas analysis testing |
|  | 18 | Assess the results of blood gas analysis |
|  | 19 | Determine the patient's condition from chest X-P, CT, and MRI (including readings) |
| Administration of medicine related to the respiratory system | 20 | Safely prepare and administer medications related to the treatment of the respiratory system |
|  | 21 | Assess the effects of medicines related to the respiratory system and adjust care and treatment according to the patient's condition |
|  | 22 | Assess airway and breathing during procedures requiring sedation |
|  | 23 | Monitor sedation depth and sedative dosage |
|  | 24 | Practice care of patients with continuous inhalation of nitric oxide |
|  | 25 | Prepare, perform, and change circuits of nebulizers |
| **1.2.** Cardiovascular system | | |
| Anatomy and Physiology of the Cardiovascular System and Understanding of Disease | 26 | Understand the anatomy and physiology related to the cardiovascular system |
|  | 27 | Explain the pathophysiology, causes, signs, symptoms, and therapeutic management of cardiovascular diseases such as hypertension, peripheral vascular disease, unstable angina, acute myocardial infarction, cardiomyopathy and inflammation, heart failure, pulmonary embolism, cardiac tamponade, arrhythmia, and pacing failure |
|  | 28 | Understand the normal cardiac cycle. |
|  | 29 | Understand the normal cardiac conduction system of stimulation |
|  | 30 | Understand the determinants of cardiac output |
|  | 31 | Understand the determinants of blood pressure |
|  | 32 | Understand the determinants of central venous pressure |
|  | 33 | Understand the effects of ventilation and intrathoracic pressure on the cardiovascular system |
| Observation, monitoring and assessment of the cardiovascular system | 34 | Appropriate hemodynamic monitoring on adult critically ill patients |
|  | 35 | Assess the arterial pressure waveforms |
|  | 36 | Assess the central venous pressure values and waveforms |
|  | 37 | Assess the pulmonary artery catheter readings and waveforms |
|  | 38 | Assess the dynamic indicators such as SVV |
|  | 39 | Assess the capillary reflex |
|  | 40 | Assess the limb and skin temperatures |
|  | 41 | Assess blood test results related to circulatory function |
|  | 42 | Understand normal ECG waveforms |
|  | 43 | Assess the common arrhythmias (atrial tachycardia, ventricular tachycardia, atrial fibrillation, ventricular fibrillation, atrioventricular block, etc.) |
|  | 44 | Appropriate continuous ECG monitoring |
|  | 45 | Correctly measure 12-lead ECG |
|  | 46 | Report abnormalities or changes in the circulatory system to medical personnel at the appropriate time |
| Nursing Practice for the Cardiovascular System | 47 | Manage critically ill patients with rapidly deteriorating circulatory status with a medical team |
|  | 48 | Manage patients after vascular or cardiac surgery |
|  | 49 | Coordinate patient activities and implement nursing care taking into account circulatory load |
| Administration of medicines related to the circulatory(cardiovascular) system | 50 | Explain infusion therapy (understand types of infusions, need for infusions, follow guidelines, accurately record fluid balance) |
|  | 51 | Explain the indications, contraindications, mechanism of action, and side effects of cardiovascular agents (vasoactive medicines, inotropic medicines, antiarrhythmic medicines, etc.) |
|  | 52 | Assess clinical findings and adjust increase/decrease of circulatory agonists under the direction of a physician |
|  | 53 | Ask the physician to adjust infusion management according to the patient's physiological status |
| Shock Management | 54 | Explain the classification and treatment of shock (cardiogenic, abnormal blood distribution, obstructive, hypovolemic) |
|  | 55 | Understand the balance between oxygen supply and demand |
|  | 56 | Understand the causes of lactic acid abnormalities |
|  | 57 | Perform necessary assessment and monitoring for patients in shock (EKG, blood pressure, temperature, urine output, IV fluids, skin, limb temperature, blood tests) |
|  | 58 | Understand treatment protocols according to different shock classifications and assist in treatment |
|  | 59 | Appropriately assess and respond to electrolyte, glucose, and acid-base disturbances in patients with shock |
| **1.3. Gastrointestinal system and nutrition** | | |
| Anatomy and physiology of the gastrointestinal system and understanding of diseases | 60 | Understand the anatomy and physiology of the gastrointestinal system (including the gastrointestinal tract, pancreas, gallbladder, and liver) |
|  | 61 | Understand gastrointestinal tract function. |
|  | 62 | Understand the endocrine and exocrine systems |
|  | 63 | Explain the pathophysiology, causes, signs, symptoms, and therapeutic management of gastrointestinal disorders such as gastrointestinal bleeding/ischemia/perforation, bowel obstruction, esophageal varices, pancreatitis, cirrhosis/liver failure, and abdominal compartment syndrome |
|  | 64 | Understand the process of critical illness from physiologic changes associated with chronic and acute liver and biliary tract disorders |
|  | 65 | Understanding of Bacterial Translocation |
|  | 66 | Understand the effects associated with increased intra-abdominal pressure |
| Observation, monitoring and assessment of the gastrointestinal system | 67 | Determine the need for monitoring patients at risk for deterioration related to gastrointestinal tract function |
|  | 68 | Assess diarrhea, constipation, etc. using appropriate scales |
|  | 69 | Assess the need to measure intra-abdominal pressure and measure it appropriately |
|  | 70 | Assess blood test results related to gastrointestinal function |
|  | 71 | Report abnormalities or changes in the gastrointestinal system to the appropriate health care provider |
| Nursing Practice for the Gastrointestinal System | 72 | Manage patients with gastrointestinal diseases such as liver failure and shock due to gastrointestinal diseases |
|  | 73 | Manage patients after typical abdominal surgeries such as Hartmann, esophagectomy, and bowel resection |
|  | 74 | Manage patients with increased intra-abdominal pressure |
|  | 75 | Manage drains associated with abdominal disease |
|  | 76 | Manage gastric tubes in critically ill patients |
|  | 77 | Manage the rectal balloon catheter for severe diarrhea |
| Administration of medicines related to the gastrointestinal system | 78 | Assess the effectiveness of medicines related to the gastrointestinal system and adjust care and treatment according to the patient's condition |
|  | 79 | Understand medicine specificity (e.g., contraindications to grinding) |
|  | 80 | Understand intestinal motility and motility enhancers, laxatives, anti-stimulants, insulin/hypoglycemic agents, probiotics, steroids, antidiarrheals, and antisecretory agents |
| Nursing Practice for Nutritional Management | 81 | Understand basic ideas about nutrition, including individual nutritional needs (calorie requirements, protein levels, vitamins and minerals, etc.) |
|  | 82 | Assess the body's response (including nutritional status and laboratory values) related to nutritional administration |
|  | 83 | Refer to the patient's past medical history and explain diseases affecting gastrointestinal function |
|  | 84 | Explain nutritional therapy according to guidelines and other recommendations |
|  | 85 | Provide consultation to specialists to adjust nutritional therapy according to the patient's condition. |
| Nursing Practice for Dysphagia | 86 | Explain the mechanism of swallowing |
|  | 87 | Explain risk factors for dysphagia |
|  | 88 | Assess swallowing function (water-only test, repeated saliva-only test, food test, etc.) |
|  | 89 | Provide consultation to specialists regarding the causes of dysphagia and therapeutic rehabilitation methods. |
|  | 90 | Prepare the environment for eating, adjust food form and thickening, and provide appropriate dietary assistance. |
| **1.4. Renal system** | | |
| Anatomy and Physiology of the Renal and Urinary System and Understanding of Disease | 91 | Understand the anatomy and physiology of the renal and urinary system |
|  | 92 | Understand the function of the kidneys. |
|  | 93 | Understand electrolyte excretion. |
|  | 94 | Explain factors affecting renal blood flow |
|  | 95 | Explain electrolyte abnormalities |
|  | 96 | Explain the pathophysiology, causes, signs, symptoms, and therapeutic management of renal and urologic diseases, including renal disorders (acute kidney injury, chronic kidney injury, end-stage renal disease) and electrolyte abnormalities |
| Observation, monitoring and assessment of the renal and urinary system | 97 | Determine the need for monitoring of people at risk of renal function decline |
|  | 98 | Understand how to monitor fluid status, fluid delivery, and renal function in patients at risk for renal impairment |
|  | 99 | Understand how to measure and record fluid volume |
|  | 100 | Understand the causes of fluid loss (drains, gastrointestinal system, hemorrhage, insensate excretion, etc.) |
|  | 101 | Assess blood collection results in patients with renal and urinary dysfunction |
|  | 102 | Appropriately report abnormalities or changes in the renal and urinary system |
| Nursing Practice for Renal and Urinary System | 103 | Insert a urinary catheter |
|  | 104 | Understand the need for urinary catheters and their harmful effects during insertion, and observe them correctly. |
|  | 105 | Understand guideline-based treatment strategies for acute kidney injury |
| Management of medicines related to the renal and urinary system | 106 | Understand diuretics, glucose and insulin, salbutamol (nebulizer), calcium, sodium bicarbonate, and other medicines related to the kidney |
|  | 107 | Assess the effectiveness of infusions and medications related to the renal and urologic system and adjust care and treatment according to the patient's condition |
| **1.5. Endocrine and metabolic systems** | | |
| Anatomy and physiology of the endocrine and metabolic systems and understanding of disease | 108 | Understand the anatomy and physiology of the endocrine and metabolic systems |
|  | 109 | Explain the pathophysiology, causes, signs, symptoms, and therapeutic management of metabolic and endocrine disorders such as diabetes, adrenal insufficiency, and thyroid |
| Observation, monitoring and assessment of endocrine and metabolic systems | 110 | Explain the need for blood glucose monitoring in critically ill patients |
|  | 111 | Assess periodic blood glucose readings based on patient condition and the effects of medications used. |
|  | 112 | Provide necessary observations and appropriate assessment for patients with abnormal thyroid function (e.g., hyperthyroidism including crisis, hypothyroidism) |
|  | 113 | Assess blood test results related to endocrine and metabolic functions |
|  | 114 | Appropriate reporting of endocrine and metabolic abnormalities and changes |
| Nursing Practice for Endocrine and Metabolic Systems | 115 | Manage glycemic control in critically ill patients |
|  | 116 | Explain the care and management of patients with diabetic ketoacidosis, nonketotic hyperosmotic syndrome, and hypoglycemia |
|  | 117 | Explain the care and management of patients with abnormal thyroid function (hyperthyroidism including crisis, hypothyroidism) |
| Management of medicines related to endocrine and metabolic systems | 118 | Understand insulin and other medicines related to the endocrine and metabolic systems |
|  | 119 | Assess the effects of medicines related to the endocrine and metabolic systems and adjust care and treatment according to the patient's condition |
|  | 120 | Manage insulin administration based on blood glucose levels |
| **1.6. Cerebral nervous system** | | |
| Anatomy and physiology of the nervous system and understanding of disease | 121 | Understand the anatomy and physiology of the cranial nervous system |
|  | 122 | Explain the state of cerebral hypertension |
|  | 123 | Understand primary and secondary brain injury |
|  | 124 | Explain the impact of neurological disorders on the patient |
|  | 125 | Explain the pathophysiology, causes, signs, symptoms, and treatment management of cerebral hemorrhage, cerebral infarction, subarachnoid hemorrhage, seizures, encephalitis and meningitis, and other neurological disorders |
| Observation, monitoring and assessment of the brain nervous system | 126 | Assess and accurately record Glasgow Coma Scale (GCS) |
|  | 127 | Observe and assess pupils (size, shape, responsiveness) |
|  | 128 | Assess cranial nervous system function such as paralysis of the extremities |
|  | 129 | Understand the mechanisms of normal control of cerebral perfusion and intracranial pressure (ICP) and explain normal parameters of intracranial pressure (ICP) and cerebral perfusion pressure (CPP) |
|  | 130 | Recognize signs and symptoms of elevated intracranial pressure (ICP) |
|  | 131 | Monitor hemodynamics with consideration of the impact on cranial nerve damage |
|  | 132 | Ability to adequately monitor patients with brain nervous system disorders during hypothermia therapy. |
|  | 133 | Recognize abnormalities in continuous EEG monitoring for seizure patients and others. |
|  | 134 | Appropriately report abnormalities or changes in the cranial nervous system |
| Nursing Practice for the Brain Nervous System | 135 | Provide nursing care for patients with neurological dysfunctions |
|  | 136 | Understand the symptoms associated with seizures and manage and care for them with safety in mind |
|  | 137 | Manage the airway in relation to impaired consciousness |
|  | 138 | Adjust body position with an understanding of changes in cerebral pressure and cerebral blood flow |
|  | 139 | Know the indications for CT and MRI of the head |
|  | 140 | Provide care that is aware of the potential impact on intracranial pressure (ICP) |
|  | 141 | Understand the patient's condition from CT and MRI of the head (including readings) |
|  | 142 | Devise and practice appropriate methods of care to maintain adequate cerebral perfusion pressure or mean arterial pressure when intracranial pressure (ICP) cannot be monitored |
|  | 143 | Assess nursing care for increased intracranial pressure (ICP) and adjust care plan accordingly |
| Manage of medicines related to the nervous system | 144 | Prepare and administer therapeutic agents (osmotic treatments, analgesics, muscle relaxants, anticonvulsants, catecholamines, steroids, antihypertensive medications, etc.) for the treatment of cranial nervous system disorders |
|  | 145 | Assess fluid balance with consideration of the impact on cranial nerve damage and consult on the administration of appropriate fluids |
|  | 146 | Assess the effects of medicines related to the nervous system, consult with patients on their conditions, and coordinate their care |
| **1.7. Skin/musculoskeletal system** | | |
| Anatomy and physiology of the skin, musculoskeletal system, and understanding of disease | 147 | Understand the anatomy and physiology of the skin, musculoskeletal system |
|  | 148 | Explain ICU-AW |
|  | 149 | Explain risk factors for ICU-AW |
|  | 150 | Explain risk factors for pressure ulcers, pressure wounds on medical devices, and skintightness in critically ill patients |
| Observation, monitoring and assessment of skin, musculoskeletal system | 151 | Monitor and assess musculoskeletal system related issues such as muscle strength (MMT, MRC score, grip strength, etc.) and ADL assessment |
|  | 152 | Observe skin and wounds (including trauma and post-operative) and detect abnormalities early |
|  | 153 | Assess pressure ulcer risk (e.g., Braden scale) in critically ill patients |
|  | 154 | Continuous monitoring (e.g. DESIGN-R) and evaluation of pressure ulcers in critically ill patients |
|  | 155 | Appropriately report abnormalities or changes in the skin, musculoskeletal system |
| Nursing practice for skin, musculoskeletal system | 156 | Explain how to prevent and treat ICU-AW |
|  | 157 | Prepare, treat, and care for wounds accordingly |
|  | 158 | Explain how to prevent pressure ulcers, pressure wounds on healthcare-related equipment, and skin lesions in critically ill patients. |
|  | 159 | Explain the care of critically ill patients with pressure ulcers, pressure wounds and skin lesions on medical related equipment. |
| Knowledge of pharmacology as it relates to the skin, musculoskeletal system | 160 | Explain how to treat pressure ulcers, pressure wounds and skin lesions in critically ill patients. |
|  | 161 | Assess the effects of medicines related to the skin, musculoskeletal system, and adjust care and treatment according to the patient's condition |
| **1.8. Infectious diseases, blood and immune system** | | |
| Anatomy and physiology of infectious diseases, blood and immune system and disease understanding | 162 | Understand the anatomy and physiology of the blood and immune system |
|  | 163 | Explain the pathophysiology, causes, signs, symptoms, and treatment management of sepsis |
|  | 164 | Understand the definition and diagnostic criteria for sepsis |
|  | 165 | Understand the septic shock |
| Nursing Practice for Infectious Diseases, Hematology, and Immune System | 166 | Recognize patients with infectious diseases who are at risk of worsening their condition |
|  | 167 | Refer to sepsis guidelines and practice appropriate care bundles |
|  | 168 | Take necessary infection control precautions for individual patients and treatment environments |
|  | 169 | Take precautions against multimedicine-resistant bacteria |
| Prevention and Nursing Care of Device-Related Infections | 170 | Explain risk factors for catheter-related bloodstream infections |
|  | 171 | Explain how to prevent and treat catheter-related bloodstream infections |
|  | 172 | Explain risk factors for ventilator-associated pneumonia and ventilator-associated events (VAE) |
|  | 173 | Explain how to prevent and treat ventilator-associated pneumonia and ventilator-associated events (VAE) |
|  | 174 | Explain risk factors for urinary catheter-related urinary tract infection |
|  | 175 | Explain how to prevent and treat urinary catheter-related urinary tract infections |
|  | 176 | Explain risk factors for surgical site infection |
|  | 177 | Explain how to prevent and treat surgical site infections |
| Nursing Practice for Blood Coagulation Abnormality | 178 | Practice nursing care for patients with blood coagulation abnormalities |
|  | 179 | Explain indications, contraindications, and side effects of blood products |
|  | 180 | Safely administer blood products and follow facility policy |
|  | 181 | Understand hematological disorders such as major bleeding requiring massive blood transfusions, immunosuppression, and immunodeficiency |
|  | 182 | Explain TRALI, GVHD, and other transfusion-related adverse events |
|  | 183 | Understand disseminated intravascular coagulation syndrome |
|  | 184 | Understand the indications and administration of anticoagulants |
|  | 185 | Explain risk factors for deep vein thrombosis |
|  | 186 | Explain how to prevent and treat deep vein thrombosis |
| **1.9. Other diseases** | | |
| Nursing Practice for Resuscitation and Sudden Changes | 187 | Prepared to anticipate, prevent, and recognize life-threatening situations and intervene |
|  | 188 | Assess and respond to rapidly changing patient and treatment situations during resuscitation and emergencies |
|  | 189 | Recognize early warning signs quickly and consult with other nursing staff, medical team members, and others as needed |
|  | 190 | Consider care and clinical priorities in response to emergencies and unforeseen circumstances |
|  | 191 | Recognize and assess critically ill patients who deteriorate rapidly and manage them toward condition stabilization |
|  | 192 | Respond to cardiopulmonary arrest (BLS, ACLS) |
|  | 193 | Identify and respond to lethal arrhythmias |
|  | 194 | Manage post-resuscitation, including management of airway, breathing, circulation, arrhythmias, and abnormal metabolic states |
| Nursing Practice for Trauma | 195 | Explain the pathophysiology, causes, signs, symptoms, and treatment management of multiple trauma, head trauma, thoracoabdominal trauma, extremity pelvic trauma, and spinal trauma |
|  | 196 | Provide nursing care for trauma patients |
| Nursing Practice for Burns | 197 | Explain the pathophysiology, causes, signs, symptoms, and management of burns |
|  | 198 | Provide nursing care for burn patients |
| Nursing practice for abnormal body temperature | 199 | Explain the pathophysiology, causes, signs, symptoms, and treatment management of hyperthermia and hypothermia, including heat stroke, malignant hyperthermia, and malignant syndrome |
|  | 200 | Provide nursing care for patients with hyperthermia and hypothermia, including heat stroke, malignant hyperthermia, and malignant syndromes |
|  | 201 | Handle, assist, observe, and manage the use of temperature control devices. |
| **1.10. Treatment equipment management** | | |
| Noninvasive ventilatory management | 202 | Explain the indications for non-invasive ventilation |
|  | 203 | Explain the advantages and disadvantages of noninvasive ventilation |
|  | 204 | Explain the physiological and psychological effects of noninvasive ventilation on patients |
|  | 205 | Correctly prepare non-invasive ventilators (including circuit assembly, proper parameter and alarm settings) |
|  | 206 | Use of a heated humidifier for non-invasive ventilators |
|  | 207 | Provide initiate, administer, and wean from noninvasive ventilation |
|  | 208 | Explain the different types of masks and mask fitting methods |
|  | 209 | Prevent complications of non-invasive ventilation (e.g., skin problems) |
|  | 210 | Assist in the daily living of patients on non-invasive ventilation while dealing with their symptoms |
|  | 211 | （Adjust sedation and other treatments and ventilator settings according to the condition of the patient being non-invasively ventilated (based on physician orders) |
|  | 212 | Correctly troubleshoot non-invasive ventilatory equipment |
| Invasive ventilatory management | 213 | Explain the indications for invasive ventilation |
|  | 214 | Explain the physiological and psychological effects of invasive mechanical ventilation on patients |
|  | 215 | Invasive mechanical ventilator is properly prepared (including circuit assembly, proper parameter and alarm settings) |
|  | 216 | Use of a Heated humidifier for invasive mechanical ventilators |
|  | 217 | Initiate, manage, and wean from invasive mechanical ventilation |
|  | 218 | Assist with daily living while dealing with symptoms of patients on invasive ventilation |
|  | 219 | Identify ventilation modes and ventilator settings explain them |
|  | 220 | Explain how to prevent complications of invasive mechanical ventilation (e.g., lung protective ventilation for ventilator lung injury) |
|  | 221 | （Adjust treatment and mechanical ventilator settings, such as sedation, according to the patient's condition during invasive ventilation (based on physician's orders) |
|  | 222 | Troubleshoot invasive mechanical ventilator (high pressure, low pressure, low tidal volume, high peak airway pressure, high breathing and other alarms, power failure response, equipment malfunction, etc.) |
|  | 223 | Explain the significance of practicing the invasive mechanical ventilator Care Bundle |
|  | 224 | Coordinate and implement personnel needed for prone therapy during invasive mechanical ventilation |
|  | 225 | Assist with bronchoscopy during invasive mechanical ventilation |
| Nursing Practices Related to Oral Intubation | 226 | Explain the indications, advantages, and disadvantages of intubation |
|  | 227 | Understand the intubation process and prepare necessary supplies and medications |
|  | 228 | Assist with intubation and extubation |
|  | 229 | Manage the safe and accurate positioning of the intubation tube as appropriate, set the appropriate cuff pressure, and select the appropriate method of immobilization for the patient. |
|  | 230 | Assess the patency of the artificial airway and assess and respond to urgent airway problems (airway obstruction due to sputum, unscheduled tracheal tube removal or dislodgement, pneumothorax). |
|  | 231 | Assess the need for suctioning during ventilation using indicators such as cough, secretions, hypoxemia, restlessness, high airway pressure, and hemodynamic changes |
|  | 232 | Explain the advantages and disadvantages of suction above the cuff |
|  | 233 | Practice measures to minimize the causes and risks of emergency reintubation |
|  | 234 | Properly care for the oral health of intubated patients |
|  | 235 | Capable of manual ventilation using a bag-valve device |
| Nursing Practices Related to Tracheostomy | 236 | Understand percutaneous tracheostomy, surgical tracheostomy, and minitrack, their indications, advantages, and disadvantages |
|  | 237 | Understand when a tracheostomy is appropriate |
|  | 238 | Explain the indications and necessity of emergency airway clearance procedures such as emergency cricothyrotomy |
|  | 239 | Explain complications after tracheostomy |
|  | 240 | Understand the process of performing a tracheostomy and be prepare and assist with necessary supplies and medications |
|  | 241 | Care for and observe patients before, during, and after tracheostomy |
|  | 242 | Properly manage (including position, fixation, and cuff pressure) tracheostomy tubes, including speaking valves |
|  | 243 | Observe patients for possible physical and psychological effects associated with tracheostomy and respond accordingly |
| Knowledge of renal replacement therapy | 244 | Explain the types of renal replacement therapy and the advantages and disadvantages of each method |
|  | 245 | Select individual patient treatment modes and set individual prescribed treatment goals, as well as adjust treatment according to coagulation, electrolyte, and acid-base goals |
|  | 246 | Explain complications during renal replacement therapy and how to minimize them (thrombocytopenia/clotting disorders, anemia, circulation, electrolytes, bleeding, hypothermia, infection, thrombosis/embolism) |
|  | 247 | The impact of renal replacement therapy on the metabolism of medicines and how to deal with it appropriately |
|  | 248 | Appropriate equipment monitoring (access pressure, membrane pressure, etc.) and troubleshooting during renal replacement therapy |
|  | 249 | Report any abnormalities or changes during renal replacement therapy to the appropriate health care provider |
|  | 250 | Provide psychological care for patients undergoing renal replacement therapy |
|  | 251 | Accurate fluid balance including cumulative balance after renal replacement therapy to record |
| Drain management | 252 | Correctly assemble the equipment needed to insert a thoracic drain |
|  | 253 | Assist with insertion and removal of chest drains |
|  | 254 | Appropriately manage patients with chest drains inserted |
|  | 255 | Understand the indications for thoracic drainage according to the patient's condition |
|  | 256 | Assist with emergency decompression of tension pneumothorax |
|  | 257 | Troubleshoot problems related to various drains, including ventricular drains and abdominal drains |
|  | 258 | Manage various types of drains, including ventricular drains and abdominal drains |
|  | 259 | Provide psychological care to patients with various drain insertions, including ventricular drains and abdominal drains |
| Management of defibrillators and pacemakers | 260 | Explain indications and timing of defibrillation |
|  | 261 | Explain indications and timing of pacing |
|  | 262 | Assist, observe, and handle the use of an implantable pacemaker |
|  | 263 | Assist, observe, and respond to the use of external (temporary) pacing |
| Management of circulatory assist devices (IABP, IMPELLA, etc.) | 264 | Understand the indications for circulatory assist devices (IABP, IMPELLA, etc.) |
|  | 265 | Manage and care for patients on circulatory assist devices (IABP, IMPELLA, etc.) |
| Management of auxiliary circulatory equipment (VA-ECMO, etc.) | 266 | Explain indications for assistive circulatory devices (e.g., VA-ECMO) |
|  | 267 | Manage and care for patients on assisted circulatory devices (e.g., VA-ECMO) |
| Management of pulmonary function assist devices (e.g. VV-ECMO) with membrane type artificial lungs | 268 | Explain indications for pulmonary function assist devices (e.g., VV-ECMO) using membrane-type artificial lungs |
|  | 269 | Manage and care for patients using (e.g., VV-ECMO, a pulmonary function support device using membrane-type artificial lungs) |
| Various line management | 270 | Assist with insertion and removal of arterial lines, central venous lines, and catheters for renal replacement therapy |
|  | 271 | Properly manage arterial lines, central venous lines, and catheters for renal replacement therapy |
| **1.11. organ transplantation** | | |
| Indications and Legal Understanding of Organ Transplantation | 272 | Understand basic institutional rules and procedures for organ transplantation |
|  | 273 | Understand the cause of irreversible disorientation |
|  | 274 | Understand patient confidentiality issues related to organ transplantation |
| Management of organ transplant patients | 275 | Work with internal and external coordinators (organ transplant team) |
|  | 276 | Manage donor patients according to the manual |
| **2. Caring** | | |
| **2.1 Nursing Diagnosis and Planning** | | |
| Collect appropriate information on critically ill patients | 277 | Use appropriate evidence-based assessment techniques to determine a patient's overall needs (including learning needs, psychological support, and psycho-social needs) |
|  | 278 | Use appropriate evidence-based assessment techniques and tools to get a complete picture of the patient |
|  | 279 | Prioritize data collection activities according to patient characteristics |
| Appropriate assessment of critically ill patients | 280 | Relate collected data to current and predicted future conditions according to patient characteristics |
|  | 281 | Analyze data from multiple sources to determine patient and family needs |
| Develop appropriate care plans for critically ill patients | 282 | Identify and prioritize evidence-based interventions to promote and restore health and prevent further disease and disability |
|  | 283 | Work with patients, families, and multidisciplinary teams to develop a plan |
|  | 284 | Assess and communicate pertinent and relevant data to the team |
| Assessment of Critical Care | 285 | Assess the effectiveness of interventions in a timely manner and modify treatment and care modalities as needed to achieve expected outcomes |
|  | 286 | Properly record the results of the evaluation |
| **2.2 Relief of discomfort symptoms** | | |
| Nursing Practice for Pain | 287 | Assess, prevent, and treat pain, including pharmacological and non-pharmacological interventions |
| Nursing practice for restlessness and sedation | 288 | Assess, prevent, and treat restless and sedated states, including pharmacological and non-pharmacological interventions |
| Nursing Practice for Delirium | 289 | Explain risk factors for delirium in critically ill patients |
|  | 290 | Responsive to the assessment, prevention, and treatment of delirium, including pharmacological and non-pharmacological interventions |
| Nursing Practice for Insomnia | 291 | Respond to the assessment, prevention, and treatment of insomnia and sleep disorders, including pharmacological and non-pharmacological interventions |
| Nursing practice for anxiety, depression, fear, etc. | 292 | Respond to the assessment, prevention, and treatment of anxiety, including pharmacological and non-pharmacological interventions |
|  | 293 | Ensure that patients and families make informed treatment and care choices and understand the consequences |
|  | 294 | Provide supportive care and coaching for patients and families during difficult procedures |
|  | 295 | Minimize the psychological impact related to critical illness and treatment on the patient and family |
|  | 296 | Respond to patients and families in the event of bereavement or traumatic events |
| Nursing practice for other discomfort symptoms | 297 | Assess, prevent, and treat discomfort symptoms such as dry mouth, dyspnea, and fatigue, including pharmacological and non-pharmacological interventions |
| **2.3. Rehabilitation of critically ill patients / PICS** | | |
| Understanding and implementing the rehabilitation needs of patients and families | 298 | Explain PICS/PICS-F and its risks and prevention methods |
|  | 299 | Explain the challenges of rehabilitation in the critical care area |
|  | 300 | Understand why critically ill patients need ongoing rehabilitation |
|  | 301 | Set rehabilitation goals (short, medium, and long term) appropriate for individual critically ill patients |
|  | 302 | Coordinate rehabilitation of critically ill patients according to guidelines and other recommendations |
|  | 303 | Understand and recognize rehabilitation prescriptions for critically ill patients |
|  | 304 | Understand the issue of individual patient diversity and how it affects the patient's rehabilitation needs |
| Nursing practice for maintenance and recovery of physical function | 305 | Practice positioning of critically ill patients |
|  | 306 | Practice joint range of motion training in critically ill patients |
|  | 307 | Practice respiratory rehabilitation of critically ill patients |
|  | 308 | Practice early mobilization of critically ill patients |
|  | 309 | Implement practices to maintain motor function and improve ADLs in critically ill patients |
|  | 310 | Understand risk assessment related to rehabilitation of critically ill patients and rehabilitate them |
| Nursing practice for maintenance and recovery of cognitive function and mental health | 311 | Explain rehabilitation methods to prevent and ameliorate cognitive dysfunction and delirium |
|  | 312 | Explain how to prevent and improve mental health disorders (depression, anxiety, PTSD) in patients and families |
| **2.4. End-of-life care** | | |
| Evaluation of patient prognosis | 313 | Assess the patient severity |
|  | 314 | Appropriate prognostic evaluation of critically ill patients |
| Withholding or withdrawing treatment | 315 | Communicate end-of-life care plan to patient's family |
|  | 316 | Discuss end-of-life care plans with the patient's family |
|  | 317 | Provide symptom relief and individualized treatment and care plans for terminally ill patients according to the patient's needs |
|  | 318 | In a multidisciplinary team, assess futility and other factors and consider withholding or terminating treatment as appropriate |
|  | 319 | Consider end-of-life care options appropriate for the patient, taking into account the patient's and family's wishes, including the choice of ward after treatment is discontinued. |
|  | 320 | Understand the procedures for forming and recording agreements regarding treatment cessation, including legal restrictions on treatment cessation or withholding, mental capacity laws, and ethical principles |
|  | 321 | Understand what care is appropriate for the patient after discontinuation of treatment |
| Assess, monitor, and observe terminally ill patients | 322 | Observe and assess symptoms of terminally ill patients, including pain, nausea, agitation, dyspnea, and fatigue. |
|  | 323 | Recognize that the palliative approach integrates palliative care principles (symptom management, patient-centered care) throughout the patient's life and is not just for the last few days of life |
|  | 324 | Work with patients, families, and professionals to determine end-of-life wishes, identify available resources, and implement strategies to promote dignity, comfort, and quality care at the end of life |
|  | 325 | Seek and incorporate patient and family input to provide quality end-of-life care that meets their needs |
| Utilize appropriate resources for end-of-life issues | 326 | Work with multidisciplinary teams to facilitate palliative care and end-of-life discussions, decisions, and care |
|  | 327 | Maintain ongoing communication with family and professionals regarding palliative approaches/care at the end of life and provide ongoing emotional support |
|  | 328 | Appropriate resources are available to guide and identify effective and possible solutions to ethically complex situations. |
| **2.5. Provide an ICU environment that promotes healing** | | |
| Patient and family environmental management | 329 | Provide a safe, healing and caring environment that respects the individual |
|  | 330 | Effectively facilitate patient orientation |
|  | 331 | Appropriate assessment of patient's sensory perceptions, such as noise, illumination, smell, care, and other stimuli |
|  | 332 | Manage the environment appropriately for the patient's senses, including noise, lighting, smells, care, and other stimuli |
|  | 333 | Minimize environmental risk factors that could cause physical harm or injury to patients, families, and health care professionals |
|  | 334 | Promote proper day/night sleep cycles in critically ill patients |
| Improve patient and family services | 335 | Work with colleagues to propose service improvements |
|  | 336 | Obtain and act upon feedback and experiences from patients, caregivers and service users |
|  | 337 | Question existing ways of doing things and propose new ways to current performance and culture |
|  | 338 | Contribute to new initiatives within the department led by experienced colleagues |
|  | 339 | Contribute to the quality of health care improvement efforts taking place in the department |
|  | 340 | Care for patients of all ages across the lifespan to restore, support, promote, and maintain physiological and psychosocial stability |
| **3. Advocate and moral agency** | | |
| **3.1. Support decision-making** | | |
| Support for decision-making | 341 | Take into account patient and family preferences for treatment and intervention |
|  | 342 | Consider patient and family preferences for treatment and interventions |
|  | 343 | Act ethically and responsibly and participate in ethical discussions and decision-making processes |
|  | 344 | Provide information to help patient families make decisions |
|  | 345 | Leverage and use a variety of evidence in making complex decisions |
|  | 346 | Support problem solving and decision making for a variety of clinical situations for the patient's family |
| **3.2. Ethical practice** | | |
| Practices based on ethical principles and compliance with the law | 347 | Understand national regulations and laws (e.g., Medical Care Act, Health and Nursing Code, etc.) related to patient care and health care delivery |
|  | 348 | Comply with national regulations and laws (e.g., Medical Care Act, Health and Nursing Code, etc.) related to patient care and health care delivery |
|  | 349 | Understand ethical practices, autonomy, equality and diversity, and legal regulations to protect the rights of patients and families |
|  | 350 | Demonstrate awareness of patient autonomy, consent, and relevant local and national laws |
|  | 351 | Articulate an understanding of ethical principles relevant to the delivery of critical care |
|  | 352 | Embracing equality and diversity and respecting without discrimination age, gender, religion, sexual orientation, race, disability, sentiments, social status, etc. |
|  | 353 | Practices that consistently nurture the autonomy, dignity, values, beliefs, and rights of patients and their families |
|  | 354 | Provide care in a fair and equitable manner that meets the diverse needs of patients, families, and the community |
|  | 355 | Take responsibility for your own words and actions. |
|  | 356 | Promote ethical accountability and integrity in relationships, organizational decision-making, and resource management |
|  | 357 | Document patient care and its ongoing evaluation in a clear, concise, accurate and timely manner, while respecting privacy and confidentiality of personal information |
|  | 358 | Protect patient confidentiality within legal regulations |
|  | 359 | Take into consideration and practice standards of confidentiality, data protection, and documentation |
|  | 360 | Report unethical, illegal, or impaired conduct |
| Practices for resolving ethical issues | 361 | Use available resources in making ethical decisions |
|  | 362 | Make ethical decisions using available resources |
|  | 363 | Address the needs of patients and families facing unanticipated treatment, quality of life, and end-of-life decisions |
|  | 364 | Maintain a therapeutic and professional nurse-patient relationship within the appropriate role assignment |
|  | 365 | Contribute to the resolution of ethical issues involving patients, families, and multidisciplinary teams |
| **3.3 Patient and Family Communication** | | |
| Respectful communication with patients and families | 366 | Respect others' viewpoints in discussions with patients and families |
|  | 367 | Attentive to patient and family wishes and communicate information in a respectful manner according to needs, developmental stage, and level of understanding |
|  | 368 | Communicate effectively with patients and families regarding treatment and nursing plans and the patient's clinical status |
|  | 369 | Effectively communicate and explain difficult clinical information using terms and language that patients and families can understand |
|  | 370 | Effectively communicate with and support patients and families in crisis situations |
|  | 371 | Effective communication in complex situations. |
|  | 372 | Communicate what may be considered bad news to the patient and family in a polite and compassionate manner |
| Accountability for nursing practice (informed consent) | 373 | Explain their roles and responsibilities to patients and families and facilitate relationship building |
|  | 374 | Recognize and comply with regulations and laws related to the nursing role |
|  | 375 | Explain treatment options to patients and families and assist in facilitating informed decision making |
|  | 376 | Enable patients and families to make informed choices and understand the consequences |
| **4. Evidence-based practices** | | |
| **4.1. Quality assurance and improvement of care (PDCA)** | | |
| Quality Assessment and Improvement Activities | 377 | Actively pursue new knowledge and skills to provide quality nursing care |
|  | 378 | Ability to question medical practices as needed to improve safety and quality |
|  | 379 | Continuously monitor and assess care processes and outcomes to determine best practices for individuals, groups, and populations |
|  | 380 | Ensure that nursing practices are in accordance with formal procedures and regulations |
|  | 381 | Implement practices to improve care processes and outcomes based on evidence, expertise, and patient preferences |
| Promoting Evidence-Based Practices | 382 | Know how to search for evidence and literature using available resources |
|  | 383 | Keep updated and current in knowledge of evidence-based practice |
|  | 384 | Know what care practices are based on the guidelines |
|  | 385 | Contribute through evidence-based critical care nursing practice |
| **5. Collaboration and management ability** | | |
| **5.1. Unit management** | | |
| Logistics Management and Cost Awareness | 386 | Implement cost and waste reduction practices |
| Participation in organizational activities | 387 | Participate in committees, councils, and multidisciplinary teams |
| Healthy work environment and work style | 388 | Demonstrate an understanding of work-life balance |
|  | 389 | Balance one's own work-life balance |
|  | 390 | Contribute to the creation and maintenance of a healthy work environment |
|  | 391 | Indicate flexibility and patient focus in a rapidly changing environment |
|  | 392 | Ask for an appropriate number of staff with the knowledge and skills to care for critically ill patients when needed |
|  | 393 | Adhere to standards governing the behavior of health care professionals to create a healthy work environment that promotes cooperation, respect, and trust |
|  | 394 | Understand, embrace and actively participate in change management and related processes |
|  | 395 | Maintain a physical and psychosocial environment that promotes safety, security, and optimal health |
|  | 396 | Comply with working conditions, employment rights, and work environment considerations set by the state and the facility |
| **5.2. Team management** | | |
| Membership and Followership | 397 | Recognize common team objectives and respect team decisions |
|  | 398 | Adopt a team approach and recognize and appreciate efforts, contributions and compromises |
|  | 399 | Recognize, respect, and promote collaboration with team members |
|  | 400 | Recognize and participate in the need to support and debrief with colleagues and each other |
|  | 401 | Support others in providing good patient care and better service |
|  | 402 | Opportunities to be more effective by working with others |
|  | 403 | Share care plans and other information widely with team members |
|  | 404 | Report appropriately to team members and other professionals using SBARs and other tools. |
|  | 405 | Respect others' viewpoints and actively ask their opinions in discussions with other healthcare professionals. |
|  | 406 | Share knowledge and collaborate with others to create change and positive outcomes |
|  | 407 | Proactively and independently derive solutions to recurring problems and issues |
|  | 408 | Clearly recognize their role, responsibilities, and purpose within the nursing team |
| leadership | 409 | Have a sense of role in the team in order to form an effective team |
|  | 410 | Demonstrate effective interpersonal communication, leadership, negotiation, and conflict resolution skills to build positive relationships with colleagues, patients, and families |
|  | 411 | Facilitate collaboration among team members |
|  | 412 | Foster a spirit of transparency, trust, and respect among team members |
|  | 413 | Provide leadership and support for service improvement initiatives within the department |
|  | 414 | Emphasize the value of shared responsibility in decision-making and support shared leadership and coordination roles |
|  | 415 | Encourage open and constructive discussion and respect the diverse viewpoints of others |
| Management and coordination of care and operations, stress management | 416 | Manage activities on a daily or shift basis |
|  | 417 | Manage patient care to ensure that it is effective and efficient |
|  | 418 | Plan and prioritize work and workload during the shift according to patient needs |
|  | 419 | Adjust priorities and change work plans as circumstances dictate |
|  | 420 | Delegate appropriate tasks to other team members as needed |
|  | 421 | Assess and adjust the workload and workload of staff and other team members to meet the changing needs of the patient's family |
|  | 422 | Effectively manage the workload in your department |
|  | 423 | Identify and, if necessary, coordinate the work of professional or other positions (including clerical, etc.) |
|  | 424 | Recognize and effectively utilize coping strategies to deal with stressful situations in the clinical environment |
|  | 425 | Know how to prevent stressors in the workplace |
|  | 426 | Observe and address the health status of other team members |
| Use of other experts (consultation) | 427 | Facilitate sharing of information and resources (other professionals) among staff |
|  | 428 | Assess individual patient needs and select the available resources (other professionals) to achieve the best outcomes |
|  | 429 | Additional and necessary resources (other professionals) can be suggested to improve nursing practice and quality of care for critically ill patients |
|  | 430 | Assist patients and families in identifying and securing appropriate services to address their health-related needs, depending on available resources (other professionals) |
|  | 431 | Work to resolve conflicts between multiple professions and contribute to interprofessional care planning |
|  | 432 | Consultation with appropriate staff/professionals to develop/review care plans and facilitate continuity of care |
|  | 433 | Contribute to the development of effective interprofessional relationships to meet the needs of patients and their families |
|  | 434 | Collaborate to provide optimal care, respecting each other's roles, responsibilities, and abilities. |
|  | 435 | Choose appropriate methods in communicating with other professionals |
|  | 436 | Explain and facilitate their roles and responsibilities to other members of the health care community |
|  | 437 | Offer own professional perspective in discussions with multidisciplinary teams |
|  | 438 | Ask questions of all members of the multidisciplinary team about the care process and the rationale supporting decisions |
| **5.3. Medical safety** | | |
| Safety Culture and Incident Reporting | 439 | Understand the value of quality indicators (e.g., length of stay, ventilatory time, prevention of infections) for patient outcomes |
|  | 440 | Understand their role in influencing the quality of safe and effective critical care services |
|  | 441 | Build a culture of safety for patients, families, and providers |
|  | 442 | Promote a safe culture of learning from and responding to risk |
|  | 443 | Identify and demonstrate observations and concerns regarding safety, hazards, and errors in the care and practice environment |
|  | 444 | Understand and comply with local and national regulations and laws regarding the prevention, reporting, and monitoring of adverse events, including medication errors, adverse events, and equipment malfunctions |
|  | 445 | Actively participate in the recognition, response, disclosure, reporting, and prevention of recurrence of adverse or potential risks or incidents |
|  | 446 | Appropriately report adverse or potential risks or incidents in accordance with unit, hospital, and national policies and protocols |
|  | 447 | Quickly identify adverse or potential risks or incidents that have occurred in the hospital |
|  | 448 | Respond quickly to protect patient safety |
|  | 449 | Respond appropriately to adverse or potential risks or incidents and take necessary actions |
|  | 450 | Consider measures to prevent recurrence of medical accidents |
|  | 451 | Use safe and effective written, verbal, telephone, and electronic communication strategies |
|  | 452 | Understand and participate in clinical audits, including patient safety, environmental safety, and medical equipment maintenance |
| Awareness of infection prevention and toxic exposure | 453 | Safe handling and disposal of contaminated, biological, chemical, and toxic materials in accordance with local and national policies and protocols |
|  | 454 | Promote proper use of disposable items in the department |
|  | 455 | Communicate environmental health risks and exposure reduction measures to patients, families, and health care professionals |
| Handling of complex medical equipment | 456 | Maintain a safe medical environment (e.g., bed space organization, emergency equipment) |
|  | 457 | Safe use of medical equipment required for critical care |
|  | 458 | Safely assist or perform advanced and complex procedures required for medical device operation |
| Safe medication and medication manegement | 459 | Maintain and update knowledge of medications administered in the critical care setting |
|  | 460 | Knowing the right patient, the right time, the right route, the right medicine, the right dose, the right label, the right calculation, and the relevant details will ensure that medication errors are prevented |
|  | 461 | Follow and promote unit, hospital, and national guidelines for critical care medications |
| **5.4. In-hospital and out-of-hospital patient transport** | | |
| Evaluation of indications for intra- and extra-hospital patient transport | 462 | Work as a team to perform a comprehensive risk assessment to ensure the patient is suitable for transport |
|  | 463 | Explain the potential risks associated with transporting critically ill patients |
|  | 464 | Explain what may happen to the patient during transport |
|  | 465 | Determine if the transport is urgent and time-critical |
|  | 466 | Review patient priorities and needs and coordinate patient transfers |
|  | 467 | Reassess safety and risk factors prior to transport |
| Preparation for in-hospital and out-of-hospital patient transport | 468 | Understand the preparation process for in-hospital/out-of-hospital transport of critically ill patients |
|  | 469 | Understands the roles of team members in arranging and conducting intra-hospital and inter-hospital transports |
|  | 470 | Assess patient needs prior to transport |
|  | 471 | Assess patient's clinical condition prior to transfer out of the ICU |
|  | 472 | Anticipate potential problems with conveyance and plan to reduce risk |
|  | 473 | Prepare spare battery packs and alternative equipment in case of failure |
|  | 474 | Prepare medical records, radiology results, and recent blood test results, etc prior to transport |
|  | 475 | Prepared transport monitoring equipment |
|  | 476 | Explain the care that should be responsibly performed during transport |
|  | 477 | Assess the competence and skills of transport personnel |
|  | 478 | Consider contingency plans/backups |
|  | 479 | Communicate with the receiving hospital that will transport the patient. |
|  | 480 | Explain the types of transport available and their advantages (out-of-hospital: ambulance/helicopter transport, in-hospital: stretcher, wheelchair, etc.) |
|  | 481 | Prepare and complete necessary paperwork such as transport documents, progress notes, nursing plans, etc. |
|  | 482 | Work with patients, families, and multidisciplinary teams to facilitate effective and safe transport across care settings |
| Practice and evaluation during transport | 483 | Monitor and assess changes in physiological indicators of patients during transport |
|  | 484 | Manage necessary medications to patients during transport |
|  | 485 | Appropriately recognize situations that may affect the quality and safety of critical care transfers |
|  | 486 | Identify issues and problems that can be remedied during transport |
|  | 487 | Reflect on the conveyance experience |
|  | 488 | Coordinate the provision of ongoing care, including specialist treatment and observation, for patients being transported |
| Communication necessary during transport, such as signing off | 489 | Provide information and informed consent for transporting a conscious patient |
|  | 490 | Communicate with family members during transport and provide ongoing status reports as needed |
|  | 491 | Communicate openly with multiple disciplines to minimize risks associated with patient transfers, intra-hospital transfers, and transitions of care |
|  | 492 | Properly contact the receiving unit upon departure when moving out |
|  | 493 | Communicate patient status and physiological requirements to transport team/staff |
|  | 494 | Share information with the team related to safety, risk assessment, and planning for the unexpected regarding transport |
|  | 495 | Appropriate transfer of patient status to receiving unit at time of transfer |
| **6. Education and self-development ability** | | |
| **6.1. Self-development** | | |
| Introspective Practice | 496 | Actively ask feedback on your practice from patients, family members, colleagues, and professionals |
|  | 497 | Reflect on nursing practice through an introspective and self-aware approach |
|  | 498 | Reflect on current activities and identify how future activities should be developed |
|  | 499 | Deepen self-awareness and recognize one's own strengths and limitations |
|  | 500 | Admit failure and treat it as a learning opportunity |
|  | 501 | Change behavior based on feedback and reflection |
| Self-development and professional development | 502 | Understand the need for education in critical care |
|  | 503 | Take initiative in one's own educational and professional development |
|  | 504 | Know how to access learning and teaching opportunities/resources that support continuing professional development |
|  | 505 | Participate in continuing education programs and learning activities to develop and maintain clinical and professional competence |
|  | 506 | Recognize and, when possible, participate in local, national, and international conferences and committees related to critical care nursing |
|  | 507 | Facilitates the transfer of information and professional development through writing, publishing, and presentations to professional and general audiences |
|  | 508 | Keep records and activity histories that provide evidence of professional competence and lifelong learning |
| **6.2. Education** | | |
| Fostering a learning environment | 509 | Participate in activities that facilitate the teaching and learning of others in a multidisciplinary team |
|  | 510 | Support individual and group self-development in a critical care environment |
|  | 511 | Contribute to a work environment conducive to professional education |
| Educational involvement with colleagues | 512 | Provide regular feedback to colleagues and acknowledge successes and need for improvement as needed |
|  | 513 | Provide formal or informal constructive feedback on colleagues' practices and role performance |
|  | 514 | Behave in a professional manner as a role model for juniors and students |
|  | 515 | Serve as a resource, educator, role model, preceptor, advocate, and mentor to students, colleagues, and multidisciplinary team members |
|  | 516 | Demonstrate ability to be an effective mentor and role model as needed |
|  | 517 | Share acquired educational knowledge, experience, and ideas with peers |
|  | 518 | Teach and review practices with colleagues |
| training of new recruits | 519 | Support newcomers. |
|  | 520 | Assist newcomers in developing their skills |
|  | 521 | Motivate and encourage newcomers to develop their skills |
|  | 522 | Guide, supervise, and assess nursing care provided by less experienced colleagues |
|  | 523 | Coach colleagues on tasks, goals, processes, and performance standards |
| Learning Support for Patients and Families | 524 | Educate patients and families as needed |
|  | 525 | Provide patients and families with the study information they need |

*Abbreviations: Acute lung injury; ALI, Acute respiratory distress syndrome; ARDS, Cannot ventilate and cannot intubate; CVCI, Stroke Volume Variation; SVV, Glasgow Coma Scale; GCS, Intracranial pressure; ICP, Cerebral perfusion pressure; CPP, ICU acquired weakness; ICU-AW, Ventilator associated event; VAE, Transfusion related acute lung injury; TRALI, Graft versus host disease; GVHD, Basic Life Support; BLS, Advanced Cardiovascular Life Support; ACLS, Intra Aortic Balloon Pumping; IABP, Venoarterial-extracorporeal membrane oxygenation; VA-ECMO, Venovenous-extracorporeal membrane oxygenation; VV-ECMO, Post intensive care syndrome/ post intensive care syndrome - family; PICS/PICS-F, Post Traumatic Stress Disorder; PTSD, Activities of Daily Living; ADL, Quality of Life; QOL, Medical Research Council; MRC, manual muscle testing; MMT
